# Increasing intra- and inter-subtype HIV diversity despite declining HIV incidence in Uganda

**DOI:** 10.1101/2024.03.14.24303990

**Authors:** Seungwon Kim, Godfrey Kigozi, Michael A. Martin, Ronald M. Galiwango, Thomas C. Quinn, Andrew D. Redd, Robert Ssekubugu, David Bonsall, Deogratius Ssemwanga, Andrew Rambaut, Joshua T. Herbeck, Steven J. Reynolds, Brian Foley, Lucie Abeler-Dörner, Christophe Fraser, Oliver Ratmann, Joseph Kagaayi, Oliver Laeyendecker, M. Kate Grabowski

## Abstract

HIV incidence has been declining in Africa with scale-up of HIV interventions. However, there is limited data on HIV evolutionary trends in African populations with waning epidemics. We evaluated changes in HIV viral diversity and genetic divergence in southern Uganda over a twenty-five-year period spanning the introduction and scale-up of HIV prevention and treatment programs using HIV sequence and survey data from the Rakai Community Cohort Study, an open longitudinal population-based HIV surveillance cohort. *Gag* (p24) and *env* (gp41) HIV data were generated from persons living with HIV (PLHIV) in 31 inland semi-urban trading and agrarian communities (1994 to 2018) and four hyperendemic Lake Victoria fishing communities (2011 to 2018) under continuous surveillance. HIV subtype was assigned using the Recombination Identification Program with phylogenetic confirmation. Inter-subtype diversity was estimated using the Shannon diversity index and intra-subtype diversity with the nucleotide diversity and pairwise TN93 genetic distance. Genetic divergence was measured using root-to-tip distance and pairwise TN93 genetic distance analyses. Evolutionary dynamics were assessed among demographic and behavioral sub-groups, including by migration status. 9,931 HIV sequences were available from 4,999 PLHIV, including 3,060 and 1,939 persons residing in inland and fishing communities, respectively. In inland communities, subtype A1 viruses proportionately increased from 14.3% in 1995 to 25.9% in 2017 (p<0.001), while those of subtype D declined from 73.2% in 1995 to 28.2% in 2017 (p<0.001). The proportion of viruses classified as recombinants significantly increased by more than four-fold. Inter-subtype HIV diversity has generally increased. While p24 intra-subtype genetic diversity and divergence leveled off after 2014, diversity and divergence of gp41 increased through 2017. Inter- and intra-subtype viral diversity increased across all population sub-groups, including among individuals with no recent migration history or extra-community sexual partners. This study provides insights into population-level HIV evolutionary dynamics in declining African HIV epidemics following the scale-up of HIV prevention and treatment programs. Continued molecular surveillance may provide a better understanding of the dynamics driving population HIV evolution and yield important insights for epidemic control and vaccine development.

## Introduction

Human immunodeficiency virus type 1 (HIV-1) remains a significant public health threat. In 2022, an estimated 1.3 million new HIV cases and 630,000 AIDS-related deaths occurred globally, of which more than one-third were in eastern and southern Africa (UNAIDS, 2023). Currently, there is no HIV vaccine to prevent HIV acquisition, partly due to extensive virus genetic diversity (Barouch & Korber, 2010; Haynes et al., 2016; Ng’uni et al., 2020). Within the predominant HIV-1 group M viruses, there exist seventeen distinct HIV-1 subtypes and sub-subtypes, and numerous inter and intra-subtype recombinant forms (LANL, 2023; Mendes Da Silva et al., 2021; B. S. Taylor et al., 2008; Yamaguchi et al., 2020). Similar to the global burden of cases and deaths, HIV genetic diversity is greatest in Africa (Hemelaar et al., 2019; Lihana et al., 2012; Peeters et al., 2003; Vidal et al., 2000). Thus, an effective preventative vaccine for African populations would mostly likely need to protect against a breadth of HIV subtypes and their derivatives (Burton et al., 2012; Burton & Hangartner, 2016; Sadanand et al., 2016; Steichen et al., 2019).

Notwithstanding its significance for HIV vaccine and treatment (e.g., broadly neutralizing antibodies) development, data on accurate and up-to-date longitudinal trends in HIV genetic diversity are limited, particularly in eastern and southern Africa (Hemelaar et al., 2019; Lihana et al., 2012). Prior analyses of publicly available HIV sequence data, mostly conducted before widespread expansion of HIV treatment and prevention programs, showed increasing viral diversity in Africa as the epidemic grew (Hemelaar et al., 2019, 2020; Lihana et al., 2012; Peeters et al., 2003). However, spatially and temporally uneven sampling of HIV genomes may have biased these earlier assessments. Moreover, by the end of 2022, 83% of people living with HIV (PLHIV) in eastern and southern Africa were using antiretroviral therapy (ART), resulting in a decline of more than 57% in the number of new HIV infections since 2010 (UNAIDS, 2023). However, it remains unclear to what extent such significant changes in the African HIV epidemic dynamics have affected viral diversity within the region.

Understanding patterns in HIV evolution, particularly in response to roll-out of treatment and prevention programs, may provide information critical for treatment development and insights into epidemic dynamics for virus control efforts. For example, declining diversity might indicate that less common strains of virus have disappeared, concentration of transmission within population sub-groups, or an epidemic increasingly driven by a small number of local outbreaks. In contrast, increasing diversity might highlight the growing role of virus introduction due to migration, having sex with non-local partners (Grabowski et al., 2014; Olawore et al., 2018), or indicate diversifying selection.

Here, we used data from more than 30 continuously surveyed communities in southern Uganda over a nearly three-decade long period to evaluate trends in intra- and inter-subtype HIV viral diversity. Data were collected prior to and during the scale-up of HIV prevention and treatment programs through the Rakai Community Cohort Study (RCCS), an open population-based household census and HIV surveillance cohort. Following scale up of antiretroviral therapy and voluntary medical male circumcision programs beginning in 2004, HIV incidence rapidly declined in RCCS communities (Grabowski et al., 2017), consistent with regional trends (Joshi et al., 2021). RCCS communities have among the highest HIV case burdens worldwide (Chang et al., 2016), with extensive HIV subtype diversity, including co-circulation of HIV-1 subtypes A1, C, D, and numerous recombinant forms (Arroyo et al., 2006; Capoferri et al., 2020; Conroy et al., 2010; Lamers et al., 2020). Our analyses provide insight into population-level HIV evolutionary dynamics in a high burden region with declining incidence following HIV program scale-up and may inform development and strategic deployment of HIV prevention and treatment interventions.

## Methods

### Study design and population

The RCCS is conducted in inland agrarian and semi-urban trading communities (HIV prevalence 9-26%) and hyperendemic Lake Victoria fishing communities (HIV prevalence 37-43%) in southern Uganda (Chang et al., 2016). The RCCS has been ongoing since 1994 in inland agrarian and trading communities and since 2011 in fishing communities. Longitudinal surveillance is based on a household census and individual survey conducted at ∼18 months intervals. The census enumerates all individuals in the household irrespective of age or presence in the community and documents residence status, including migrations into and out of the household. The survey follows the census and enrolls consenting adolescents and adults 15-49 years who are resident in the community. During the survey, demographic, sexual behavior (e.g., sex with partners outside community of residence), and HIV service utilization data (e.g., ART use and male circumcision status) are collected and blood samples are obtained for HIV testing and molecular epidemiology (Grabowski et al., 2017; Wawer et al., 1998). All RCCS participants are provided with HIV test results and offered free pre and post-test counseling with referral to prevention and treatment services. All study participants provide written informed consent at baseline and follow-up visits.

Here, we used HIV genomic sequence and survey data from PLHIV in 31 inland agrarian and semi-urban trading communities surveyed between November 1994 and May 2018 and four hyperendemic Lake Victoria fishing communities surveyed between November 2011 and August 2017.

### Sequencing

HIV sequence data has been generated using blood samples from RCCS participants living with HIV since the 1990s. Sequencing was attempted in some surveys for all participants with viremic HIV or not self-reporting ART use if viral load data was unavailable (Round 1: 1994-1995, Round 9: 2002-2003, and Round 13-18: 2008-2018) and only for selected participant visits for others. For blood samples collected prior to 2010, Sanger sequencing was used to generate consensus sequences for p24 in the g*ag* gene (HXB2: 1249-1704) and gp41 in the *env* gene (HXB2: 7857-8260) (C. Yang et al., 2000). For blood samples collected from 2010 onwards (beginning with survey round 14), near full-length genomes were generated using a next generation sequencing (NGS) approach in collaboration with the PANGEA-HIV consortium (Abeler-Dörner et al., 2019; Pillay et al., 2015) using the Gall protocol (2010-2015) and the ve-SEQ HIV protocol (2015-2018) (Bonsall et al., 2020; Gall et al., 2012). Resulting deep-sequencing reads were assembled using *shiver* to generate HIV consensus sequences (Wymant et al., 2018). Near full-length HIV genomes collected between January 2010 and May 2018 were aligned using MAFFT (v 7.52) and the HIV-1 regions of p24 and gp41 were extracted from the aligned sequences to compare consensus HIV genome sequences between earlier and later surveys (Katoh & Standley, 2013).

### Subtyping analyses

The Recombinant Identification Program (RIP) version 3.0 was used to assign HIV subtypes to p24 and gp41 HIV genomes (Fig. 1) (Siepel et al., 1995). A RIP window of 100 bases and 90% confidence threshold was used to calculate similarity scores (s-distance) with the four most common circulating HIV-1 group M subtypes in Rakai (consensus reference sequences of A1, C, D, and G) (Supplementary Table 1). HIV sequences were assigned as a pure subtype if they had a single best match with one of the reference sequences with 90% confidence. If there were more than two matches with the reference sequences with 90% confidence in any region, the length of the second best-matching reference sequence was measured, and if it was shorter than 25 bases, viruses were classified as pure subtypes. Otherwise, the s-distance gap between the first and second best-matching reference sequences in the potential recombinant region was used to further classify them; if the gap was greater than 2%, sequences were subtyped as recombinants, otherwise they were classified as potential recombinants. Next, pure, definitive, and potential recombinant sequences were aligned. Maximum Likelihood (ML) phylogenetic trees were generated from these alignments in IQ-TREE (v2.1.4) (Minh et al., 2020), and using resulting trees, recombinant sequences were classified as either pure subtype or recombinant viruses. We further analyzed HIV subtypes of p24 and gp41 from RCCS participants who had available sequence data from both genes in the same surveys. HIV subtypes from these two gene regions were combined if the sequence data were obtained from the same individuals in the same survey round. For example, a participant with subtype A1 virus in both genes, would be considered as having a subtype A1 infection whereas a participant with subtype A1 in one gene and subtype D in the other would be considered an A1/D recombinant infection. We also assigned HIV subtypes to p24 and gp41 sequences using COMET and REGA subtyping tools, both with a 70% of bootstrap support threshold, and results were compared to subtype assignment based on RIP and phylogenetic analyses (Supplementary Tables 2 and 3) (Pineda-Peña et al., 2013; Struck et al., 2014). The Cochran-Armitage test for trend in proportions was used to access the statistical significance of longitudinal trends in HIV-1 subtype distributions over calendar time using *stats* package (v4.3.2) (part of base R) in R (v4.3.1) (R Core Team, 2021).

**Figure 1.**
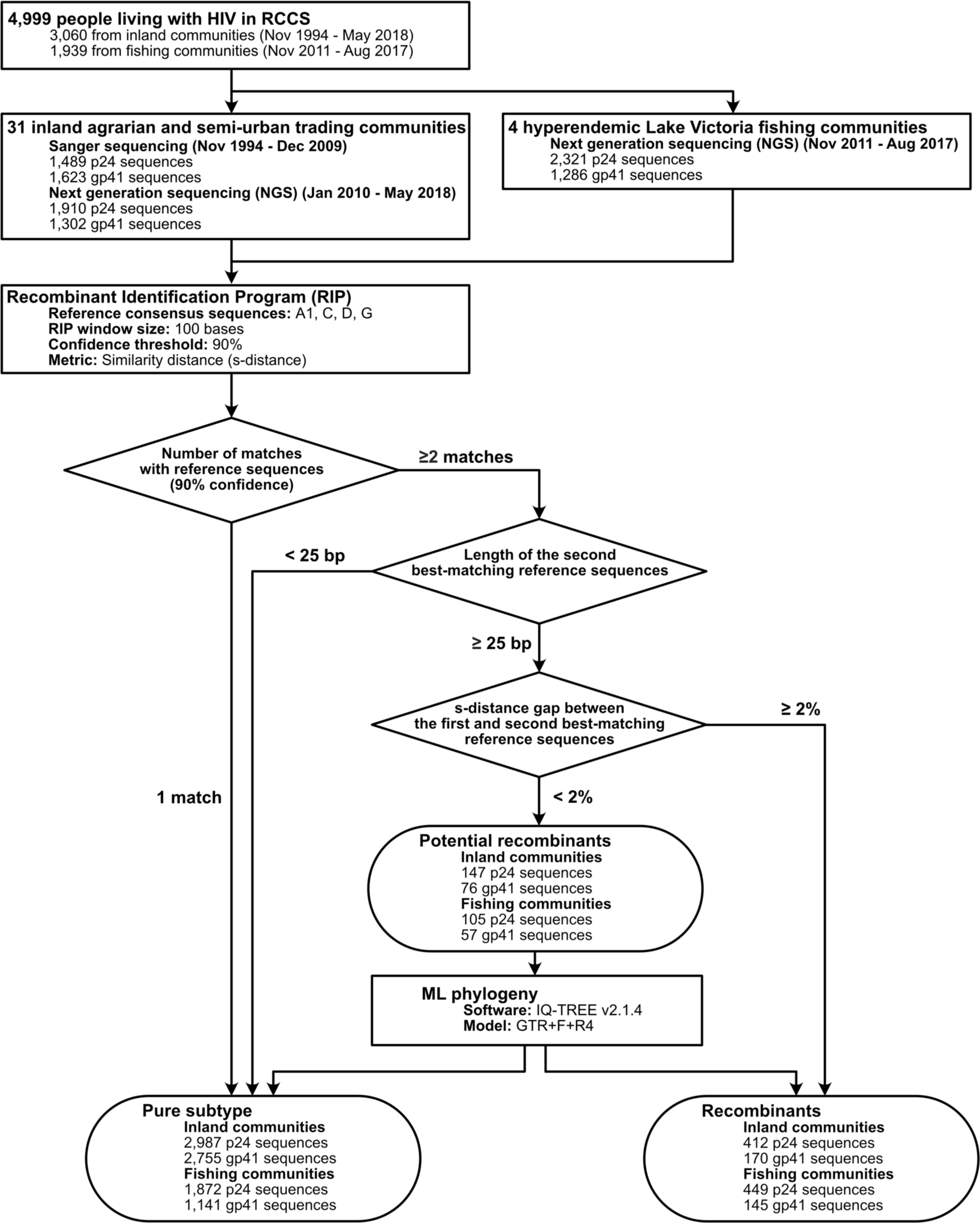
HIV genome sequence data collected from RCCS participants living with HIV in 31 inland agrarian and semi-urban trading communities and four hyperendemic Lake Victoria fishing communities and subtype analyses using the Recombinant Identification Program (RIP) and phylogenetic reconstruction

### Diversity measures

#### Shannon diversity index

We used the Shannon Diversity Index (SDI) to evaluate longitudinal trends in HIV inter-subtype diversity in inland and fishing communities in Rakai. SDI quantifies biodiversity and species richness within a population (Shannon, 1948). SDI was calculated as follows: *H′ =* 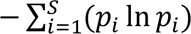, where S is the total number of HIV subtypes in one survey round and *p_i_* is the proportion of subtype *i* sequences relative to the total number of sequences in one survey round. A high *H′* value indicates greater inter-subtype diversity and more even distribution of sequences among subtypes, whereas low *H′* implies lower intra-subtype diversity and the dominance of particular subtypes. For these analyses inter-subtype recombinants of the same subtypes were classified as a single subtype irrespective of breakpoint.

We measured SDI in p24 and gp41 within inland and fishing communities overall and then within various population sub-groups including by sex, age group, occupation, HIV-related risk behavior (numbers of past year sexual partners, sexual partners outside the community), incident case (i.e. participants testing positive for the first time with an HIV seronegative test result at the prior visit), and history of recent migration into study communities. Occupations were grouped differently for inland (agricultural, trading/trucking, other) and fishing communities (fisherfolk/bar workers/restaurant workers, other) because their demographic composition varies significantly between inland and fishing communities (Chang et al., 2016). SDI values were calculated using *poppr* R package (v2.9.4) and 95% bootstrap confidence intervals were generated using the package *boot* (v1.3.28.1) (part of base R) in R (Kamvar et al., 2014).

#### Nucleotide diversity

We next estimated nucleotide diversity (*π*) and pairwise TN93 genetic distance in p24 and gp41 to measure longitudinal trends in both inter- and intra-subtype genetic diversity. The nucleotide diversity index is a measure of genetic variation that quantifies the average number of nucleotide differences per site between all possible pairs of sequences in a population. *π* was calculated as follows: 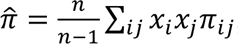, where n is the number of sequences, *x_i_* and *x_j_* are the frequencies of the *i*th and *j*th sequences, and *π_ij_* is the number of nucleotide differences per site between *i*th and *j*th sequences. Firstly, the overall nucleotide diversity statistics, regardless of subtype, were calculated to evaluate the degree of genetic variation of HIV within the entire population in each survey round. Then, subtype-specific estimates within HIV subtypes A1 and D was measured to evaluate temporal trends in the genetic diversity of two most common HIV subtypes in our dataset. Similar to SDI, we calculated HIV nucleotide diversity in the entire population and within various population sub-groups. Nucleotide diversity measures were calculated using *pegas* package (v1.2) and 95% bootstrap confidence intervals were calculated using *boot* package in R (Paradis, 2010).

#### Genetic divergence

We also calculated root-to-tip distances and pairwise TN93 genetic distances to evaluate longitudinal trends of population-level HIV genetic divergence in p24 and gp41. Firstly, subtype-specific (A1, C, and D) p24 and gp41 ML phylogenetic trees were reconstructed in IQ-TREE using the General Time Reversible model with empirical base frequencies and free rate model with four categories (GTR+F+R4) (Z. Yang, 1995). ML phylogenetic trees were rooted using sequences from other subtypes as the outgroup. Patristic distances from the root to the tips of phylogenetic trees (root-to-tip distances) were measured and plotted against the sampling times of sequences. The evolutionary rates and time to the most recent common ancestor were calculated using a simple linear regression. Root-to-tip distances were calculated using *adephylo* package (v1.1.16) (Jombart et al., 2010).

Additionally, we measured pairwise TN93 genetic distances of HIV subtype A1, C, and D between the HIV consensus sequences of Los Alamos National Laboratory (LANL) HIV Database subtype references and HIV sequence data collected in Rakai (Tamura & Nei, 1993). LANL reference sequences of subtypes A1, C, and D (Supplementary Table 1) were retrieved and the consensus sequences were generated using the Consensus Maker Tools (https://www.hiv.lanl.gov/content/sequence/CONSENSUS/consensus.html). Pairwise TN93 genetic distances were calculated using *ape* package (v5.7.1) in R (Paradis & Schliep, 2019).

### HIV evolution and selective pressures

Lastly, we assessed if there was evidence of selective virus evolution. First, we calculated Tajima’s D statistic in each survey to quantify deviations from the expected genetic diversity under neutral evolution, where most of the genetic variation arises from neutral mutations. Tajima’s D is calculated as the normalized difference between the nucleotide genetic diversity and the expected number of segregating sites (Tajima, 1989). A substantial deviation of Tajima’s D from 0 indicates the potential effects of natural selection, changes in population size, or other non-neutral processes. To prevent overestimation of segregating sites, p24 and gp41 sequences with gaps greater than 10 bases were excluded for the calculation of Tajima’s D statistic. The p-values of Tajima’s D statistic were calculated using a beta distribution with a mean of zero and a variance of one. Tajima’s D statistic was calculated using *pegas* package in R.

We further calculated the dN/dS ratio (*ω*) to evaluate a population-level gene-wide selective pressure on HIV-1 subtypes A1 and D p24 and gp41 (Z. Yang & Bielawski, 2000). This metric measures the ratio of non-synonymous substitutions to synonymous substitutions in a gene to assess evolutionary forces acting on the gene within a population. A value of *ω* < 1 suggests purifying or negative selection, indicating that non-synonymous changes are being removed or are not favored within a population. A value of *ω* = 1 implies neutral evolution, whereas *ω* > 1 indicates positive or diversifying selection, suggesting that non-synonymous changes are favored and occur more frequently than synonymous changes. To estimate, we first reconstructed ML phylogenetic trees in IQ-TREE (v2.1.4) and from these trees, we then calculated *ω* in codeml in the PAML package (v4.9j) and BUSTED in the HyPhy package (v2.5.33) (Murrell et al., 2015; Z. Yang, 2007). For the dN/dS ratio analyses conducted in codeml, we subsampled down to 50 sequences from each survey round (n=400 in inland communities and n=176 in fishing communities) to maintain computational efficiency. Sensitivity analyses with five different random sequence sets were performed to assess the impact of down-sampling on the estimated parameters values.

## Results

### Population characteristics

Between 1994 and 2018, 46,565 individuals participated in the RCCS of whom 37,606 lived in inland agrarian and semi-urban trading communities and 8,959 in hyperendemic Lake Victoria fishing communities. Of these participants, 5,352 (14.2%) in inland communities and 3,402 (38.0%) in fishing communities were living with HIV. Among participants living with HIV, 3,060 (57.2%) in inland communities and 1,939 (57.0%) in fishing communities had HIV sequence data available in either p24 or gp41 genes from at least one survey visit. Of these participants with HIV sequence data available, 2,111 (69.0%) in inland communities and 1,121 (57.8%) in fishing communities had both p24 and gp41 sequence data. Individuals with HIV sequence data were less likely to be using ART, be male, and younger than individuals without HIV sequence data (Table 1, Supplementary Table 4 and 5). Despite more frequent sampling in later survey periods, HIV sequence data was less commonly available among persons living with HIV because of the scale-up of ART and resulting increases in HIV viral load suppression over calendar time.

**Table 1.** Demographic and HIV-related risk behavioral characteristics of RCCS participants living with HIV, with and without HIV sequence data in 31 inland agrarian and semi-urban trading communities between 1994 and 2018.

| Inland communities | R1 (Nov-1994 – Aug-1995) |  |  | R13 (June-2008 – Dec-2009) |  |  | R18 (Oct-2016 – May-2018) |  |  |
| --- | --- | --- | --- | --- | --- | --- | --- | --- | --- |
|  | PLHIV | PLHIV w/ seq | PLHIV wo/ seq | PLHIV | PLHIV w/ seq | PLHIV wo/ seq | PLHIV | PLHIV w/ seq | PLHIV wo/ seq |
| N | 1,162 | 604 | 558 | 1,218 | 703 | 515 | 1,648 | 269 | 1,379 |
| **Age** |  |  |  |  |  |  |  |  |  |
| Median age (IQR) | 29<br>(24, 35) | 30<br>(24, 35) | 28<br>(24, 35) | 33<br>(28, 39) | 31<br>(26, 37) | 35<br>(31, 41) | 36<br>(30, 42) | 30<br>(25, 35) | 37<br>(31, 42) |
| 15 – 24 | 315<br>(27.1%) | 154<br>(25.5%) | 161<br>(28.9%) | 170<br>(14%) | 130<br>(18.5%) | 40<br>(7.8%) | 173<br>(10.5%) | 66<br>(24.5%) | 107<br>(7.8%) |
| 25 – 34 | 531<br>(45.7%) | 279<br>(46.2%) | 252<br>(45.2%) | 542<br>(44.5%) | 346<br>(49.2%) | 196<br>(38.1%) | 559<br>(33.9%) | 130<br>(48.3%) | 429<br>(31.1%) |
| 35 + | 316<br>(27.2%) | 171<br>(28.3%) | 145<br>(26%) | 506<br>(41.5%) | 227<br>(32.3%) | 279<br>(54.2%) | 916<br>(55.6%) | 73<br>(27.1%) | 843<br>(61.1%) |
| **Sex** |  |  |  |  |  |  |  |  |  |
| Male | 440<br>(37.9%) | 251<br>(41.6%) | 189<br>(33.9%) | 425<br>(34.9%) | 263<br>(37.4%) | 162<br>(31.5%) | 507<br>(30.8%) | 116<br>(43.1%) | 391<br>(28.4%) |
| Female | 722<br>(62.1%) | 353<br>(58.4%) | 369<br>(66.1%) | 793<br>(65.1%) | 440<br>(62.6%) | 353<br>(68.5%) | 1,141<br>(69.2%) | 153<br>(56.9%) | 988<br>(71.6%) |
| **Occupation** |  |  |  |  |  |  |  |  |  |
| Agriculture | 654<br>(56.3%) | 331<br>(54.8%) | 323<br>(57.9%) | 637<br>(52.3%) | 359<br>(51.1%) | 278<br>(54%) | 870<br>(52.8%) | 115<br>(42.8%) | 755<br>(54.7%) |
| Trade/Truck | 150<br>(12.9%) | 90<br>(14.9%) | 60<br>(10.8%) | 168<br>(13.8%) | 94<br>(13.4%) | 74<br>(14.4%) | 216<br>(13.1%) | 52<br>(19.3%) | 164<br>(11.9%) |
| Restaurant/Bar | 32<br>(2.8%) | 18<br>(3%) | 14<br>(2.5%) | 58<br>(4.8%) | 32<br>(4.6%) | 26<br>(5%) | 103<br>(6.2%) | 10<br>(3.7%) | 93<br>(6.7%) |
| Other | 326<br>(28.1%) | 165<br>(27.3%) | 161<br>(28.9%) | 355<br>(29.1%) | 218<br>(31%) | 137<br>(26.6%) | 459<br>(27.9%) | 92<br>(34.2%) | 367<br>(26.6%) |
| **Number of sexual partners in the past year** |  |  |  |  |  |  |  |  |  |
| 0 – 1 | 959<br>(82.5%) | 488<br>(80.8%) | 471<br>(84.4%) | 794<br>(65.2%) | 478<br>(68%) | 316<br>(61.4%) | 1,020<br>(61.9%) | 149<br>(55.4%) | 871<br>(63.2%) |
| 2 + | 197<br>(17%) | 114<br>(18.9%) | 83<br>(14.9%) | 239<br>(19.6%) | 149<br>(21.2%) | 90<br>(17.5%) | 346<br>(21%) | 82<br>(30.5%) | 264<br>(19.1%) |
| No response | 6<br>(0.5%) | 2<br>(0.3%) | 4<br>(0.7%) | 185<br>(15.2%) | 76<br>(10.8%) | 109<br>(21.2%) | 282<br>(17.1%) | 38<br>(14.1%) | 244<br>(17.7%) |
| **Any sexual partner outside the community in the past year** |  |  |  |  |  |  |  |  |  |
| Yes | 193<br>(16.6%) | 101<br>(16.7%) | 92<br>(16.5%) | 333<br>(27.3%) | 202<br>(28.7%) | 131<br>(25.4%) | 482<br>(29.2%) | 101<br>(37.5%) | 381<br>(27.6%) |
| No | 968<br>(83.3%) | 502<br>(83.1%) | 466<br>(83.5%) | 700<br>(57.5%) | 425<br>(60.5%) | 275<br>(53.4%) | 884<br>(53.6%) | 130<br>(48.3%) | 754<br>(54.7%) |
| No response | 1<br>(0.1%) | 1<br>(0.2%) | 0<br>(0%) | 185<br>(15.2%) | 76<br>(10.8%) | 109<br>(21.2%) | 282<br>(17.1%) | 38<br>(14.1%) | 244<br>(17.7%) |
| **Recent history of migration** |  |  |  |  |  |  |  |  |  |
| Yes | 0<br>(0%) | 0<br>(0%) | 0<br>(0%) | 171<br>(14%) | 113<br>(16.1%) | 58<br>(11.3%) | 405<br>(24.6%) | 124<br>(46.1%) | 281<br>(20.4%) |
| No | 1,115<br>(96%) | 578<br>(95.7%) | 537<br>(96.2%) | 1,045<br>(85.8%) | 588<br>(83.6%) | 457<br>(88.7%) | 1,243<br>(75.4%) | 145<br>(53.9%) | 1,098<br>(79.6%) |
| Non-resident | 47<br>(4%) | 26<br>(4.3%) | 21<br>(3.8%) | 2<br>(0.2%) | 2<br>(0.3%) | 0<br>(0%) | 0<br>(0%) | 0<br>(0%) | 0<br>(0%) |
| **Self-reported ART use** |  |  |  |  |  |  |  |  |  |
| Yes | - | - | - | 268<br>(22%) | 9<br>(1.3%) | 259<br>(50.3%) | 1,277<br>(77.5%) | 64<br>(23.8%) | 1,213<br>(88%) |
| No | - | - | - | 731<br>(60%) | 524<br>(74.5%) | 207<br>(40.2%) | 332<br>(20.1%) | 186<br>(69.1%) | 146<br>(10.6%) |
| No response | - | - | - | 219<br>(18%) | 170<br>(24.2%) | 49<br>(9.5%) | 39<br>(2.4%) | 19<br>(7.1%) | 20<br>(1.5%) |
| **Viral load** |  |  |  |  |  |  |  |  |  |
| ≥ 1,000 | 462<br>(39.8%) | 277<br>(45.9%) | 185<br>(33.2%) | 690<br>(56.7%) | 573<br>(81.5%) | 117<br>(22.7%) | 312<br>(18.9%) | 269<br>(100%) | 43<br>(3.1%) |
| 50 ≤ VL < 1,000 | 24<br>(2.1%) | 11<br>(1.8%) | 13<br>(2.3%) | 117<br>(9.6%) | 71<br>(10.1%) | 46<br>(8.9%) | 56<br>(3.4%) | 0<br>(0%) | 56<br>(4.1%) |
| BD | 21<br>(1.8%) | 2<br>(0.3%) | 19<br>(3.4%) | 355<br>(29.1%) | 29<br>(4.1%) | 326<br>(63.3%) | 1,264<br>(76.7%) | 0<br>(0%) | 1,264<br>(91.7%) |
| No data | 655<br>(56.4%) | 314<br>(52%) | 341<br>(61.1%) | 56<br>(4.6%) | 30<br>(4.3%) | 26<br>(5%) | 16<br>(1%) | 0<br>(0%) | 16<br>(1.2%) |

### Subtype distribution

In inland communities, the most common HIV subtypes were A1 and D subtypes, followed by A1/D recombinant viruses, although the proportions of these subtypes varied over calendar time (Fig. 2A, Supplementary Fig. 1A, 2A, Supplementary Table 6, 7, and 8). Among participants who had both p24 and gp41 sequence data in the same survey, the proportion of subtype A viruses increased from 14.3% in 1995 to 25.0% until 2009 (p<0.001), and then leveled off around 25% (p=0.8515). Subtype D viruses consistently declined from 73.2% in 1995 to 28.2% in 2017 (p<0.001). Similar patterns were observed in HIV subtype distribution in p24 and gp41. Subtype A1 viruses proportionately increased from 16.6% in p24 and 21% in gp41 until 2010 (p24: p<0.001, gp41: p<0.001), after which stabilizing around ∼31% in p24 and ∼47% in gp41 between 2012 and 2017 (p24: p=0.8232, gp41: p= 0.9843). Meanwhile, the proportion of subtype D viruses significantly declined in p24 from 79.6% in 1995 to 44.7% in 2017 (p<0.001) and in gp41 from 75.9% in 1995 to 38.2% in 2017 (p<0.001).

**Figure 2.**
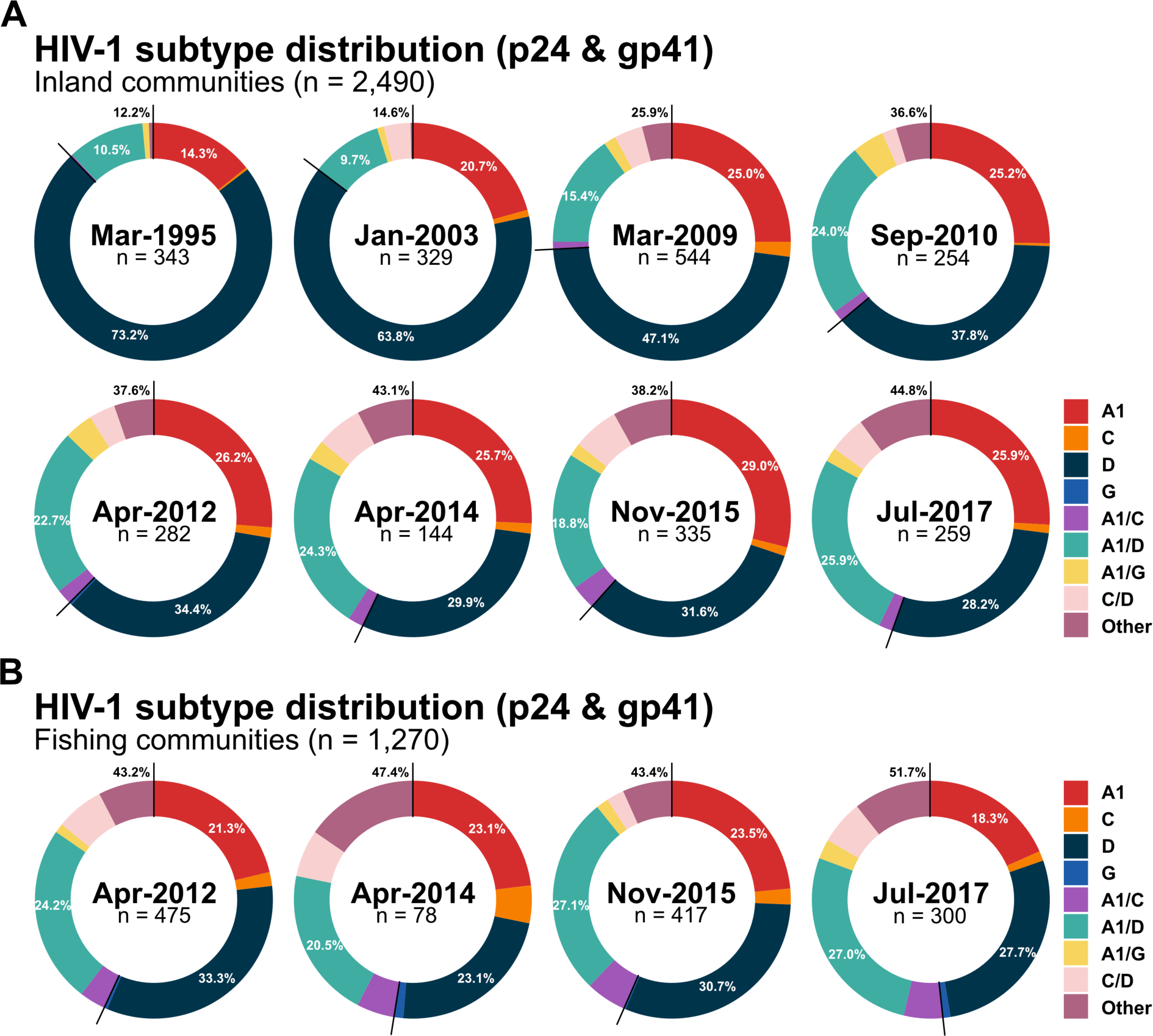
HIV subtype distribution of p24 and gp41 from RCCS participants living with HIV who had available sequence data from both genes in the same survey in 31 inland agrarian and semi-urban trading communities (Panel A) and four hyperendemic Lake Victoria fishing communities (Panel B) by calendar time. HIV subtypes of p24 and gp41 were independently assigned in the RIP and HIV subtypes of p24 and gp41 from the same individuals in the same survey were combined.

In the analyses of inland community participants with subtype data in both genes, the proportion of recombinants increased from 12.2% in 1995 to 44.8% in 2017 (p<0.001) (Fig 2A and Supplementary Table 6). The proportion of A1/D recombinant viruses substantially increased during the earlier surveys from 10.5% in 1995 to 24.0% in 2010 (p<0.001), after which leveled off around 23% (p=0.2046). Recombinant viruses of more than three different subtypes (e.g., A1/C/D, A/D/G) or rare recombinant forms in our sequence data (e.g., D/G, C/G) also significantly increased over calendar time, from 0.5% in 1995 to 10.0% in 2017 (p<0.001). By 2017, 44.8% of participants with sequence data in both genes were classified as having an inter-subtype recombinant infection. p24 sequences were significantly more likely to be classified as recombinants over the analysis period. The proportion of recombinants in p24 significantly increased from 2.7% in 1995 to 21.4% in 2017 (p<0.001) (Supplementary Figure 1A and Supplementary Table 7), in gp41 from 2.4% in 1995 to 9.5% in 2017 (p<0.001) (Supplementary Fig. 2A and Supplementary Table 8). In both genes, the most common recombinant infection was A1/D recombinants.

In Lake Victoria fishing communities, the proportion of viruses classified as either subtype A1 or D did not substantially change between 2012 and 2017 (Fig. 2B, Supplementary Fig. 1B, 2B, Supplementary Table 9, 10, and 11). Overall, recombinant viruses were slightly more common in fishing communities than inland communities, but these differences were not statistically significant in either p24 or gp41. Among individuals with subtype data in both genes, the proportions of subtypes A1, D, and A1/D recombinant viruses were generally similar to those in inland communities over the analysis period. Similar to inland communities, recombinants were more frequently detected in p24 than in gp41, and were usually A1/D, A1/C, or C/D recombinants.

We further stratified HIV subtypes by participant demographic and HIV-related risk behavioral characteristics across three calendar epochs for inland and fishing communities: pre-ART (1994-2004), ART roll-out and expansion (2008-2013), and Universal Test and Treatment era (2013-2018) (Supplementary Fig. 3-8). Overall, the proportion of subtype A1 and recombinant viruses increased, whereas subtype D viruses proportionally decreased regardless of population sub-groups over calendar time.

#### Inter-subtype diversity as measured by Shannon Diversity Index (SDI)

Next, we evaluated trends in inter-subtype HIV diversity by calculating the SDI in p24 and gp41 genes over calendar time for both inland and fishing communities (Fig. 3). Overall, the SDI increased in p24 through the study period regardless of community type. The SDI also increased in gp41, although no significant changes from 2014 onwards were identified in either inland or fishing communities. SDI trends were generally similar irrespective of age, sex, occupation, number of sexual partners in the past year, extra-community partners in the past year, or among incident/non-incident cases (Supplementary Fig. 9-15). Notable exceptions included adolescents and young adults (15-24 years) and persons with a recent migration history in inland communities, with these groups exhibiting slightly higher levels of p24 inter-subtype HIV diversity than the rest of the population in the most recent survey period (Supplementary Fig. 10 and 14).

**Figure 3.**
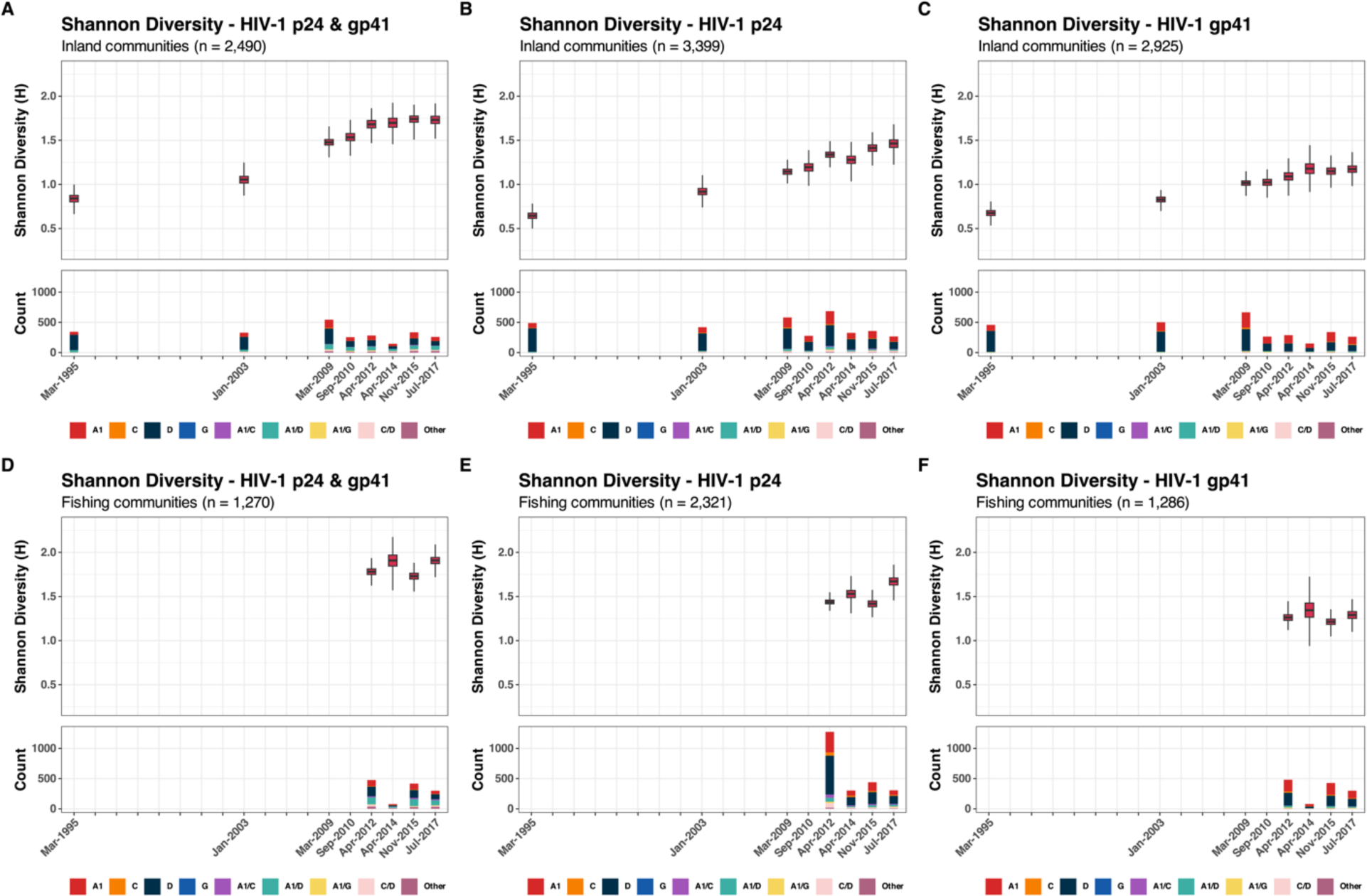
The Shannon diversity index of p24, gp41, and both p24 and gp41 in 31 inland agrarian and semi-urban trading communities and four Lake Victoria fishing communities between 1995 and 2017

#### Trends in nucleotide diversity and genetic divergence

Within inland communities, overall nucleotide diversity, irrespective of viral subtype, increased in both genes over the analysis period (Supplementary Fig. 16). However, intra-subtype diversity in A1 and D p24 subtypes leveled off after 2014 (Fig. 4). In contrast, A1 and D gp41 nucleotide diversity increased continuously. In fishing communities, p24 nucleotide diversity in subtype A1 increased through 2014 then decreased, while p24 nucleotide diversity of subtype D increased through 2015 then decreased (Fig. 4 and Supplementary Fig. 17). Similar to inland communities, overall and intra-subtype specific nucleotide gp41 diversity increased throughout the analysis periods. Nucleotide diversity trends were generally similar across population-subgroups (Supplementary Fig. 18-30), although slightly higher overall p24 nucleotide diversity, but not intra-subtype diversity, was observed among younger individuals 15-24 years in inland communities (Supplementary Fig. 20).

**Figure 4.**
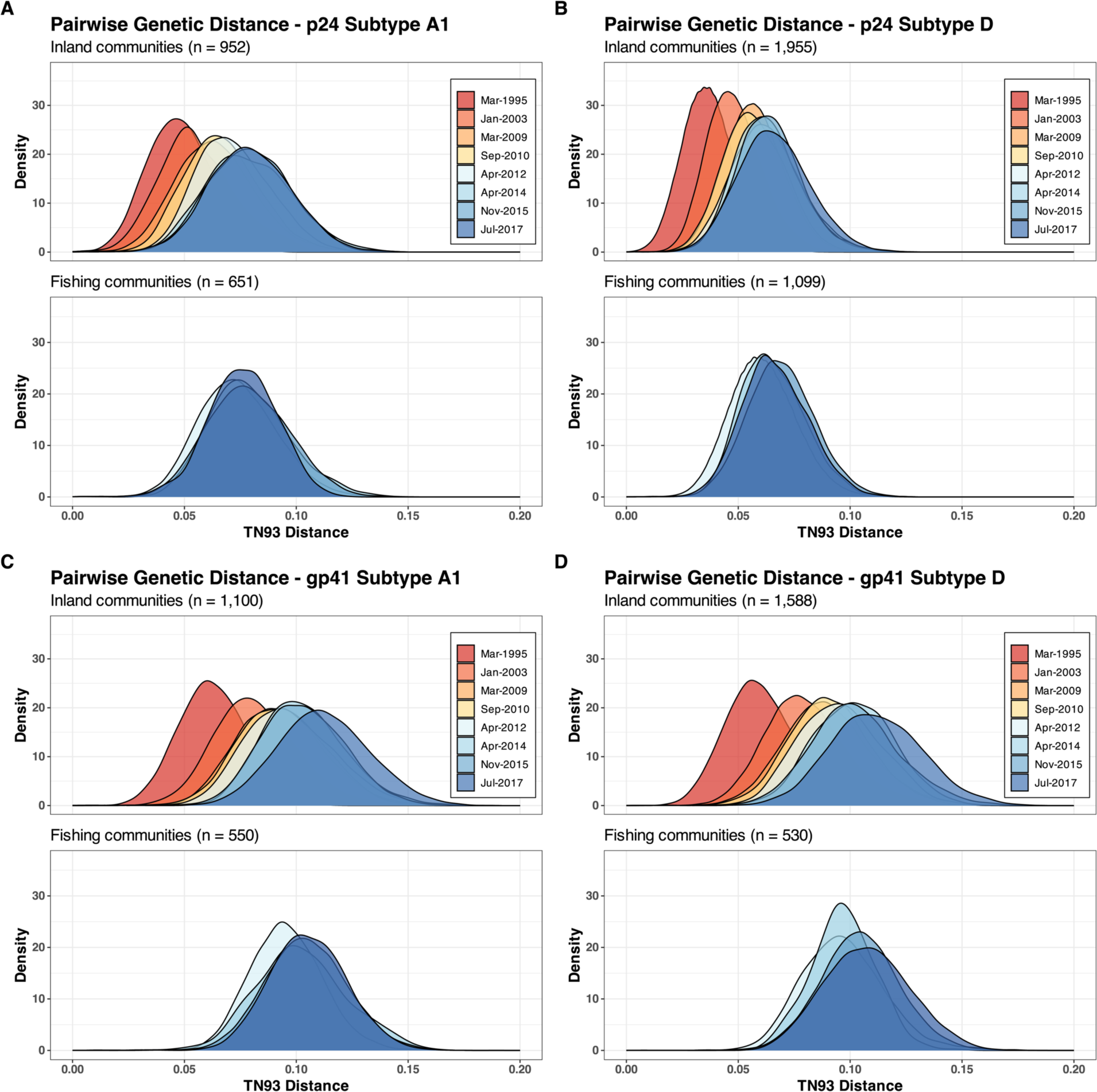
Pairwise TN93 genetic distances of p24 and gp41 subtypes A1 and D in 31 inland agrarian and semi-urban trading communities and four hyperendemic Lake Victoria fishing communities between 1995 and 2017

We further evaluated population-level HIV genetic divergence by using root-to-tip divergence analyses (Supplementary Fig. 31) and pairwise TN93 distances between subtype reference consensus sequences (A1, C, and D) and HIV sequences in Rakai (Supplementary Fig. 32). Similar to nucleotide diversity, p24 diverged through 2014 and then leveled off, while gp41 diverged over the analysis period. In the analyses of root-to-tip divergence, higher degrees of genetic variability were observed in gp41 compared to p24 in the same surveys.

#### HIV selection pressure measures

Overall, Tajima’s D values were less than 0, lower in p24 than in gp41 over the analysis period, indicating stronger purifying selection in p24 than in gp41 (Supplementary Fig. 33 and 34). In both inland and fishing communities, *ω* values were less than 1, indicating a gene-wide purifying selection acting on p24 and gp41 subtype A1 and D viruses (Supplementary Table 12). Consistent with Tajima’s D analyses, *ω* values in p24 were also lower than those in gp41. In p24, subtype A1 viruses had slightly higher *ω* than subtype D viruses, suggesting relatively weaker purifying selection pressure acting on subtype A1. In contrast, *ω* values were slightly higher among gp41 subtype D than among gp41 subtype A1, indicating relatively stronger purifying selection acting on subtype A1 gp41.

## Discussion

In this study, we examined longitudinal trends in population-level HIV diversity within 31 inland agrarian and semi-urban trading communities and four hyperendemic Lake Victoria fishing communities in Rakai, Uganda over a 25-year period including the introduction and the expansion of HIV prevention and treatment programs. While HIV incidence in eastern and southern Africa has declined by more than 50% since 2010 (Grabowski et al., 2017; Joshi et al., 2021; UNAIDS, 2023), we found that population HIV genetic diversity and divergence increased over calendar time. Increases in HIV genetic diversity were primarily driven by an increasing proportion of inter-subtype recombinant forms and rising intra-subtype gp41 genetic diversity. While Lake Victoria fishing communities are recognized as key populations with a high HIV burden and population mobility (Chang et al., 2016; Kagaayi et al., 2019), neither inter-nor intra-subtype genetic diversity was significantly higher compared to inland communities.

Overall, our HIV-1 genomic sequence data showed that trends in HIV genetic diversity were generally consistent with those from previous analyses conducted in Uganda and Eastern Africa. Notably, the proportions of inter-subtype recombinant viruses and their increasing trends, and a decline in subtype D viruses were remarkably similar to those in previous systematic reviews and molecular studies of HIV sequence data from Uganda (Grant et al., 2020; Hemelaar et al., 2020) and within Africa more broadly (Hemelaar et al., 2019; Lee et al., 2017). Here, we further show that changes in HIV subtype distribution persisted following the extensive scale-up of HIV programs; subtypes A1 and D remained predominant, while the proportion of inter-subtype recombinants increased. A previous molecular epidemiologic study from the RCCS reported a decline in HIV intra-subtype genetic diversity following the ART roll-out (Lamers et al., 2020). However, our findings from both nucleotide diversity and pairwise TN93 distance analyses indicate a consistent increase in genetic diversity in gp41, whereas diversity of subtypes A1 and D in p24 leveled off after the scale-up of HIV programs.

Between 1994 and 2018, we observed a substantial decrease in the proportion of subtype D viruses and an increasing proportion of subtype A1 viruses among individuals living in inland communities. These trends are consistent with findings in previous studies from Uganda (Conroy et al., 2010; Hemelaar et al., 2019; Lamers et al., 2020), and is likely attributed to pathogenic and clinical characteristics of subtypes A1 and D viruses. For example, infection with HIV-1 subtype D is associated with a rapid decrease in CD4+ cell count compared to subtype A1 and other non-D subtypes, resulting in faster progression to acquired immunodeficiency syndrome (AIDS) and earlier initiation of ART. Thus, there is a relatively shorter amount of time for subtype D viruses to be transmitted than subtype A1 or other subtypes (Baeten et al., 2007; Kaleebu et al., 2002; Kiwanuka et al., 2008; Ssemwanga et al., 2013). Additionally, in a previous study in Uganda, individuals in HIV serodiscordant couples whose partners were infected with subtype D viruses were found to be at a lower risk of acquiring HIV compared to those with subtype A1 (Kiwanuka et al., 2009). The higher rate of HIV disease progression and lower transmissibility of subtype D compared to subtype A1 or other subtypes may have led to the observed patterns in the proportion of subtype A1 and D viruses in this study.

We further showed that increases in HIV genetic diversity were largely driven by an increase in HIV inter-subtype recombinant infections. In both inland and fishing communities, the most common recombinants were A1/D recombinants, consistent with previous molecular and surveillance studies in Uganda (Arroyo et al., 2006; Capoferri et al., 2020; Conroy et al., 2010; Eshleman et al., 2002; Grant et al., 2020; Harris et al., 2002; Lamers et al., 2020; Yirrell et al., 2002). Interestingly, despite the low prevalence of pure subtype C viruses in our HIV sequence data, we observed a substantial increase in inter-subtype C recombinants (A1/C and C/D) in p24. This might be attributed to introduction of subtype C, which accounted for 15% of HIV infections in Eastern Africa by 2015 (Hemelaar et al., 2019). A previous study in Lake Victoria fishing communities in southern Uganda indicated a higher HIV prevalence among in-migrating populations from other geographic locations, with the highest rates observed among migrants from Tanzania (Grabowski et al., 2020). Another molecular study, conducted prior to ART roll-out in Rakai, revealed that 19% of inter-subtype recombinants were A1/C and C/D viruses in trading centers along Masaka road connecting Kampala to the border with Tanzania (Arroyo et al., 2006). These studies suggest that there might be a continuous influx of subtype C viruses or C recombinants into Uganda (Bbosa et al., 2019; Grabowski et al., 2020), resulting in high levels of inter-subtype viral mixing and increasing HIV genetic diversity.

Increasing HIV subtype diversity might be partly driven by individuals acquiring multiple infections of different HIV subtypes. Previous analyses from this same population revealed that individuals who remain viremic tended to have substantially higher levels of HIV-related risk behaviors compared to those who were virally suppressed (Brophy et al., 2021; Kennedy et al., 2023), potentially increasing the risk of coinfections and superinfections. Interestingly, however, our analyses of inter-subtype genetic diversity revealed no significant differences in levels of recombinant infection across various population sub-groups, including those with and without recent history of migration or those having extra-community sexual partners. In particular, despite high levels of HIV-related risk behaviors, population mobility in fishing communities, inter-subtype diversity was not significantly higher compared to inland communities. This might be due to widespread viral population mixing and pervasive inter-subtype recombination in the generalized and diverse HIV epidemic in Uganda (Grant et al., 2020; Lee et al., 2017). In analyses of HIV subtype distribution from individuals with HIV sequence data in both genes, the proportion of recombinant viruses in 2017 were 44.8% in inland communities and 51.7% in fishing communities. These estimates are consistent with findings in a previous study of near full-length genomes of HIV in Uganda, indicating that nearly 50% of viruses were unique recombinants forms (URFs) (Grant et al., 2020). Inter-subtype recombination via coinfections and superinfections is common in Rakai (Redd et al., 2012, 2013; J. E. Taylor & Korber, 2005), and potentially driving HIV diversity in declining epidemic in this region.

Our data showed increasing trends in intra-subtype diversity and genetic divergence in gp41 irrespective of population sub-groups. We also found higher Tajima’s D and *ω* values in gp41, consistent with the previous analyses at both individual and population-level in Rakai and elsewhere (Alizon & Fraser, 2013; Raghwani et al., 2018). These findings suggest that diversifying selection is acting more intensively on gp41 (or *env*), possibly due to its critical role in immune evasion, while p24 (or *gag*) is likely under stronger functional constraints (Lin et al., 2019). However, it is unclear why subtype-specific genetic diversity in p24 leveled off following the widespread expansion of HIV treatment and prevention programs. One possibility is that the success of HIV intervention programs might limit the opportunities for mutation and recombination events. Continued molecular surveillance with full-length HIV genome sequence data from diverse geographic locations may be needed to understand the changing dynamics of HIV and the long-term efficacy of these HIV programs.

High levels of genetic diversity in gp41 have significant implications for vaccine development. Various candidate HIV vaccines have either failed or demonstrated limited efficacy due to the HIV genetic diversity, which led to calls for developing new vaccines that elicit broadly neutralizing antibodies (bNAbs) responses (Burton, 2019; Burton & Hangartner, 2016; Sadanand et al., 2016; Steichen et al., 2019). bNAbs are capable of recognizing and neutralizing a breadth of HIV subtypes by binding to relatively conserved regions on the *env* protein. The primary challenge in developing vaccines is to reliably induce these bNAbs responses, which could be complicated by the high mutation rate and variability on the *env* gene. For example, 2F5 and 4E10 are two extensively studied bNAbs that target the membrane-proximal external region (MPER) of the *env* protein, a part of the region in our gp41 sequence analyses (Alam et al., 2007; Brunel et al., 2006; Cardoso et al., 2005; Ofek et al., 2004). Increasing genetic diversity in gp41, especially around the MPER or other potential target regions in the whole *env* protein, may pose significant challenges for vaccine design, as genetic changes through mutations or inter-subtype recombination in these regions may allow the viruses to escape bNAbs responses, potentially reducing the efficacy of vaccines. Our findings highlight the need for a multifaceted approach in HIV vaccine development that not only focuses on the robust induction of bNAbs but also considers the rapid genetic shifts in gp41 though enhanced molecular surveillance and monitoring.

There are several limitations to this study. We used two short regions of HIV for our analyses; thus, it is possible that our assessment of the HIV subtype distribution might be biased and the extent of HIV genetic diversity is likely underestimated. Previous analyses of near full-length genomes from Uganda estimated that 30-50% of viruses were inter-subtype recombinants (Grant et al., 2020; Harris et al., 2002; Lee et al., 2017). In particular, the frequency of recombination breakpoints tends to be lower in the essential regions (e.g., *gag*, *pol*, and *env*), with the *env* gene exhibiting the lowest frequency (Grant et al., 2020; Lee et al., 2017). This could explain the relatively lower proportion of inter-subtype recombinant viruses in gp41 compared to p24 in our data. Although we analyzed HIV diversity across both genes, this underestimates the proportion of recombinant viruses. Thus, the analyses of short regions do not fully capture the overall subtype distribution, and genome-wide studies would be necessary to provide a more accurate assessment of the HIV genetic diversity. Additionally, intra-subtype recombination, which may occur just as frequently as inter-subtype recombination, could significantly contribute to genetic variation within subtypes, but we did not evaluate it here. We utilized HIV sequence data generated from different sequencing methods and protocols. These changes in sequencing methods may have influenced our assessment of intra-subtype HIV genetic diversity. Young individuals and men in Rakai generally had lower participation rates in the RCCS (Grabowski et al., 2020). However, these population sub-groups were more likely to be sequenced, particularly in the later survey rounds, thus these individuals living with HIV were overrepresented in our sequence data. Lastly, we used self-reported data to evaluate the HIV-1 genetic diversity among individuals who had HIV-related high-risk behaviors, which may be influenced by reporting bias.

In conclusion, we observed continued HIV evolution, with a growing number of recombinant viruses and increasing intra-subtype evolution of gp41, despite declining HIV incidence. Overall, HIV genetic diversity increased in both inland agrarian and trading communities and hyperendemic Lake Victoria fishing communities, irrespective of population sub-groups, potentially reflecting the generalized nature of the HIV epidemic in Uganda. Comprehensive and wide-ranging molecular surveillance is essential for a better understanding of HIV dynamics driving population HIV evolution and the development of new HIV vaccines and treatments for epidemic control.

## Data Availability

Data are available upon reasonable request to the study authors.

## Acknowledgements

This study is supported by the Division of Intramural Research, National Institute of Allergy and Infectious Diseases, NIH (R01AI110324, R01AI143333-01, R01AI155080-01A1, R01MH115799-02, R01HL152813-01A1), the HIV Prevention Trials Network Laboratory Center (UM1-AI068613), and the Bill and Melinda Gates Foundation (OPP1175094, OPP1084362).

