## Supplementary information for "Increasing intra- and inter-subtype HIV diversity despite declining HIV incidence in Uganda"

**Supplementary Table 1.** GenBank accession numbers of Los Alamos National Laboratory subtype reference sequences

| Subtype | Accession number | Year of sampling | Country of origin |
|---|---|---|---|
| **A1** | U51190 | 1992 | Uganda |
| | AF004885 | 1994 | Kanya |
| | AF069670 | 1994 | Somalia |
| | AF484509 | 1998 | Uganda |
| **C** | U46016 | 1986 | Ethiopia |
| | U52953 | 1992 | Brazil |
| | AF067155 | 1995 | India |
| | AY772699 | 2004 | South Africa |
| **D** | K03454 | 1983 | DRC |
| | U88824 | 1994 | Uganda |
| | AY371157 | 2001 | Cameroon |
| | AY253311 | 2001 | Tanzania |
| **G** | AF061642 | 1993 | DRC |
| | AF061640 | 1993 | Kenya |
| | AF084936 | 1996 | DRC |
| | U88826 | 1992 | Nigeria |

**Supplementary Table 2.** The results of subtype analyses from COMET, REGA, and RIP in 31 inland agrarian and semi-urban trading communities between 1995 and 2017

| Community | Inland communities | | | | | | | |
|---|---|---|---|---|---|---|---|---|
| | p24 (n = 3,399) | | | | gp41 (n = 2,925) | | | |
| Subtype | COMET | REGA | COMET & REGA | RIP | COMET | REGA | COMET & REGA | RIP |
| A1 | 1,052 | 1,000 | 1,099 | 952 | 673 | 799 | 936 | 1,100 |
| B | 19 | 0 | 19 | 0 | 1 | 0 | 1 | 0 |
| C | 105 | 95 | 108 | 75 | 81 | 76 | 84 | 65 |
| D | 1,973 | 0 | 1,973 | 1,955 | 1,165 | 648 | 1,333 | 1,588 |
| G | 5 | 5 | 5 | 5 | 1 | 0 | 1 | 2 |
| 01_AE | 1 | 0 | 1 | 0 | 0 | 0 | 0 | 0 |
| 02_AG | 1 | 0 | 1 | 0 | 1 | 0 | 1 | 0 |
| A1/C | 0 | 0 | 0 | 52 | 0 | 0 | 0 | 14 |
| A1/D | 0 | 0 | 0 | 170 | 0 | 0 | 0 | 67 |
| A1/G | 0 | 0 | 0 | 41 | 0 | 0 | 0 | 62 |
| C/D | 0 | 0 | 0 | 114 | 0 | 0 | 0 | 11 |
| Other | 0 | 0 | 0 | 35 | 0 | 0 | 0 | 16 |
| Not assigned | 243 | 2,299 | 193 | 0 | 1,003 | 1,402 | 569 | 0 |

**Supplementary Table 3.** The results of subtype analyses from COMET, REGA, and RIP in four hyperendemic Lake Victoria communities between 2012 and 2017

| Community | Fishing communities | | | | | | | |
|---|---|---|---|---|---|---|---|---|
| | p24 (n = 2,321) | | | | gp41 (n = 1,286) | | | |
| Subtype | COMET | REGA | COMET & REGA | RIP | COMET | REGA | COMET & REGA | RIP |
| A1 | 771 | 721 | 829 | 651 | 301 | 346 | 452 | 550 |
| B | 10 | 0 | 10 | 0 | 0 | 0 | 0 | 0 |
| C | 159 | 144 | 163 | 111 | 68 | 54 | 71 | 54 |
| D | 1,134 | 0 | 1,135 | 1,099 | 464 | 178 | 487 | 530 |
| G | 11 | 9 | 11 | 11 | 7 | 0 | 7 | 7 |
| 01_AE | 1 | 0 | 1 | 0 | 0 | 0 | 0 | 0 |
| 02_AG | 0 | 0 | 0 | 0 | 7 | 0 | 7 | 0 |
| A1/C | 0 | 0 | 0 | 102 | 0 | 0 | 0 | 11 |
| A1/D | 0 | 0 | 0 | 146 | 0 | 0 | 0 | 72 |
| A1/G | 0 | 0 | 0 | 41 | 0 | 0 | 0 | 44 |
| C/D | 0 | 0 | 0 | 128 | 0 | 0 | 0 | 7 |
| Other | 2 | 0 | 2 | 32 | 0 | 0 | 0 | 11 |
| Not assigned | 223 | 1,447 | 171 | 0 | 439 | 708 | 262 | 0 |

**Supplementary Table 4.** Demographic and HIV-related risk behavioral characteristics of RCCS participants living with HIV, with and without HIV sequence data in 31 inland agrarian and semi-urban trading communities between 1995 and 2017

| Inland communities | R1 (Nov-1994 – Aug-1995) | | | R9 (July-2002 – Aug-2003) | | | R13 (June-2008 – Dec-2009) | | | R14 (Jan-2010 – June-2011) | | |
|---|---|---|---|---|---|---|---|---|---|---|---|---|
| | PLHIV | PLHIV w/ seq | PLHIV wo/ seq | PLHIV | PLHIV w/ seq | PLHIV wo/ seq | PLHIV | PLHIV w/ seq | PLHIV wo/ seq | PLHIV | PLHIV w/ seq | PLHIV wo/ seq |
| **N** | 1,162 | 604 | 558 | 988 | 591 | 397 | 1,218 | 703 | 515 | 1,365 | 287 | 1,078 |
| **Age** | | | | | | | | | | | | |
| Median age (IQR) | 29 (24, 35) | 30 (24, 35) | 28 (24, 35) | 31 (26, 37) | 31 (26, 37) | 31 (27, 37) | 33 (28, 39) | 31 (26, 37) | 35 (31, 41) | 33 (28, 39) | 30 (25, 36) | 34 (28, 40) |
| 15 – 24 | 315 (27.1%) | 154 (25.5%) | 161 (28.9%) | 190 (19.2%) | 113 (19.1%) | 77 (19.4%) | 170 (14%) | 130 (18.5%) | 40 (7.8%) | 184 (13.5%) | 66 (23%) | 118 (10.9%) |
| 25 – 34 | 531 (45.7%) | 279 (46.2%) | 252 (45.2%) | 454 (46%) | 275 (46.5%) | 179 (45.1%) | 542 (44.5%) | 346 (49.2%) | 196 (38.1%) | 572 (41.9%) | 127 (44.3%) | 445 (41.3%) |
| 35 + | 316 (27.2%) | 171 (28.3%) | 145 (26%) | 344 (34.8%) | 203 (34.3%) | 141 (35.5%) | 506 (41.5%) | 227 (32.3%) | 279 (54.2%) | 609 (44.6%) | 94 (32.8%) | 515 (47.8%) |
| **Sex** | | | | | | | | | | | | |
| Male | 440 (37.9%) | 251 (41.6%) | 189 (33.9%) | 355 (35.9%) | 208 (35.2%) | 147 (37%) | 425 (34.9%) | 263 (37.4%) | 162 (31.5%) | 469 (34.4%) | 115 (40.1%) | 354 (32.8%) |
| Female | 722 (62.1%) | 353 (58.4%) | 369 (66.1%) | 633 (64.1%) | 383 (64.8%) | 250 (63%) | 793 (65.1%) | 440 (62.6%) | 353 (68.5%) | 896 (65.6%) | 172 (59.9%) | 724 (67.2%) |
| **Occupation** | | | | | | | | | | | | |
| Agriculture | 654 (56.3%) | 331 (54.8%) | 323 (57.9%) | 550 (55.7%) | 332 (56.2%) | 218 (54.9%) | 637 (52.3%) | 359 (51.1%) | 278 (54%) | 735 (53.8%) | 142 (49.5%) | 593 (55%) |
| Trade/Truck | 150 (12.9%) | 90 (14.9%) | 60 (10.8%) | 98 (9.9%) | 56 (9.5%) | 42 (10.6%) | 168 (13.8%) | 94 (13.4%) | 74 (14.4%) | 166 (12.2%) | 37 (12.9%) | 129 (12%) |
| Restaurant/Bar | 32 (2.8%) | 18 (3%) | 14 (2.5%) | 34 (3.4%) | 27 (4.6%) | 7 (1.8%) | 58 (4.8%) | 32 (4.6%) | 26 (5%) | 71 (5.2%) | 15 (5.2%) | 56 (5.2%) |
| Other | 326 (28.1%) | 165 (27.3%) | 161 (28.9%) | 306 (31%) | 176 (29.8%) | 130 (32.7%) | 355 (29.1%) | 218 (31%) | 137 (26.6%) | 393 (28.8%) | 93 (32.4%) | 300 (27.8%) |
| **Number of sexual partners in the past year** | | | | | | | | | | | | |
| 0 – 1 | 959 (82.5%) | 488 (80.8%) | 471 (84.4%) | 635 (64.3%) | 379 (64.1%) | 256 (64.5%) | 794 (65.2%) | 478 (68%) | 316 (61.4%) | 911 (66.7%) | 180 (62.7%) | 731 (67.8%) |
| 2 + | 197 (17%) | 114 (18.9%) | 83 (14.9%) | 199 (20.1%) | 123 (20.8%) | 76 (19.1%) | 239 (19.6%) | 149 (21.2%) | 90 (17.5%) | 262 (19.2%) | 75 (26.1%) | 187 (17.3%) |
| No response | 6 (0.5%) | 2 (0.3%) | 4 (0.7%) | 154 (15.6%) | 89 (15.1%) | 65 (16.4%) | 185 (15.2%) | 76 (10.8%) | 109 (21.2%) | 192 (14.1%) | 32 (11.1%) | 160 (14.8%) |
| **Any sexual partner outside the community in the past year** | | | | | | | | | | | | |
| Yes | 193 (16.6%) | 101 (16.7%) | 92 (16.5%) | 271 (27.4%) | 170 (28.8%) | 101 (25.4%) | 333 (27.3%) | 202 (28.7%) | 131 (25.4%) | 424 (31.1%) | 96 (33.4%) | 328 (30.4%) |
| No | 968 (83.3%) | 502 (83.1%) | 466 (83.5%) | 563 (57%) | 332 (56.2%) | 231 (58.2%) | 700 (57.5%) | 425 (60.5%) | 275 (53.4%) | 747 (54.7%) | 159 (55.4%) | 588 (54.5%) |
| No response | 1 (0.1%) | 1 (0.2%) | 0 (0%) | 154 (15.6%) | 89 (15.1%) | 65 (16.4%) | 185 (15.2%) | 76 (10.8%) | 109 (21.2%) | 194 (14.2%) | 32 (11.1%) | 162 (15%) |
| **Recent history of migration** | | | | | | | | | | | | |
| Yes | 0 (0%) | 0 (0%) | 0 (0%) | 136 (13.8%) | 99 (16.8%) | 37 (9.3%) | 171 (14%) | 113 (16.1%) | 58 (11.3%) | 258 (18.9%) | 69 (24%) | 189 (17.5%) |
| No | 1,115 (96%) | 578 (95.7%) | 537 (96.2%) | 852 (86.2%) | 492 (83.2%) | 360 (90.7%) | 1,045 (85.8%) | 588 (83.6%) | 457 (88.7%) | 1,107 (81.1%) | 218 (76%) | 889 (82.5%) |
| Non-resident | 47 (4%) | 26 (4.3%) | 21 (3.8%) | 0 (0%) | 0 (0%) | 0 (0%) | 2 (0.2%) | 2 (0.3%) | 0 (0%) | 0 (0%) | 0 (0%) | 0 (0%) |
| **Self-reported ART use** | | | | | | | | | | | | |
| Yes | - | - | - | - | - | - | 268 (22%) | 9 (1.3%) | 259 (50.3%) | 365 (26.7%) | 0 (0%) | 365 (33.9%) |
| No | - | - | - | - | - | - | 731 (60%) | 524 (74.5%) | 207 (40.2%) | 768 (56.3%) | 216 (75.3%) | 552 (51.2%) |
| No response | - | - | - | - | - | - | 219 (18%) | 170 (24.2%) | 49 (9.5%) | 232 (17%) | 71 (24.7%) | 161 (14.9%) |
| **Viral load** | | | | | | | | | | | | |
| ≥1,000 | 462 (39.8%) | 277 (45.9%) | 185 (33.2%) | 81 (8.2%) | 62 (10.5%) | 19 (4.8%) | 690 (56.7%) | 573 (81.5%) | 117 (22.7%) | 65 (4.8%) | 21 (7.3%) | 44 (4.1%) |
| 50≤VL<1,000 | 24 (2.1%) | 11 (1.8%) | 13 (2.3%) | 3 (0.3%) | 1 (0.2%) | 2 (0.5%) | 117 (9.6%) | 71 (10.1%) | 46 (8.9%) | 3 (0.2%) | 0 (0%) | 3 (0.3%) |
| BD | 21 (1.8%) | 2 (0.3%) | 19 (3.4%) | 12 (1.2%) | 7 (1.2%) | 5 (1.3%) | 355 (29.1%) | 29 (4.1%) | 326 (63.3%) | 8 (0.6%) | 0 (0%) | 8 (0.7%) |
| No data | 655 (56.4%) | 314 (52%) | 341 (61.1%) | 892 (90.3%) | 521 (88.2%) | 371 (93.5%) | 56 (4.6%) | 30 (4.3%) | 26 (5%) | 1,289 (94.4%) | 266 (92.7%) | 1,023 (94.9%) |

**Supplementary Table 4.** Demographic and HIV-related risk behavioral characteristics of RCCS participants living with HIV, with and without HIV sequence data in 31 inland agrarian and semi-urban trading communities between 1995 and 2017 (Cont.)

| Inland communities | R15 (Aug-2011 –Apr-2013) | | | R16 (July-2013 – Jan-2015) | | | R17 (Feb-2015 – Oct-2016) | | | R18 (Oct-2016 – May-2018) | | |
|---|---|---|---|---|---|---|---|---|---|---|---|---|
| | PLHIV | PLHIV w/ seq | PLHIV wo/ seq | PLHIV | PLHIV w/ seq | PLHIV wo/ seq | PLHIV | PLHIV w/ seq | PLHIV wo/ seq | PLHIV | PLHIV w/ seq | PLHIV wo/ seq |
| **N** | 1,474 | 691 | 783 | 1,630 | 331 | 1,299 | 1,720 | 360 | 1,360 | 1,648 | 269 | 1,379 |
| **Age** | | | | | | | | | | | | |
| Median age (IQR) | 33 (27, 39) | 31 (25, 36) | 36 (30, 41) | 34 (28, 40) | 30 (25, 37) | 35 (29, 41) | 35 (28, 41) | 31 (25, 37) | 36 (29, 41) | 36 (30, 42) | 30 (25, 35) | 37 (31, 42) |
| 15 – 24 | 215 (14.6%) | 143 (20.7%) | 72 (9.2%) | 220 (13.5%) | 67 (20.2%) | 153 (11.8%) | 219 (12.7%) | 82 (22.8%) | 137 (10.1%) | 173 (10.5%) | 66 (24.5%) | 107 (7.8%) |
| 25 – 34 | 601 (40.8%) | 325 (47%) | 276 (35.2%) | 603 (37%) | 157 (47.4%) | 446 (34.3%) | 602 (35%) | 149 (41.4%) | 453 (33.3%) | 559 (33.9%) | 130 (48.3%) | 429 (31.1%) |
| 35 + | 658 (44.6%) | 223 (32.3%) | 435 (55.6%) | 807 (49.5%) | 107 (32.3%) | 700 (53.9%) | 899 (52.3%) | 129 (35.8%) | 770 (56.6%) | 916 (55.6%) | 73 (27.1%) | 843 (61.1%) |
| **Sex** | | | | | | | | | | | | |
| Male | 503 (34.1%) | 286 (41.4%) | 217 (27.7%) | 536 (32.9%) | 138 (41.7%) | 398 (30.6%) | 536 (31.2%) | 150 (41.7%) | 386 (28.4%) | 507 (30.8%) | 116 (43.1%) | 391 (28.4%) |
| Female | 971 (65.9%) | 405 (58.6%) | 566 (72.3%) | 1,094 (67.1%) | 193 (58.3%) | 901 (69.4%) | 1,184 (68.8%) | 210 (58.3%) | 974 (71.6%) | 1,141 (69.2%) | 153 (56.9%) | 988 (71.6%) |
| **Occupation** | | | | | | | | | | | | |
| Agriculture | 739 (50.1%) | 322 (46.6%) | 417 (53.3%) | 503 (30.9%) | 119 (36%) | 384 (29.6%) | 853 (49.6%) | 154 (42.8%) | 699 (51.4%) | 870 (52.8%) | 115 (42.8%) | 755 (54.7%) |
| Trade/Truck | 207 (14%) | 98 (14.2%) | 109 (13.9%) | 122 (7.5%) | 45 (13.6%) | 77 (5.9%) | 230 (13.4%) | 63 (17.5%) | 167 (12.3%) | 216 (13.1%) | 52 (19.3%) | 164 (11.9%) |
| Restaurant/Bar | 109 (7.4%) | 55 (8%) | 54 (6.9%) | 64 (3.9%) | 20 (6%) | 44 (3.4%) | 112 (6.5%) | 16 (4.4%) | 96 (7.1%) | 103 (6.2%) | 10 (3.7%) | 93 (6.7%) |
| Other | 419 (28.4%) | 216 (31.3%) | 203 (25.9%) | 941 (57.7%) | 147 (44.4%) | 794 (61.1%) | 525 (30.5%) | 127 (35.3%) | 398 (29.3%) | 459 (27.9%) | 92 (34.2%) | 367 (26.6%) |
| **Number of sexual partners in the past year** | | | | | | | | | | | | |
| 0 – 1 | 945 (64.1%) | 438 (63.4%) | 507 (64.8%) | 1,043 (64%) | 204 (61.6%) | 839 (64.6%) | 1,121 (65.2%) | 231 (64.2%) | 890 (65.4%) | 1,020 (61.9%) | 149 (55.4%) | 871 (63.2%) |
| 2 + | 309 (21%) | 189 (27.4%) | 120 (15.3%) | 316 (19.4%) | 93 (28.1%) | 223 (17.2%) | 359 (20.9%) | 92 (25.6%) | 267 (19.6%) | 346 (21%) | 82 (30.5%) | 264 (19.1%) |
| No response | 220 (14.9%) | 64 (9.3%) | 156 (19.9%) | 271 (16.6%) | 34 (10.3%) | 237 (18.2%) | 240 (14%) | 37 (10.3%) | 203 (14.9%) | 282 (17.1%) | 38 (14.1%) | 244 (17.7%) |
| **Any sexual partner outside the community in the past year** | | | | | | | | | | | | |
| Yes | 448 (30.4%) | 235 (34%) | 213 (27.2%) | 300 (18.4%) | 92 (27.8%) | 208 (16%) | 519 (30.2%) | 120 (33.3%) | 399 (29.3%) | 482 (29.2%) | 101 (37.5%) | 381 (27.6%) |
| No | 801 (54.3%) | 388 (56.2%) | 413 (52.7%) | 569 (34.9%) | 164 (49.5%) | 405 (31.2%) | 961 (55.9%) | 203 (56.4%) | 758 (55.7%) | 884 (53.6%) | 130 (48.3%) | 754 (54.7%) |
| No response | 225 (15.3%) | 68 (9.8%) | 157 (20.1%) | 761 (46.7%) | 75 (22.7%) | 686 (52.8%) | 240 (14%) | 37 (10.3%) | 203 (14.9%) | 282 (17.1%) | 38 (14.1%) | 244 (17.7%) |
| **Recent history of migration** | | | | | | | | | | | | |
| Yes | 381 (25.8%) | 223 (32.3%) | 158 (20.2%) | 464 (28.5%) | 144 (43.5%) | 320 (24.6%) | 402 (23.4%) | 116 (32.2%) | 286 (21%) | 405 (24.6%) | 124 (46.1%) | 281 (20.4%) |
| No | 1,093 (74.2%) | 468 (67.7%) | 625 (79.8%) | 1,163 (71.3%) | 184 (55.6%) | 979 (75.4%) | 1,317 (76.6%) | 244 (67.8%) | 1,073 (78.9%) | 1,243 (75.4%) | 145 (53.9%) | 1,098 (79.6%) |
| Non-resident | 0 (0%) | 0 (0%) | 0 (0%) | 3 (0.2%) | 3 (0.9%) | 0 (0%) | 1 (0.1%) | 0 (0%) | 1 (0.1%) | 0 (0%) | 0 (0%) | 0 (0%) |
| **Self-reported ART use** | | | | | | | | | | | | |
| Yes | 496 (33.6%) | 0 (0%) | 496 (63.3%) | 877 (53.8%) | 0 (0%) | 877 (67.5%) | 1,181 (68.7%) | 60 (16.7%) | 1,121 (82.4%) | 1,277 (77.5%) | 64 (23.8%) | 1,213 (88%) |
| No | 777 (52.7%) | 544 (78.7%) | 233 (29.8%) | 630 (38.7%) | 278 (84%) | 352 (27.1%) | 487 (28.3%) | 271 (75.3%) | 216 (15.9%) | 332 (20.1%) | 186 (69.1%) | 146 (10.6%) |
| No response | 201 (13.6%) | 147 (21.3%) | 54 (6.9%) | 123 (7.5%) | 53 (16%) | 70 (5.4%) | 52 (3%) | 29 (8.1%) | 23 (1.7%) | 39 (2.4%) | 19 (7.1%) | 20 (1.5%) |
| **Viral load** | | | | | | | | | | | | |
| ≥1,000 | 149 (10.1%) | 119 (17.2%) | 30 (3.8%) | 593 (36.4%) | 311 (94%) | 282 (21.7%) | 419 (24.4%) | 358 (99.4%) | 61 (4.5%) | 312 (18.9%) | 269 (100%) | 43 (3.1%) |
| 50≤VL<1,000 | 20 (1.4%) | 6 (0.9%) | 14 (1.8%) | 89 (5.5%) | 10 (3%) | 79 (6.1%) | 77 (4.5%) | 2 (0.6%) | 75 (5.5%) | 56 (3.4%) | 0 (0%) | 56 (4.1%) |
| BD | 112 (7.6%) | 7 (1%) | 105 (13.4%) | 927 (56.9%) | 7 (2.1%) | 920 (70.8%) | 1,212 (70.5%) | 0 (0%) | 1,212 (89.1%) | 1,264 (76.7%) | 0 (0%) | 1,264 (91.7%) |
| No data | 1,193 (80.9%) | 559 (80.9%) | 634 (81%) | 21 (1.3%) | 3 (0.9%) | 18 (1.4%) | 12 (0.7%) | 0 (0%) | 12 (0.9%) | 16 (1%) | 0 (0%) | 16 (1.2%) |

**Supplementary Table 5.** Demographic and HIV-related risk behavioral characteristics of people living with HIV, with and without HIV sequence data in four hyperendemic Lake Victoria fishing communities between 2012 and 2017

| Fishing communities | R15 (Aug-2011 –Apr-2013) | | | R16 (July-2013 – Jan-2015) | | | R17 (Feb-2015 – Oct-2016) | | | R18 (Oct-2016 – May-2018) | | |
|---|---|---|---|---|---|---|---|---|---|---|---|---|
| | PLHIV | PLHIV w/ seq | PLHIV wo/ seq | PLHIV | PLHIV w/ seq | PLHIV wo/ seq | PLHIV | PLHIV w/ seq | PLHIV wo/ seq | PLHIV | PLHIV w/ seq | PLHIV wo/ seq |
| **N** | 3,174 | 1,278 | 1,896 | 1,526 | 304 | 1,222 | 1,603 | 449 | 1,154 | 1,747 | 306 | 1,441 |
| **Age** | | | | | | | | | | | | |
| Median age (IQR) | 31 (26, 36) | 30 (25, 35) | 32 (27, 37) | 32 (27, 37) | 28 (24, 33) | 33 (28, 38) | 32 (28, 38) | 31 (26, 36) | 33 (28, 38) | 34 (28, 39) | 30 (26, 36) | 34 (29, 40) |
| 15 – 24 | 511 (16.1%) | 250 (19.6%) | 261 (13.8%) | 244 (16%) | 96 (31.6%) | 148 (12.1%) | 212 (13.2%) | 84 (18.7%) | 128 (11.1%) | 168 (9.6%) | 51 (16.7%) | 117 (8.1%) |
| 25 – 34 | 1,672 (52.7%) | 693 (54.2%) | 979 (51.6%) | 751 (49.2%) | 147 (48.4%) | 604 (49.4%) | 741 (46.2%) | 224 (49.9%) | 517 (44.8%) | 792 (45.3%) | 163 (53.3%) | 629 (43.7%) |
| 35 + | 991 (31.2%) | 335 (26.2%) | 656 (34.6%) | 531 (34.8%) | 61 (20.1%) | 470 (38.5%) | 650 (40.5%) | 141 (31.4%) | 509 (44.1%) | 787 (45%) | 92 (30.1%) | 695 (48.2%) |
| **Sex** | | | | | | | | | | | | |
| Male | 1,416 (44.6%) | 647 (50.6%) | 769 (40.6%) | 668 (43.8%) | 143 (47%) | 525 (43%) | 696 (43.4%) | 257 (57.2%) | 439 (38%) | 801 (45.9%) | 178 (58.2%) | 623 (43.2%) |
| Female | 1,758 (55.4%) | 631 (49.4%) | 1,127 (59.4%) | 858 (56.2%) | 161 (53%) | 697 (57%) | 907 (56.6%) | 192 (42.8%) | 715 (62%) | 946 (54.1%) | 128 (41.8%) | 818 (56.8%) |
| **Occupation** | | | | | | | | | | | | |
| Agriculture | 509 (16%) | 202 (15.8%) | 307 (16.2%) | 254 (16.6%) | 54 (17.8%) | 200 (16.4%) | 290 (18.1%) | 77 (17.1%) | 213 (18.5%) | 319 (18.3%) | 52 (17%) | 267 (18.5%) |
| Trade/Truck | 641 (20.2%) | 226 (17.7%) | 415 (21.9%) | 299 (19.6%) | 45 (14.8%) | 254 (20.8%) | 304 (19%) | 66 (14.7%) | 238 (20.6%) | 324 (18.5%) | 46 (15%) | 278 (19.3%) |
| Restaurant/Bar | 390 (12.3%) | 133 (10.4%) | 257 (13.6%) | 185 (12.1%) | 33 (10.9%) | 152 (12.4%) | 195 (12.2%) | 42 (9.4%) | 153 (13.3%) | 193 (11%) | 33 (10.8%) | 160 (11.1%) |
| Other | 998 (31.4%) | 468 (36.6%) | 530 (28%) | 480 (31.5%) | 108 (35.5%) | 372 (30.4%) | 508 (31.7%) | 189 (42.1%) | 319 (27.6%) | 589 (33.7%) | 135 (44.1%) | 454 (31.5%) |
| **Number of sexual partners in the past year** | | | | | | | | | | | | |
| 0 – 1 | 1,700 (53.6%) | 625 (48.9%) | 1,075 (56.7%) | 833 (54.6%) | 139 (45.7%) | 694 (56.8%) | 873 (54.5%) | 198 (44.1%) | 675 (58.5%) | 952 (54.5%) | 142 (46.4%) | 810 (56.2%) |
| 2 + | 1,326 (41.8%) | 614 (48%) | 712 (37.6%) | 598 (39.2%) | 154 (50.7%) | 444 (36.3%) | 638 (39.8%) | 223 (49.7%) | 415 (36%) | 677 (38.8%) | 148 (48.4%) | 529 (36.7%) |
| No response | 148 (4.7%) | 39 (3.1%) | 109 (5.7%) | 95 (6.2%) | 11 (3.6%) | 84 (6.9%) | 92 (5.7%) | 28 (6.2%) | 64 (5.5%) | 118 (6.8%) | 16 (5.2%) | 102 (7.1%) |
| **Any sexual partner outside the community in the past year** | | | | | | | | | | | | |
| Yes | 913 (28.8%) | 412 (32.2%) | 501 (26.4%) | 393 (25.8%) | 84 (27.6%) | 309 (25.3%) | 412 (25.7%) | 125 (27.8%) | 287 (24.9%) | 501 (28.7%) | 110 (35.9%) | 391 (27.1%) |
| No | 2,106 (66.4%) | 823 (64.4%) | 1,283 (67.7%) | 1,038 (68%) | 209 (68.8%) | 829 (67.8%) | 1,099 (68.6%) | 296 (65.9%) | 803 (69.6%) | 1,128 (64.6%) | 180 (58.8%) | 948 (65.8%) |
| No response | 155 (4.9%) | 43 (3.4%) | 112 (5.9%) | 95 (6.2%) | 11 (3.6%) | 84 (6.9%) | 92 (5.7%) | 28 (6.2%) | 64 (5.5%) | 118 (6.8%) | 16 (5.2%) | 102 (7.1%) |
| **Recent history of migration** | | | | | | | | | | | | |
| Yes | 448 (14.1%) | 260 (20.3%) | 188 (9.9%) | 292 (19.1%) | 135 (44.4%) | 157 (12.8%) | 410 (25.6%) | 149 (33.2%) | 261 (22.6%) | 395 (22.6%) | 100 (32.7%) | 295 (20.5%) |
| No | 2,723 (85.8%) | 1,017 (79.6%) | 1,706 (90%) | 1,234 (80.9%) | 169 (55.6%) | 1,065 (87.2%) | 1,193 (74.4%) | 300 (66.8%) | 893 (77.4%) | 1,351 (77.3%) | 205 (67%) | 1,146 (79.5%) |
| Non-resident | 3 (0.1%) | 1 (0.1%) | 2 (0.1%) | 0 (0%) | 0 (0%) | 0 (0%) | 0 (0%) | 0 (0%) | 0 (0%) | 1 (0.1%) | 1 (0.3%) | 0 (0%) |
| **Self-reported ART use** | | | | | | | | | | | | |
| Yes | 645 (20.3%) | 28 (2.2%) | 617 (32.5%) | 647 (42.4%) | 0 (0%) | 647 (52.9%) | 1,039 (64.8%) | 74 (16.5%) | 965 (83.6%) | 1,427 (81.7%) | 101 (33%) | 1,326 (92%) |
| No | 1,789 (56.4%) | 721 (56.4%) | 1,068 (56.3%) | 804 (52.7%) | 253 (83.2%) | 551 (45.1%) | 507 (31.6%) | 340 (75.7%) | 167 (14.5%) | 294 (16.8%) | 189 (61.8%) | 105 (7.3%) |
| No response | 740 (23.3%) | 529 (41.4%) | 211 (11.1%) | 75 (4.9%) | 51 (16.8%) | 24 (2%) | 57 (3.6%) | 35 (7.8%) | 22 (1.9%) | 26 (1.5%) | 16 (5.2%) | 10 (0.7%) |
| **Viral load** | | | | | | | | | | | | |
| ≥1,000 | 1,076 (33.9%) | 897 (70.2%) | 179 (9.4%) | 765 (50.1%) | 289 (95.1%) | 476 (39%) | 511 (31.9%) | 443 (98.7%) | 68 (5.9%) | 354 (20.3%) | 306 (100%) | 48 (3.3%) |
| 50≤VL<1,000 | 94 (3%) | 40 (3.1%) | 54 (2.8%) | 88 (5.8%) | 10 (3.3%) | 78 (6.4%) | 61 (3.8%) | 5 (1.1%) | 56 (4.9%) | 47 (2.7%) | 0 (0%) | 47 (3.3%) |
| BD | 450 (14.2%) | 21 (1.6%) | 429 (22.6%) | 666 (43.6%) | 5 (1.6%) | 661 (54.1%) | 1,024 (63.9%) | 1 (0.2%) | 1,023 (88.6%) | 1,337 (76.5%) | 0 (0%) | 1,337 (92.8%) |
| No data | 1,554 (49%) | 320 (25%) | 1,234 (65.1%) | 7 (0.5%) | 0 (0%) | 7 (0.6%) | 6 (0.4%) | 0 (0%) | 6 (0.5%) | 9 (0.5%) | 0 (0%) | 9 (0.6%) |

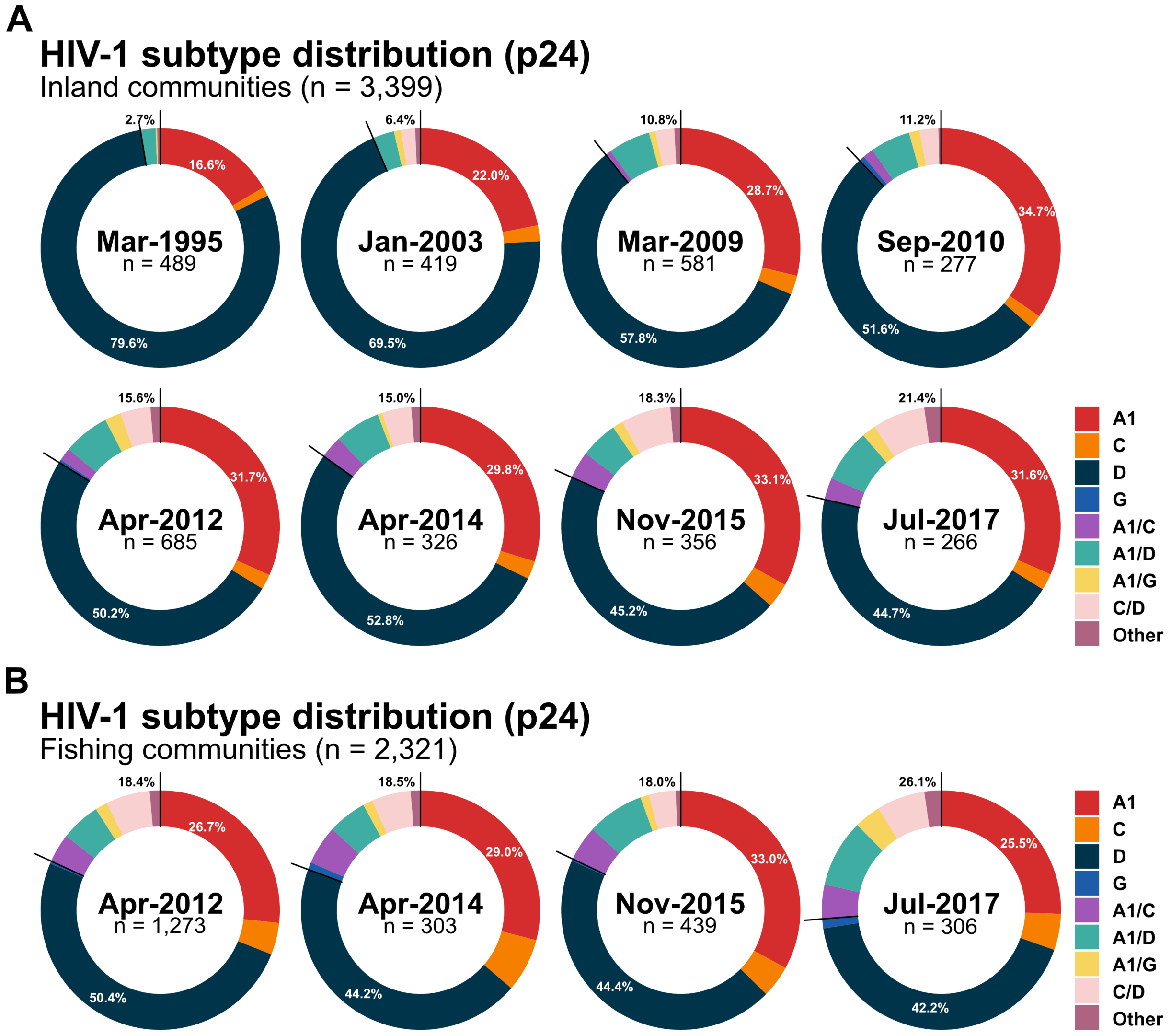


**Supplementary Figure 1.** HIV subtype distribution of p24 in 31 inland agrarian and semi-urban trading communities (Panel A) and four hyperendemic Lake Victoria fishing communities (Panel B) by calendar time

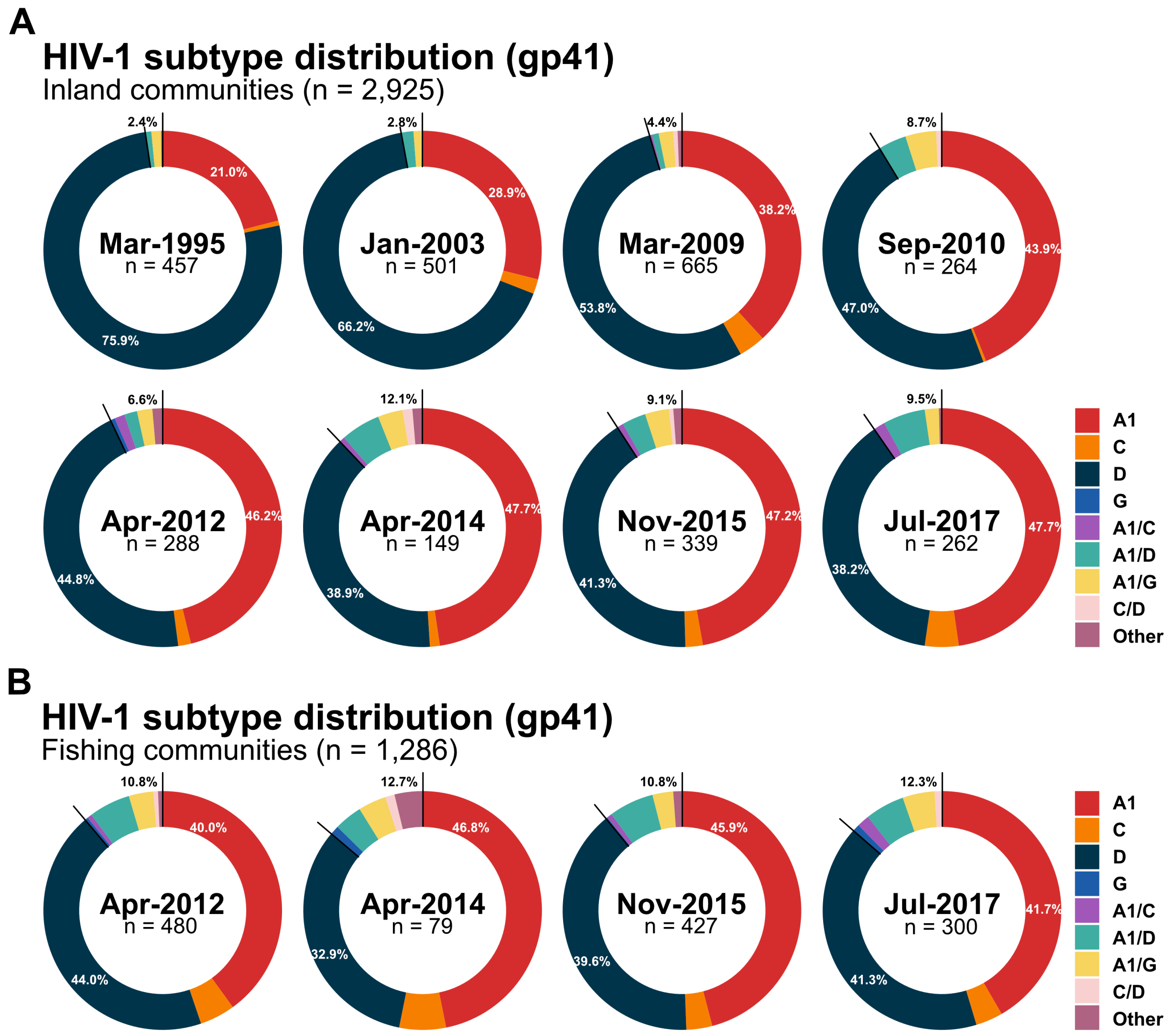


**Supplementary Figure 2.** HIV subtype distribution of gp41 in 31 inland agrarian and semi-urban trading communities (Panel A) and four hyperendemic Lake Victoria fishing communities (Panel B) by calendar time

**Supplementary Table 6.** HIV subtype distribution of p24 and gp41 from RCCS participants living with HIV who had available sequence data from both genes in the same survey rounds in 31 inland agrarian and semi-urban trading communities between 1995 and 2017

| Round | Midpoint | N | Pure subtypes | | | | | Recombinants | | | | | |
|---|---|---|---|---|---|---|---|---|---|---|---|---|---|
| | | | Total | A1 | C | D | G | Total | A1/C | A1/D | A1/G | C/D | Other |
| **Total** | - | 2,490 | 1,754 (70.4%) | 1,100 (44.2%) | 29 (1.2%) | 1,132 (45.5%) | 1 (0.1%) | 736 (29.6%) | 35 (1.4%) | 442 (17.8%) | 52 (2.1%) | 91 (3.7%) | 116 (4.7%) |
| **R001** | Mar-1995 | 343 | 301 (87.8%) | 49 (14.3%) | 1 (0.3%) | 251 (73.2%) | 0 (0%) | 42 (12.2%) | 1 (0.3%) | 36 (10.5%) | 3 (0.9%) | 0 (0%) | 2 (0.6%) |
| **R009** | Jan-2003 | 329 | 281 (85.4%) | 68 (20.7%) | 3 (0.9%) | 210 (63.8%) | 0 (0%) | 48 (14.6%) | 0 (0%) | 32 (9.7%) | 3 (0.9%) | 12 (3.6%) | 1 (0.3%) |
| **R013** | Mar-2009 | 544 | 403 (74.1%) | 136 (25.0%) | 11 (2.0%) | 256 (47.1%) | 0 (0%) | 141 (25.9%) | 5 (0.9%) | 84 (15.4%) | 9 (1.7%) | 21 (3.9%) | 22 (4.0%) |
| **R014** | Sep-2010 | 254 | 161 (63.4%) | 64 (25.2%) | 1 (0.4%) | 96 (37.8%) | 0 (0%) | 93 (36.6%) | 4 (1.6%) | 61 (24.0%) | 11 (4.3%) | 5 (2.0%) | 12 (4.7%) |
| **R015** | Apr-2012 | 282 | 176 (62.4%) | 74 (26.2%) | 4 (1.4%) | 97 (34.4%) | 1 (0.4%) | 106 (37.6%) | 6 (2.1%) | 64 (22.7%) | 11 (3.9%) | 10 (3.5%) | 15 (5.3%) |
| **R016** | Apr-2014 | 144 | 82 (56.9%) | 37 (25.7%) | 2 (1.4%) | 43 (29.9%) | 0 (0%) | 62 (43.1%) | 3 (2.1%) | 35 (24.3%) | 4 (2.8%) | 9 (6.2%) | 11 (7.6%) |
| **R017** | Nov-2015 | 335 | 207 (61.8%) | 97 (29.0%) | 4 (1.2%) | 106 (31.6%) | 0 (0%) | 128 (38.2%) | 11 (3.3%) | 63 (18.8%) | 6 (1.8%) | 21 (6.3%) | 27 (8.1%) |
| **R018** | Jul-2017 | 259 | 143 (55.2%) | 67 (25.9%) | 3 (1.2%) | 73 (28.2%) | 0 (0%) | 116 (44.8%) | 5 (1.9%) | 67 (25.9%) | 5 (1.9%) | 13 (5.0%) | 26 (10.0%) |

**Supplementary Table 7.** HIV subtype distribution of p24 in 31 inland agrarian and semi-urban trading communities between 1995 and 2017

| Round | Midpoint | p24 | Pure subtypes | | | | | Recombinants | | | | | |
|---|---|---|---|---|---|---|---|---|---|---|---|---|---|
| | | N | Total | A1 | C | D | G | Total | A1/C | A1/D | A1/G | C/D | Other |
| **Total** | - | 3,399 | 2,987 (87.9%) | 952 (28%) | 75 (2.2%) | 1,955 (57.5%) | 5 (0.1%) | 412 (12.1%) | 52 (1.5%) | 170 (5%) | 41 (1.2%) | 114 (3.4%) | 35 (1%) |
| **R001** | Mar-1995 | 489 | 476 (97.3%) | 81 (16.6%) | 6 (1.2%) | 389 (79.6%) | 0 (0%) | 13 (2.7%) | 0 (0%) | 10 (2%) | 1 (0.2%) | 0 (0%) | 2 (0.4%) |
| **R009** | Jan-2003 | 419 | 392 (93.6%) | 92 (22%) | 9 (2.1%) | 291 (69.5%) | 0 (0%) | 27 (6.4%) | 0 (0%) | 12 (2.9%) | 4 (1%) | 8 (1.9%) | 3 (0.7%) |
| **R013** | Mar-2009 | 581 | 518 (89.2%) | 167 (28.7%) | 15 (2.6%) | 336 (57.8%) | 0 (0%) | 63 (10.8%) | 5 (0.9%) | 33 (5.7%) | 5 (0.9%) | 15 (2.6%) | 5 (0.9%) |
| **R014** | Sep-2010 | 277 | 246 (88.8%) | 96 (34.7%) | 5 (1.8%) | 143 (51.6%) | 2 (0.7%) | 31 (11.2%) | 4 (1.4%) | 15 (5.4%) | 4 (1.4%) | 7 (2.5%) | 1 (0.4%) |
| **R015** | Apr-2012 | 685 | 578 (84.4%) | 217 (31.7%) | 14 (2%) | 344 (50.2%) | 3 (0.4%) | 107 (15.6%) | 12 (1.8%) | 43 (6.3%) | 15 (2.2%) | 28 (4.1%) | 9 (1.3%) |
| **R016** | Apr-2014 | 326 | 277 (85%) | 97 (29.8%) | 8 (2.5%) | 172 (52.8%) | 0 (0%) | 49 (15%) | 10 (3.1%) | 20 (6.1%) | 2 (0.6%) | 13 (4%) | 4 (1.2%) |
| **R017** | Nov-2015 | 356 | 291 (81.7%) | 118 (33.1%) | 12 (3.4%) | 161 (45.2%) | 0 (0%) | 65 (18.3%) | 13 (3.7%) | 18 (5.1%) | 5 (1.4%) | 24 (6.7%) | 5 (1.4%) |
| **R018** | Jul-2017 | 266 | 209 (78.6%) | 84 (31.6%) | 6 (2.3%) | 119 (44.7%) | 0 (0%) | 57 (21.4%) | 8 (3%) | 19 (7.1%) | 5 (1.9%) | 19 (7.1%) | 6 (2.3%) |

**Supplementary Table 8.** HIV subtype distribution of gp41 in 31 inland agrarian and semi-urban trading communities between 1995 and 2017

| Round | Midpoint | gp41 | Pure subtypes | | | | | Recombinants | | | | | |
|---|---|---|---|---|---|---|---|---|---|---|---|---|---|
| | | N | Total | A1 | C | D | G | Total | A1/C | A1/D | A1/G | C/D | Other |
| **Total** | - | 2,925 | 2,755 (94.2%) | 1,100 (37.6%) | 65 (2.2%) | 1,588 (54.3%) | 2 (0.1%) | 170 (5.8%) | 14 (0.5%) | 67 (2.3%) | 62 (2.1%) | 11 (0.4%) | 16 (0.5%) |
| **R001** | Mar-1995 | 457 | 446 (97.6%) | 96 (21%) | 3 (0.7%) | 347 (75.9%) | 0 (0%) | 11 (2.4%) | 0 (0%) | 4 (0.9%) | 6 (1.3%) | 0 (0%) | 1 (0.2%) |
| **R009** | Jan-2003 | 501 | 487 (97.2%) | 145 (28.9%) | 10 (2%) | 332 (66.3%) | 0 (0%) | 14 (2.8%) | 0 (0%) | 8 (1.6%) | 5 (1%) | 1 (0.2%) | 0 (0%) |
| **R013** | Mar-2009 | 665 | 636 (95.6%) | 254 (38.2%) | 24 (3.6%) | 358 (53.8%) | 0 (0%) | 29 (4.4%) | 2 (0.3%) | 6 (0.9%) | 13 (2%) | 4 (0.6%) | 4 (0.6%) |
| **R014** | Sep-2010 | 264 | 241 (91.3%) | 116 (43.9%) | 1 (0.4%) | 124 (47%) | 0 (0%) | 23 (8.7%) | 0 (0%) | 10 (3.8%) | 11 (4.2%) | 2 (0.8%) | 0 (0%) |
| **R015** | Apr-2012 | 288 | 269 (93.4%) | 133 (46.2%) | 5 (1.7%) | 129 (44.8%) | 2 (0.7%) | 19 (6.6%) | 4 (1.4%) | 5 (1.7%) | 6 (2.1%) | 0 (0%) | 4 (1.4%) |
| **R016** | Apr-2014 | 149 | 131 (87.9%) | 71 (47.7%) | 2 (1.3%) | 58 (38.9%) | 0 (0%) | 18 (12.1%) | 1 (0.7%) | 8 (5.4%) | 5 (3.4%) | 2 (1.3%) | 2 (1.3%) |
| **R017** | Nov-2015 | 339 | 308 (90.9%) | 160 (47.2%) | 8 (2.4%) | 140 (41.3%) | 0 (0%) | 31 (9.1%) | 3 (0.9%) | 11 (3.2%) | 11 (3.2%) | 2 (0.6%) | 4 (1.2%) |
| **R018** | Jul-2017 | 262 | 237 (90.5%) | 125 (47.7%) | 12 (4.6%) | 100 (38.2%) | 0 (0%) | 25 (9.5%) | 4 (1.5%) | 15 (5.7%) | 5 (1.9%) | 0 (0%) | 1 (0.4%) |

**Supplementary Table 9.** HIV subtype distribution of p24 in four hyperendemic Lake Victoria fishing communities between 2012 and 2017

| Round | Midpoint | p24 | Pure subtypes | | | | | Recombinants | | | | | |
|---|---|---|---|---|---|---|---|---|---|---|---|---|---|
| | | N | Total | A1 | C | D | G | Total | A1/C | A1/D | A1/G | C/D | Other |
| **Total** | - | 2,321 | 1,872 (80.7%) | 651 (28%) | 111 (4.8%) | 1,099 (47.4%) | 11 (0.5%) | 449 (19.3%) | 102 (4.4%) | 146 (6.3%) | 41 (1.8%) | 128 (5.5%) | 32 (1.4%) |
| **R015** | Apr-2012 | 1,273 | 1039 (81.6%) | 340 (26.7%) | 55 (4.3%) | 641 (50.4%) | 3 (0.2%) | 234 (18.4%) | 51 (4%) | 68 (5.3%) | 21 (1.6%) | 76 (6%) | 18 (1.4%) |
| **R016** | Apr-2014 | 303 | 247 (81.5%) | 88 (29%) | 22 (7.3%) | 134 (44.2%) | 3 (1%) | 56 (18.5%) | 16 (5.3%) | 16 (5.3%) | 4 (1.3%) | 16 (5.3%) | 4 (1.3%) |
| **R017** | Nov-2015 | 439 | 360 (82%) | 145 (33%) | 19 (4.3%) | 195 (44.4%) | 1 (0.2%) | 79 (18%) | 21 (4.8%) | 34 (7.7%) | 5 (1.1%) | 16 (3.6%) | 3 (0.7%) |
| **R018** | Jul-2017 | 306 | 226 (73.9%) | 78 (25.5%) | 15 (4.9%) | 129 (42.2%) | 4 (1.3%) | 80 (26.1%) | 14 (4.6%) | 28 (9.2%) | 11 (3.6%) | 20 (6.5%) | 7 (2.3%) |

**Supplementary Table 10.** HIV subtype distribution of gp41 in four hyperendemic Lake Victoria fishing communities between 2012 and 2017

| Round | Midpoint | gp41 | Pure subtypes | | | | | Recombinants | | | | | |
|---|---|---|---|---|---|---|---|---|---|---|---|---|---|
| | | N | Total | A1 | C | D | G | Total | A1/C | A1/D | A1/G | C/D | Other |
| **Total** | - | 1,286 | 1,141 (88.7%) | 550 (42.8%) | 54 (4.2%) | 530 (41.2%) | 7 (0.5%) | 145 (11.3%) | 11 (0.9%) | 72 (5.6%) | 44 (3.4%) | 7 (0.5%) | 11 (0.9%) |
| **R015** | Apr-2012 | 480 | 428 (89.2%) | 192 (40%) | 23 (4.8%) | 211 (44%) | 2 (0.4%) | 52 (10.8%) | 3 (0.6%) | 27 (5.6%) | 16 (3.3%) | 3 (0.6%) | 3 (0.6%) |
| **R016** | Apr-2014 | 79 | 69 (87.3%) | 37 (46.8%) | 5 (6.3%) | 26 (32.9%) | 1 (1.3%) | 10 (12.7%) | 0 (0%) | 3 (3.8%) | 3 (3.8%) | 1 (1.3%) | 3 (3.8%) |
| **R017** | Nov-2015 | 427 | 381 (89.2%) | 196 (45.9%) | 15 (3.5%) | 169 (39.6%) | 1 (0.2%) | 46 (10.8%) | 3 (0.7%) | 26 (6.1%) | 12 (2.8%) | 0 (0%) | 5 (1.2%) |
| **R018** | Jul-2017 | 300 | 263 (87.7%) | 125 (41.7%) | 11 (3.7%) | 124 (41.3%) | 3 (1%) | 37 (12.3%) | 5 (1.7%) | 16 (5.3%) | 13 (4.3%) | 3 (1%) | 0 (0%) |

**Supplementary Table 11.** HIV subtype distribution of p24 and gp41 from RCCS participants living with HIV who had available sequence data from both genes in the same survey rounds in four hyperendemic Lake Victoria fishing communities between 2012 and 2017

| Round | Midpoint | N | Pure subtypes | | | | | Recombinants | | | | | |
|---|---|---|---|---|---|---|---|---|---|---|---|---|---|
| | | | Total | A1 | C | D | G | Total | A1/C | A1/D | A1/G | C/D | Other |
| **Total** | - | 1,270 | 692 (54.5%) | 272 (21.4%) | 26 (2.0%) | 387 (30.5%) | 7 (0.6%) | 578 (45.5%) | 60 (4.7%) | 325 (25.6%) | 21 (1.7%) | 64 (5.0%) | 108 (8.5%) |
| **R015** | Apr-2012 | 475 | 270 (56.8%) | 101 (21.3%) | 9 (1.9%) | 158 (33.3%) | 2 (0.4%) | 205 (43.2%) | 17 (3.6%) | 115 (24.2%) | 6 (1.3%) | 31 (6.5%) | 36 (7.6%) |
| **R016** | Apr-2014 | 78 | 41 (52.6%) | 18 (23.1%) | 4 (5.1%) | 18 (23.1%) | 1 (1.3%) | 37 (47.4%) | 4 (5.1%) | 16 (20.5%) | 0 (0%) | 5 (6.4%) | 12 (15.4%) |
| **R017** | Nov-2015 | 417 | 236 (56.6%) | 98 (23.5%) | 9 (2.2%) | 128 (30.7%) | 1 (0.2%) | 181 (43.4%) | 23 (5.5%) | 113 (27.1%) | 7 (1.7%) | 10 (2.4%) | 28 (6.7%) |
| **R018** | Jul-2017 | 300 | 145 (48.3%) | 55 (18.3%) | 4 (1.3%) | 83 (27.7%) | 3 (1%) | 155 (51.7%) | 16 (5.3%) | 81 (27.0%) | 8 (2.7%) | 18 (6.0%) | 32 (10.7%) |

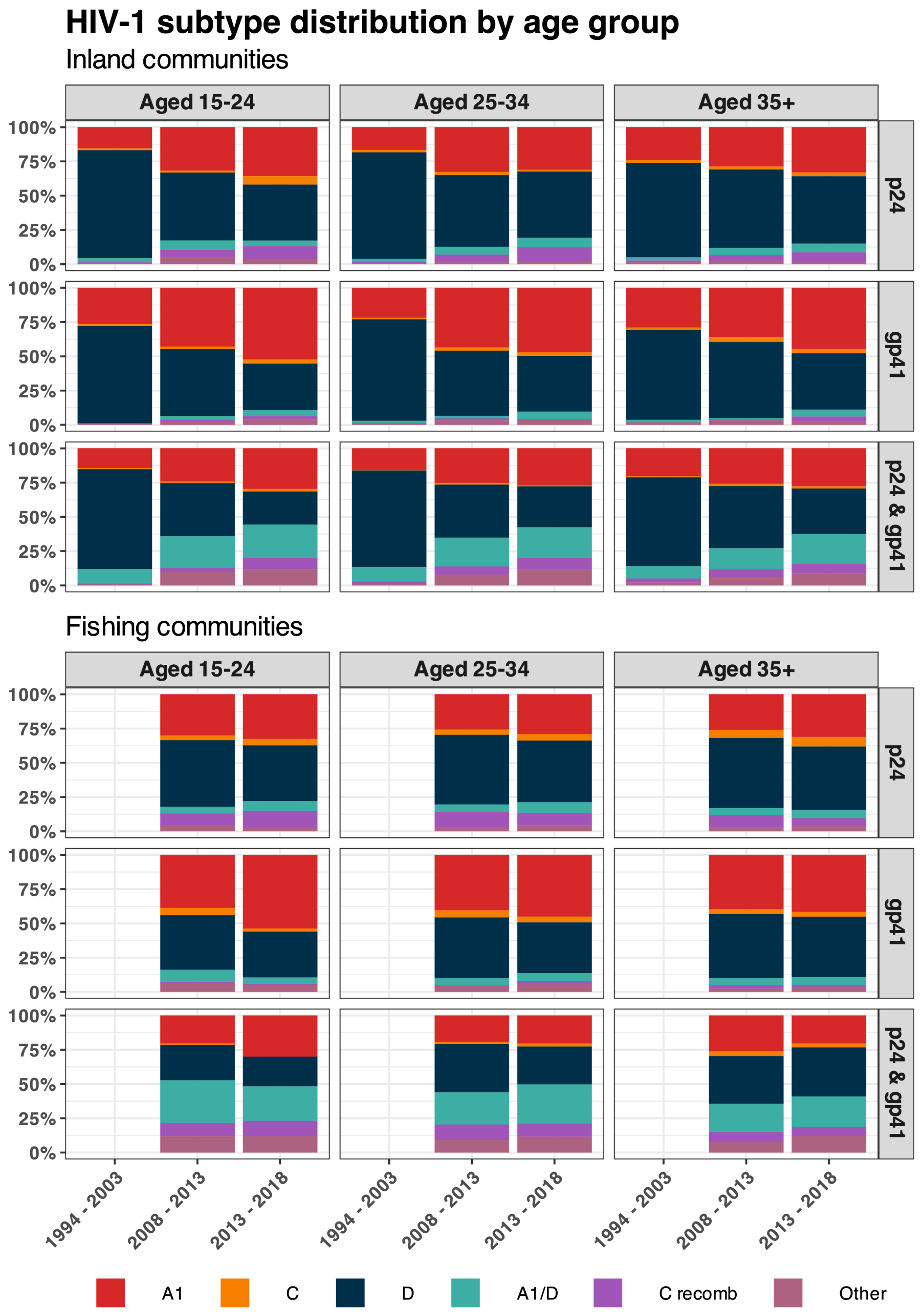


**Supplementary Figure 3.** HIV subtype distribution by age group in 31 inland agrarian and semi-urban trading communities and four hyperendemic Lake Victoria fishing communities between 1995 and 2017

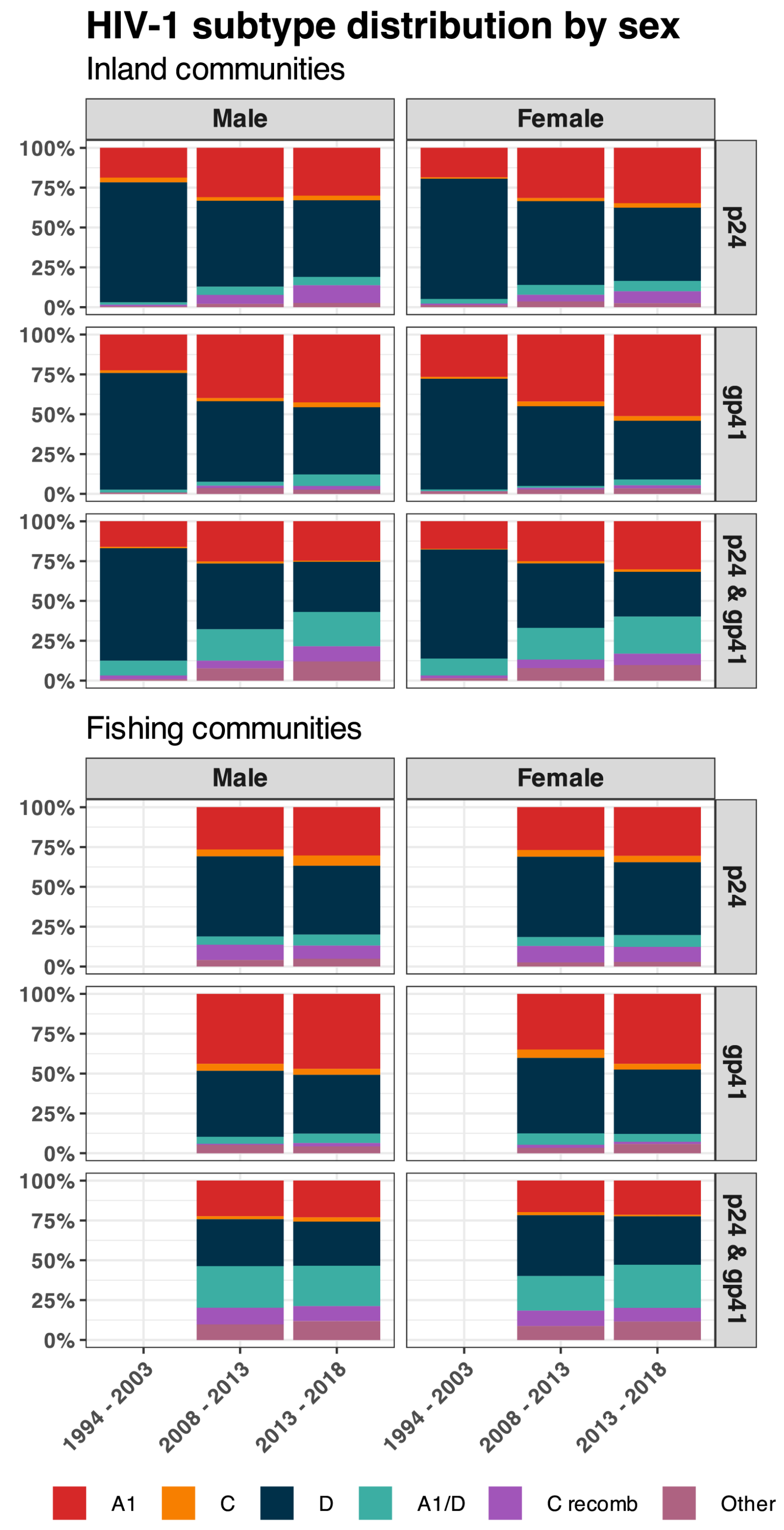


**Supplementary Figure 4.** HIV subtype distribution by sex in 31 inland agrarian and semi-urban trading communities and four hyperendemic Lake Victoria fishing communities between 1995 and 2017

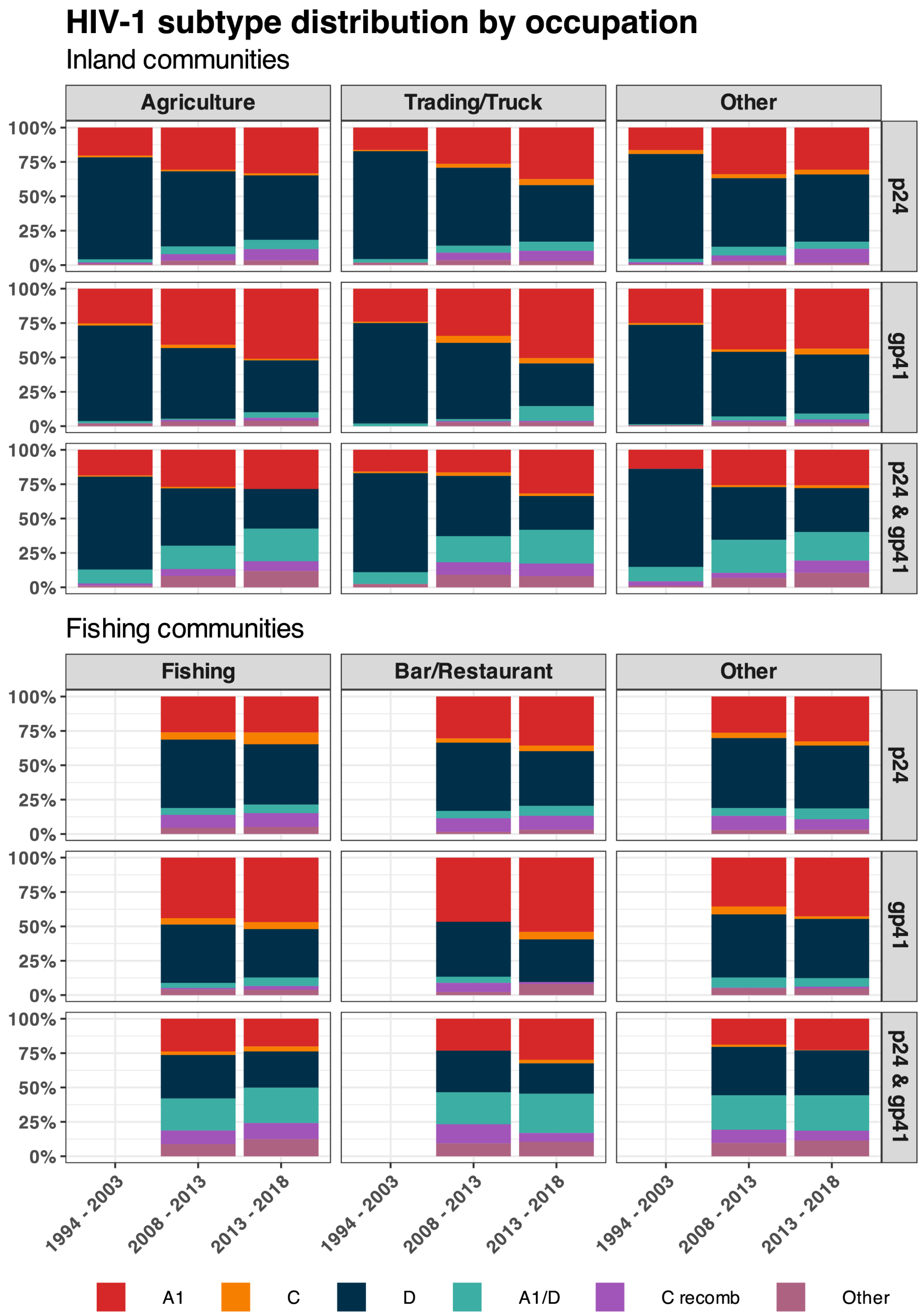


**Supplementary Figure 5.** HIV subtype distribution by occupation in 31 inland agrarian and semi-urban trading communities and four hyperendemic Lake Victoria fishing communities between 1995 and 2017

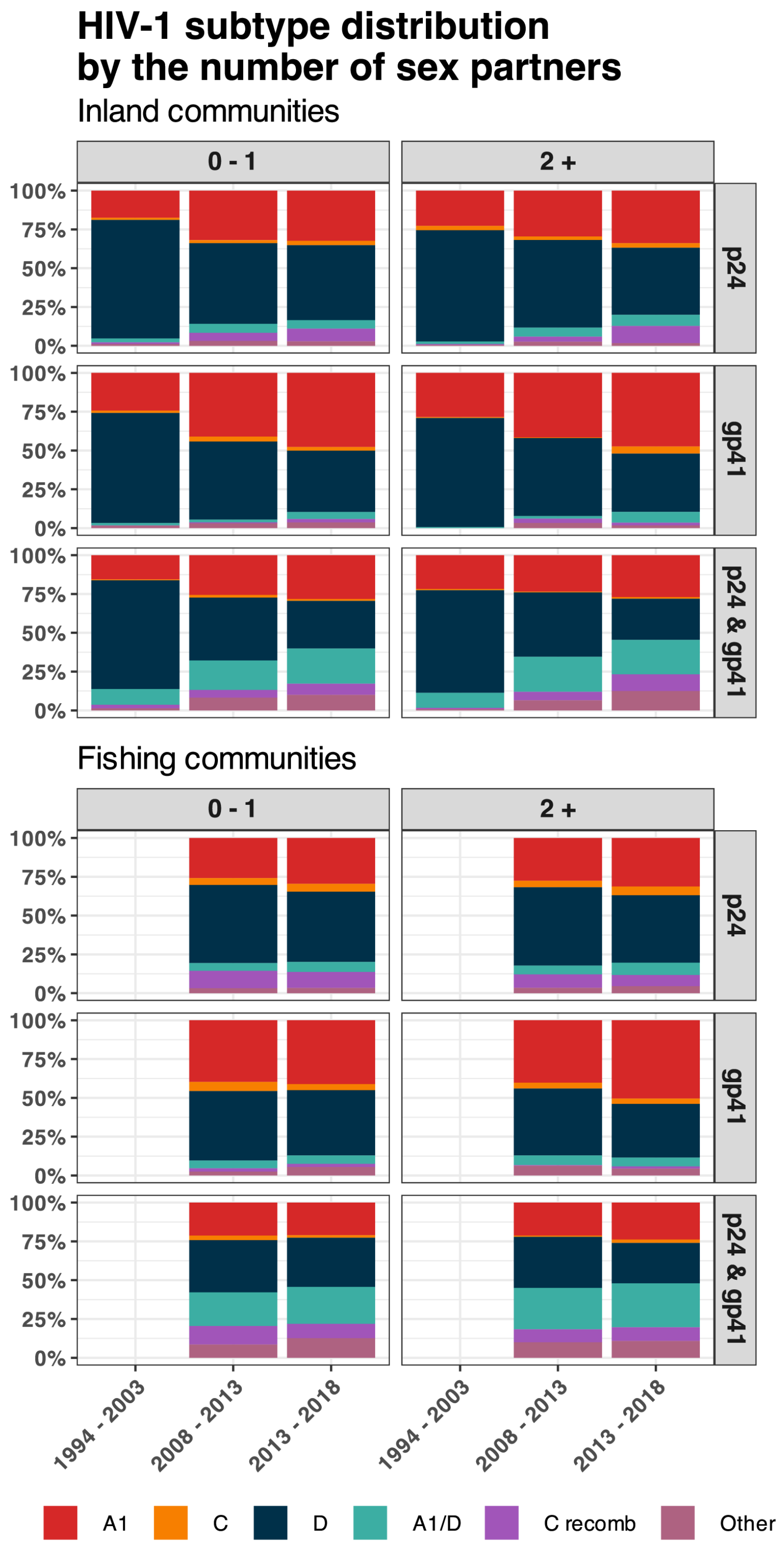


**Supplementary Figure 6.** HIV-1 subtype distribution by the number of sexual partners in the past year in 31 inland agrarian and semi-urban trading communities and four hyperendemic Lake Victoria fishing communities between 1995 and 2017

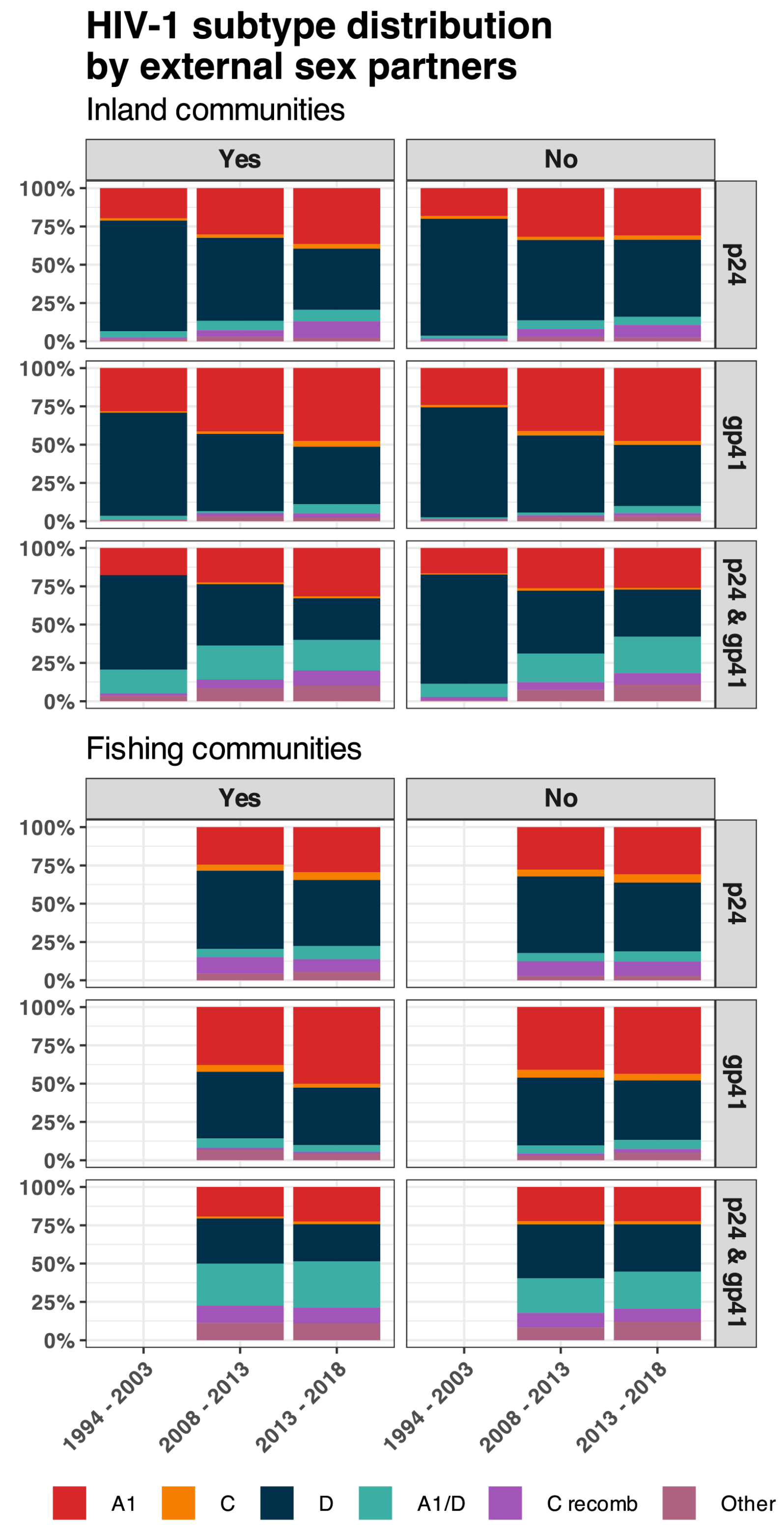


**Supplementary Figure 7.** HIV subtype distribution by the presence of external sexual partners in the past year in 31 inland agrarian and semi-urban trading communities and four hyperendemic Lake Victoria fishing communities between 1995 and 2017

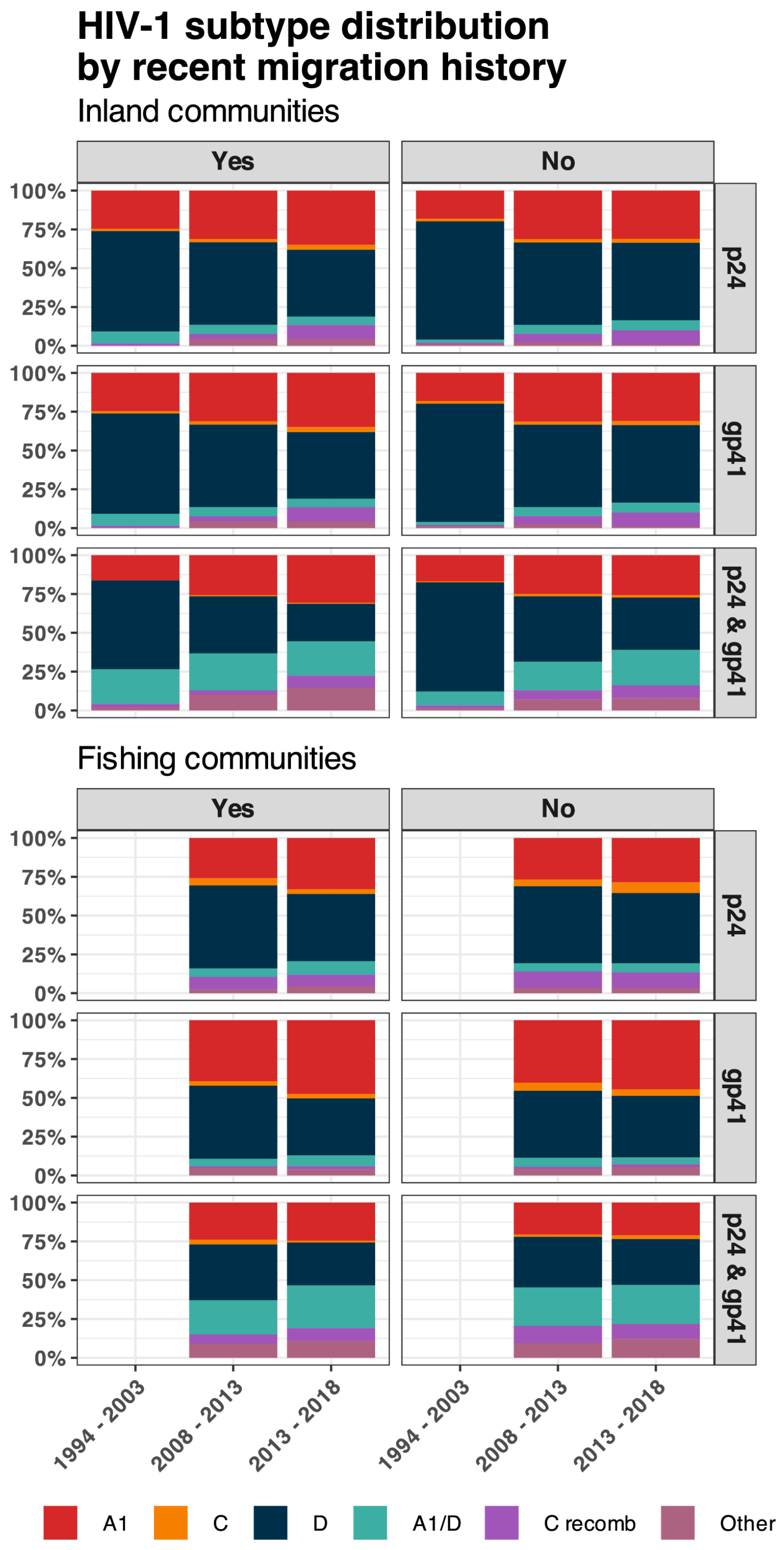


**Supplementary Figure 8.** HIV-1 subtype distribution by recent history of migration in the past year in 31 inland agrarian and semi-urban trading communities and four hyperendemic Lake Victoria fishing communities between 1995 and 2017

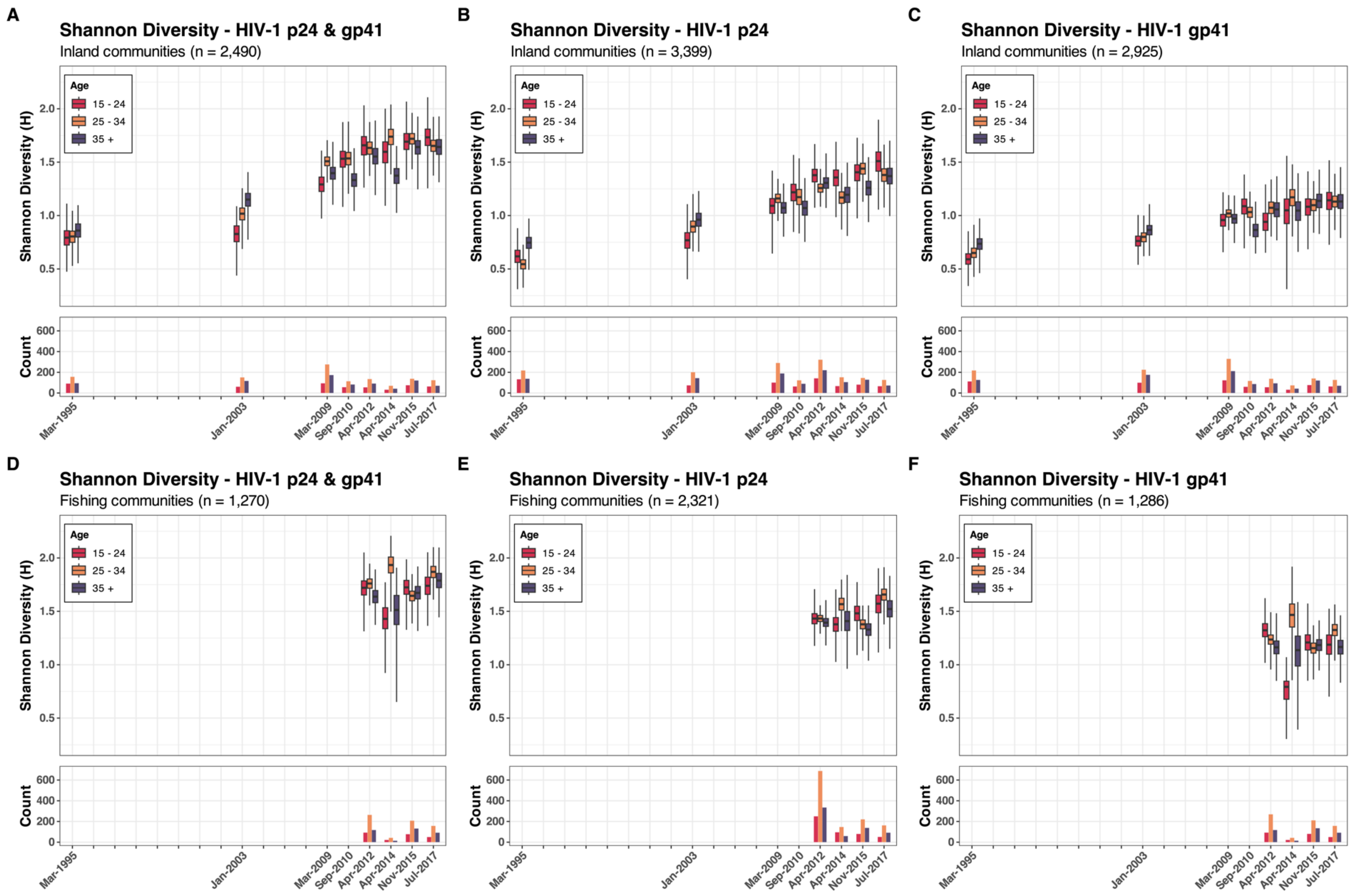


**Supplementary Figure 9.** The Shannon diversity index of p24, gp41, and both p24 and gp41 by age group in 31 inland agrarian and semi-urban trading communities and four hyperendemic Lake Victoria fishing communities between 1995 and 2017

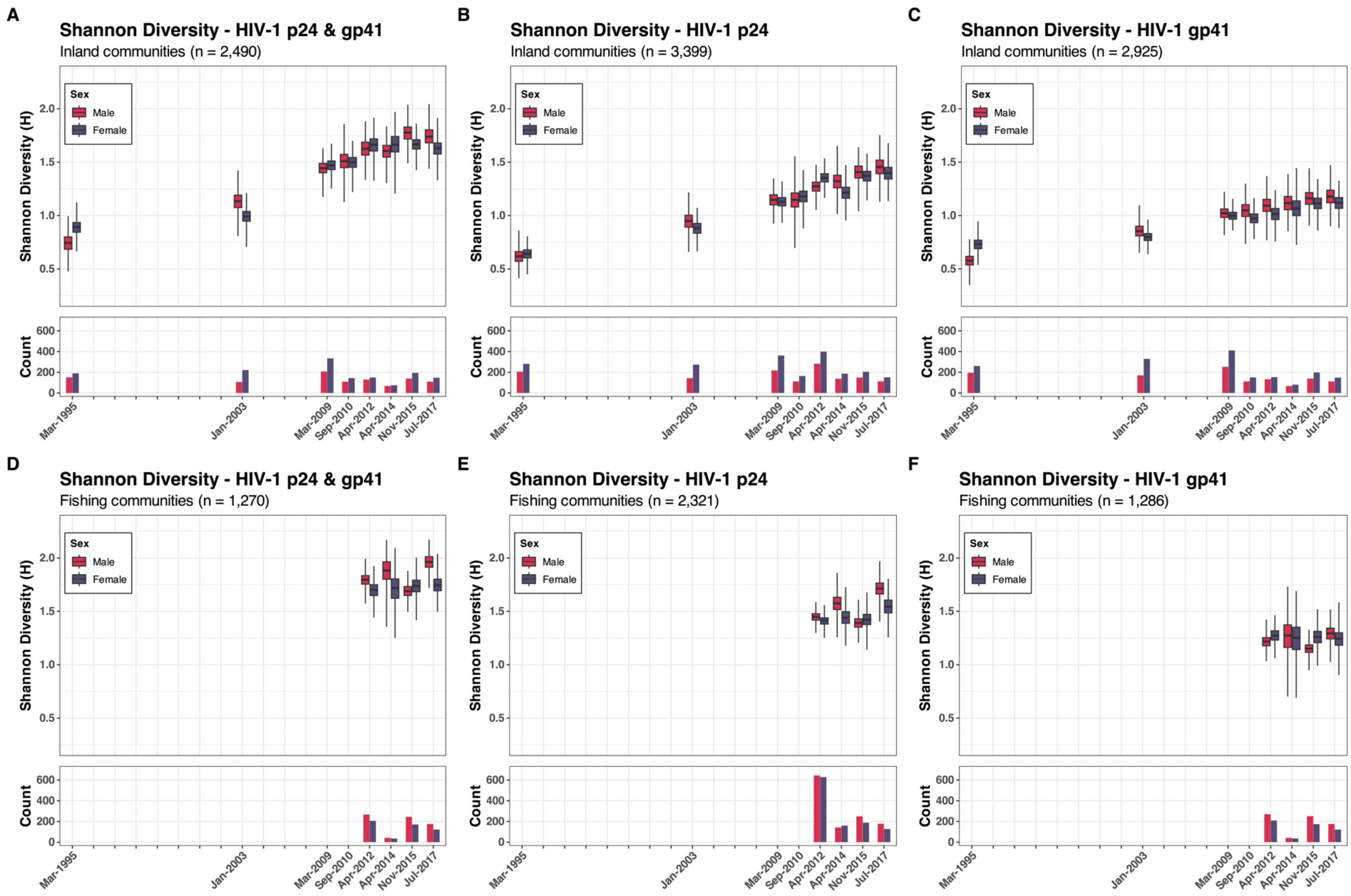


**Supplementary Figure 10.** The Shannon diversity index of p24, gp41, and both p24 and gp41 by sex in 31 inland agrarian and semi-urban trading communities and four hyperendemic Lake Victoria fishing communities between 1995 and 2017

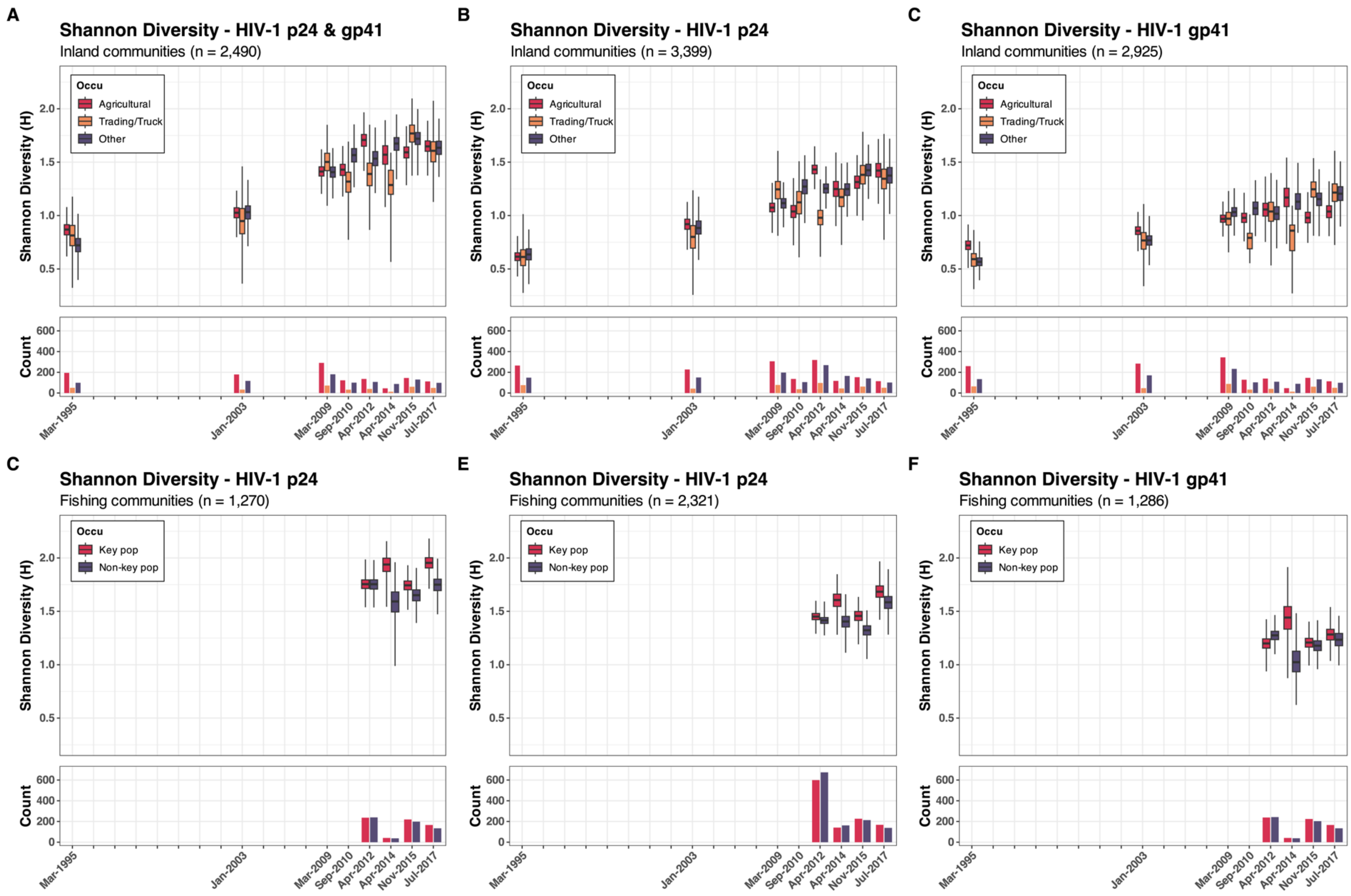


**Supplementary Figure 11.** The Shannon diversity index of p24, gp41, and both p24 and gp41 by occupation in 31 inland agrarian and semi-urban trading communities and four hyperendemic Lake Victoria fishing communities between 1995 and 2017

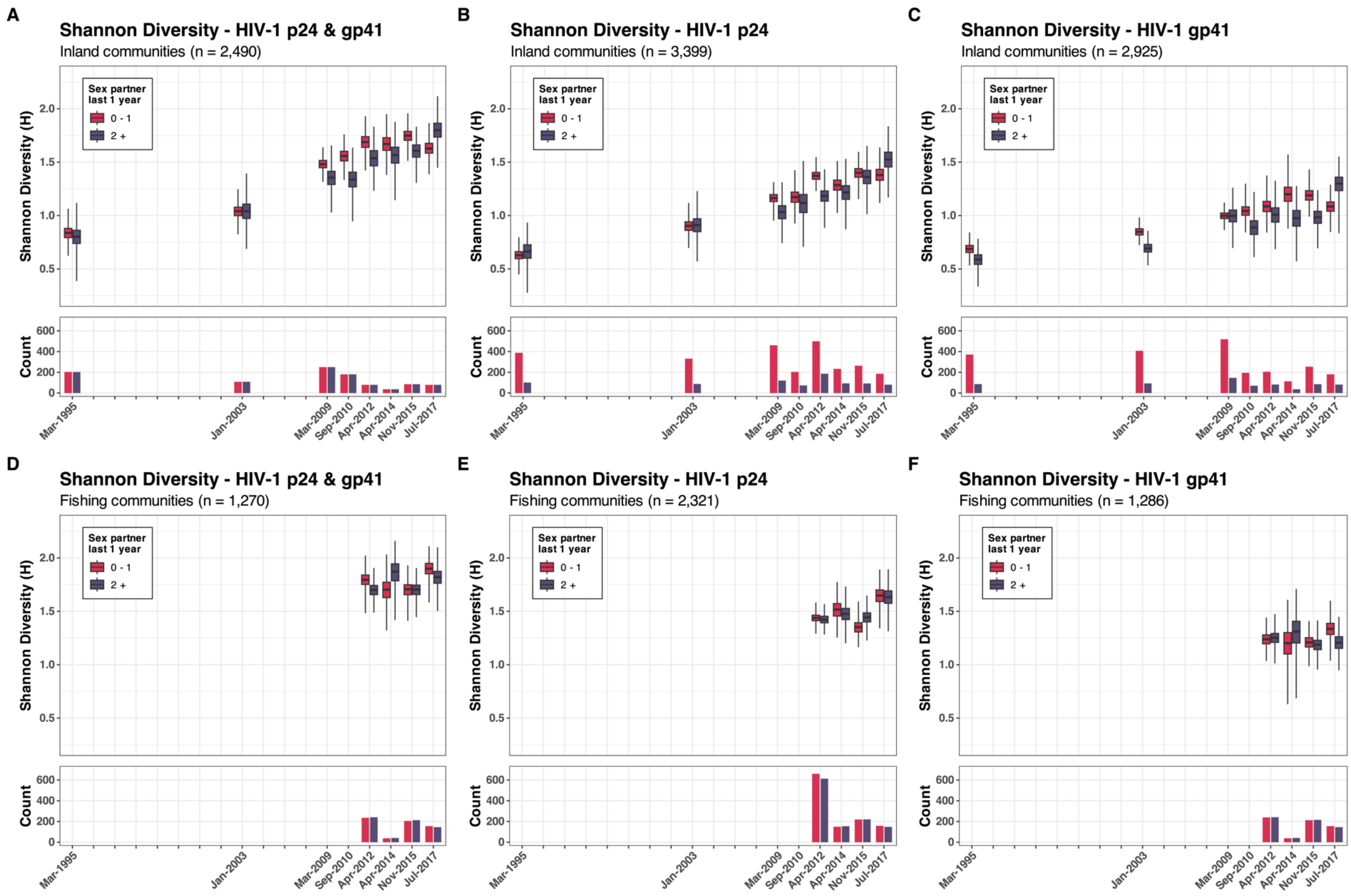


**Supplementary Figure 12.** The Shannon diversity index of p24 and gp41, and both p24 and gp41 by the number of sexual partners in the past year in 31 inland agrarian and semi-urban trading communities and four hyperendemic Lake Victoria fishing communities between 1995 and 2017

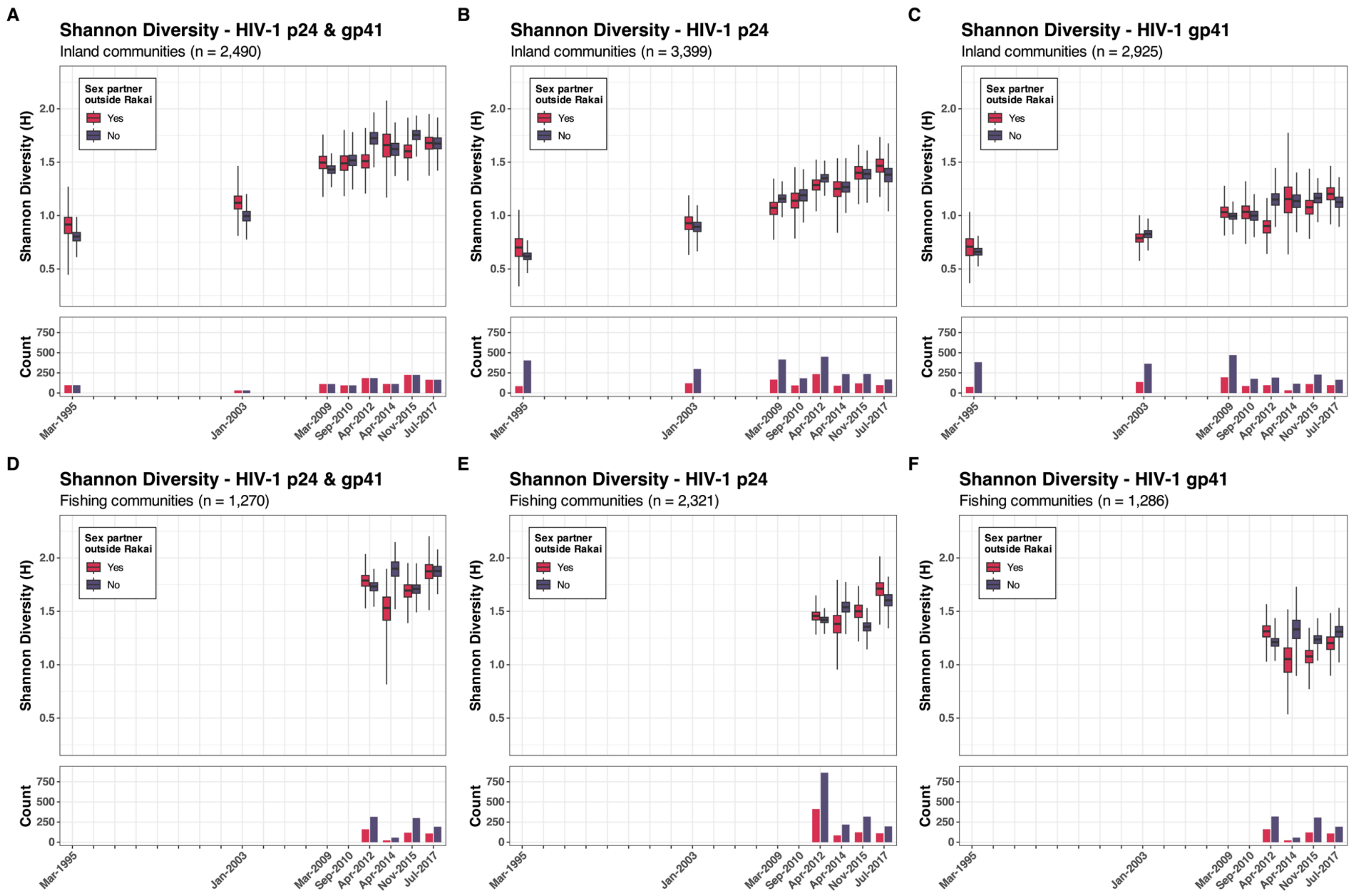


**Supplementary Figure 13.** The Shannon diversity index of p24, gp41, and both p24 and gp41 by the presence of external sexual partners in the past year in 31 inland agrarian and semi-urban trading communities and four hyperendemic Lake Victoria fishing communities between 1995 and 2017

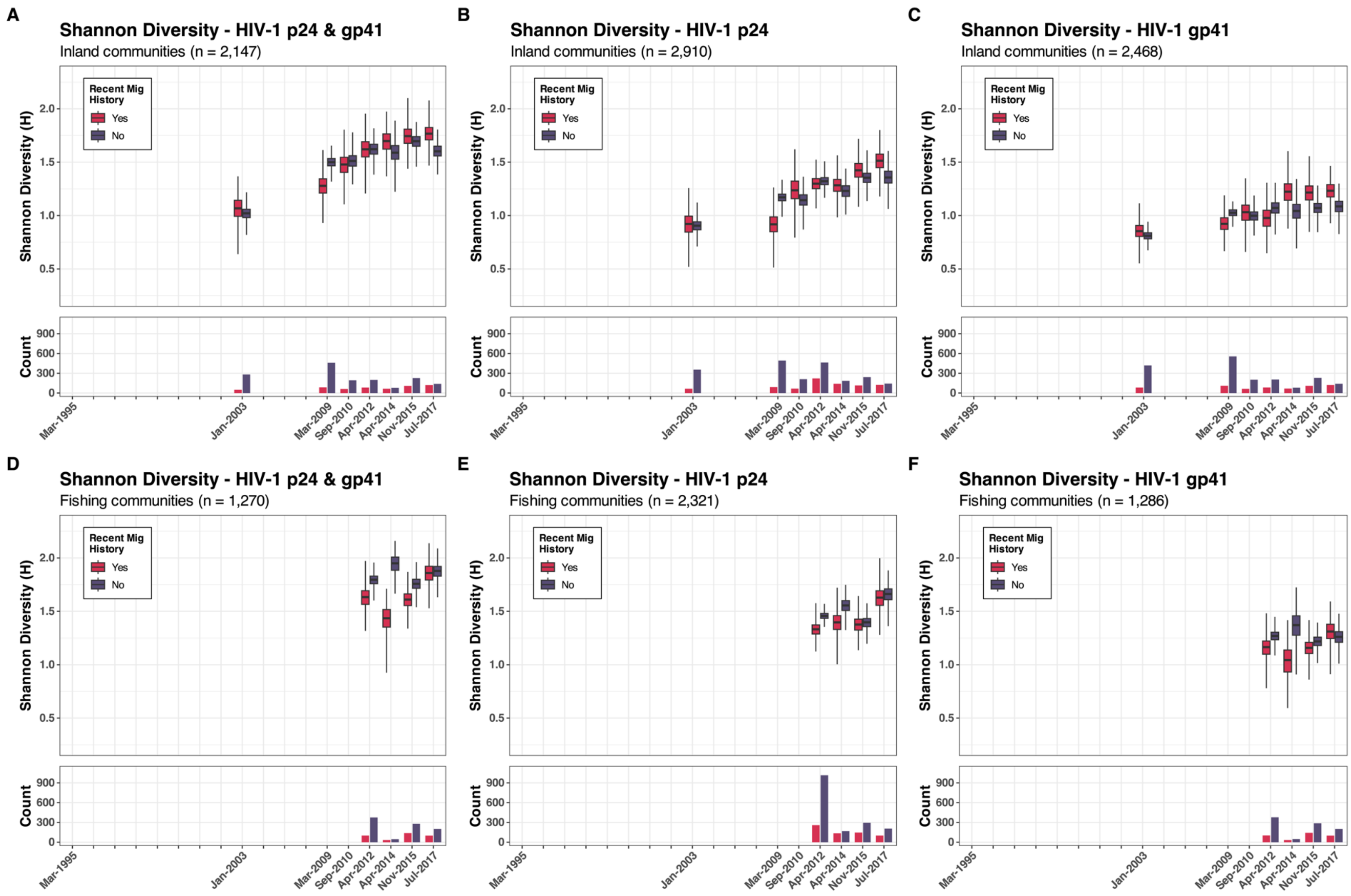


**Supplementary Figure 14.** The Shannon diversity index of p24, gp41, and both p24 and gp41 by recent history of migration in the past year in 31 inland agrarian and semi-urban trading communities and four hyperendemic Lake Victoria fishing communities between 1995 and 2017

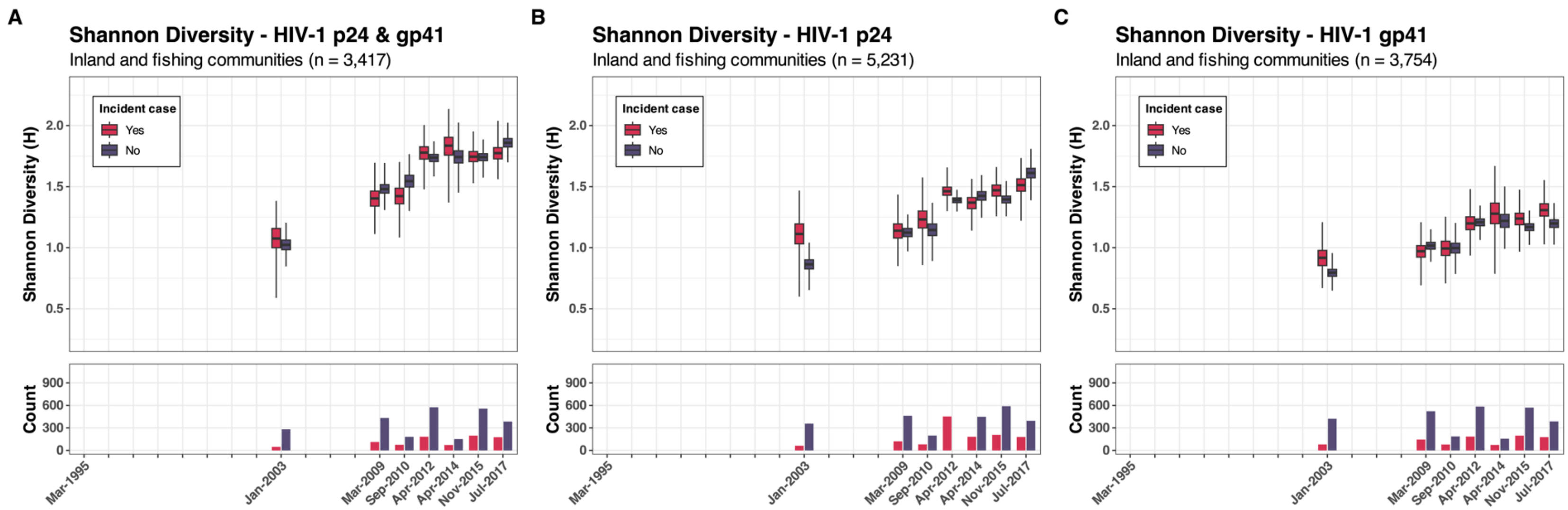


**Supplementary Figure 15.** The Shannon diversity index of p24 and gp41, and both p24 and gp41 by incident case in Rakai between 1995 and 2017

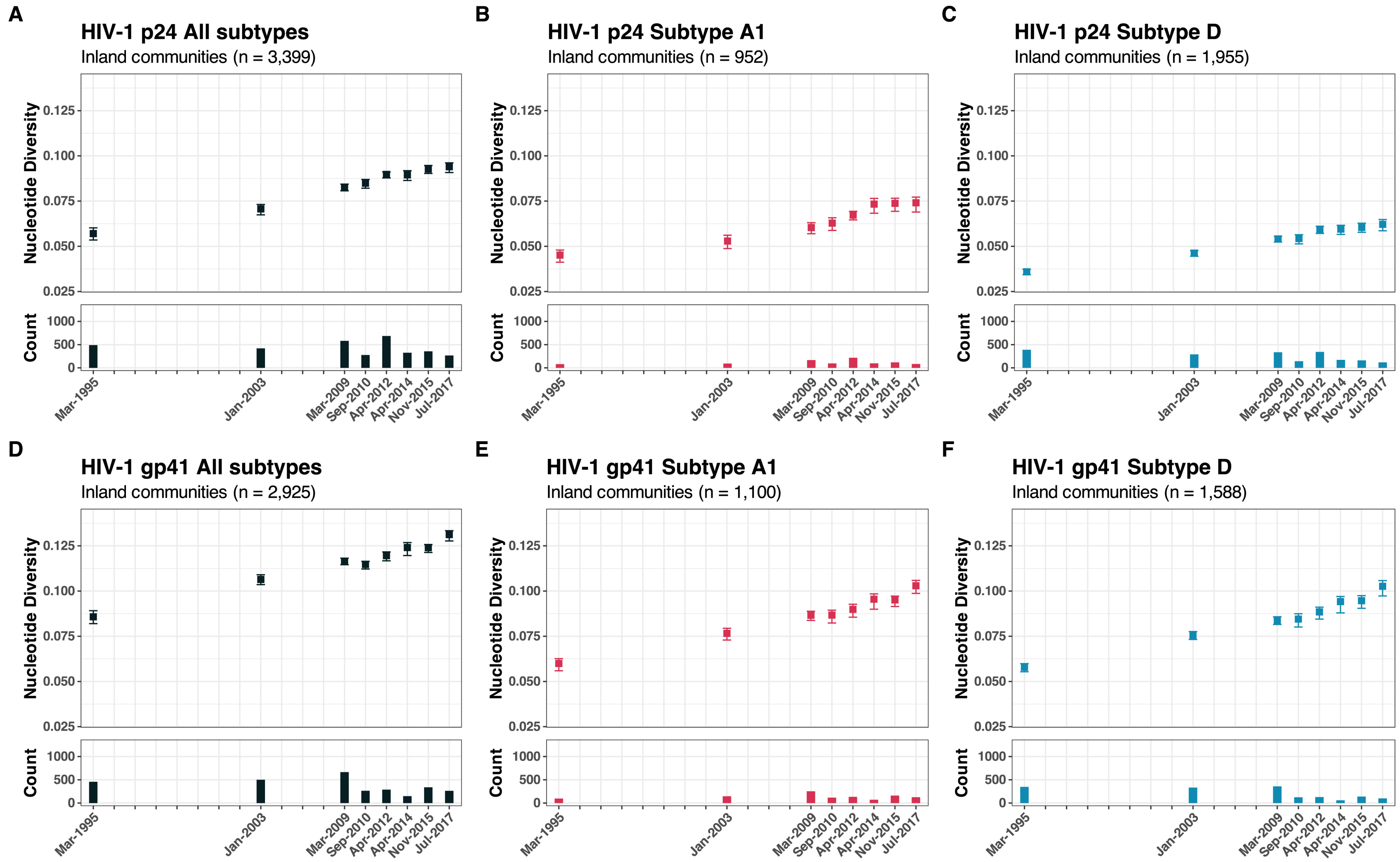


**Supplementary Figure 16.** The overall and subtype specific (A1 & D) nucleotide genetic diversity of p24 and gp41 in 31 inland agrarian and semi-urban trading communities between 1995 and 2017

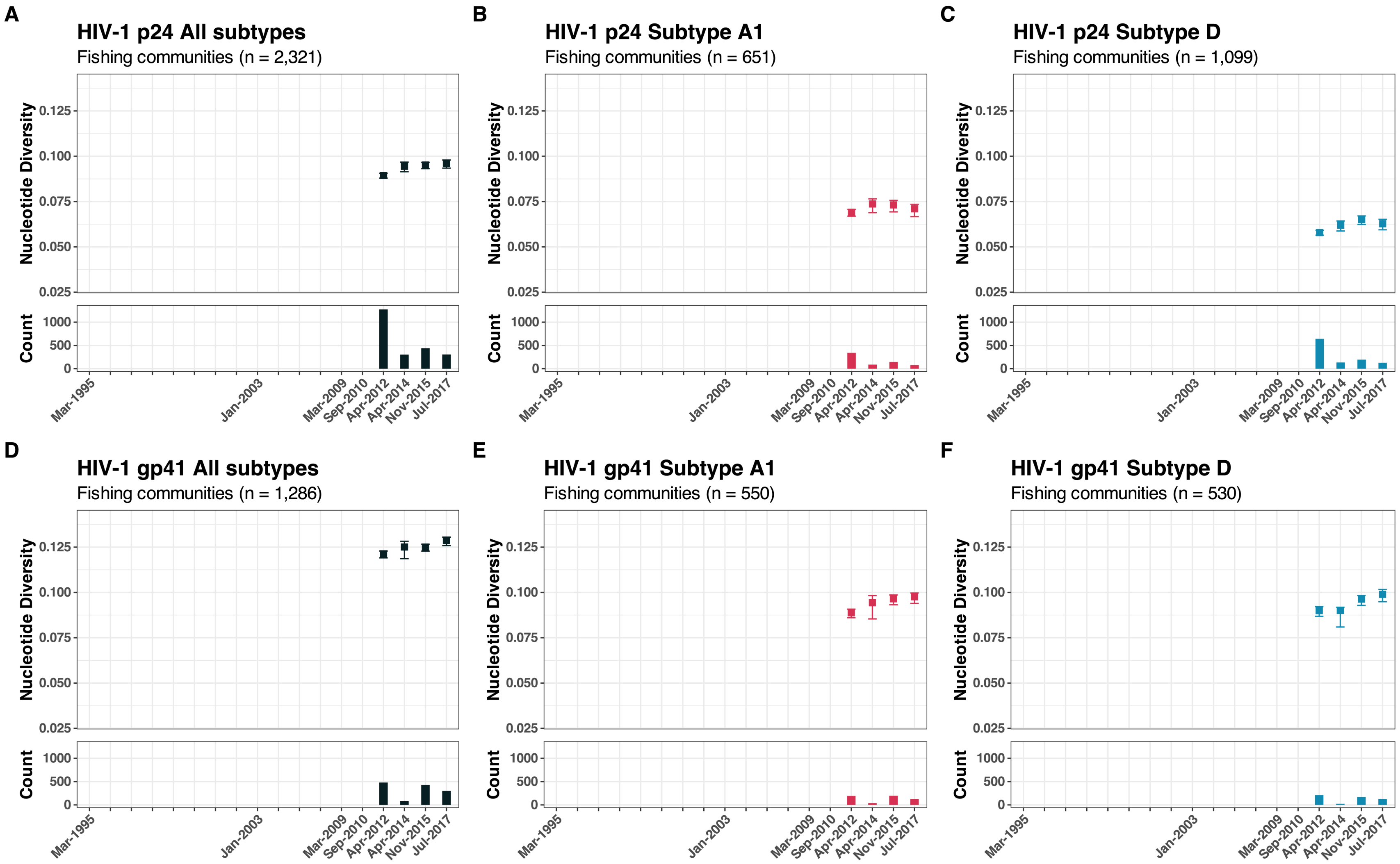


**Supplementary Figure 17.** The overall and subtype specific (A1 & D) nucleotide genetic diversity of p24 and gp41 in four hyperendemic Lake Victoria fishing communities between 2012 and 2017

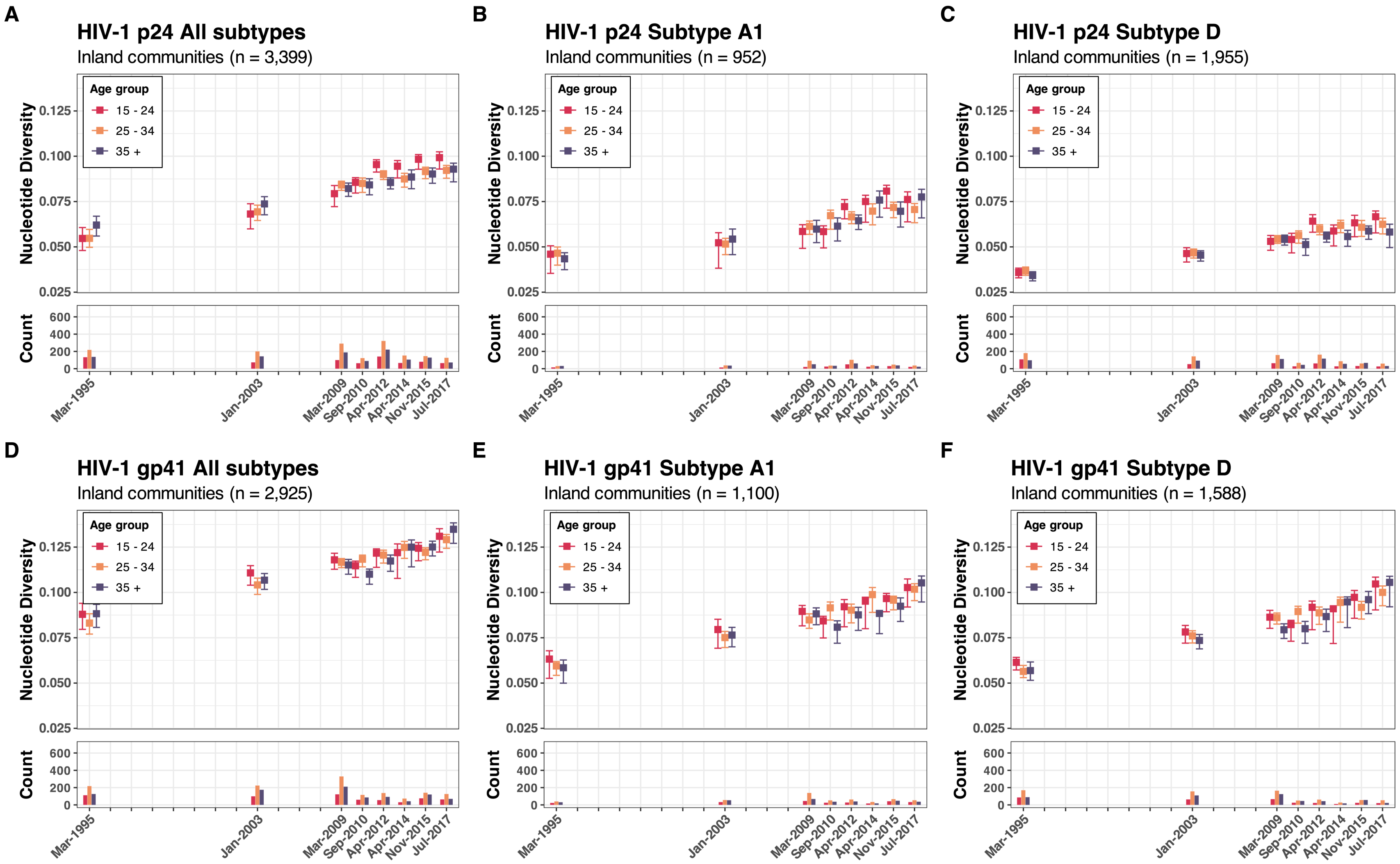


**Supplementary Figure 18.** The overall and subtype specific (A1 & D) nucleotide genetic diversity of p24 and gp41 by age group in 31 inland agrarian and semi-urban trading communities between 1995 and 2017

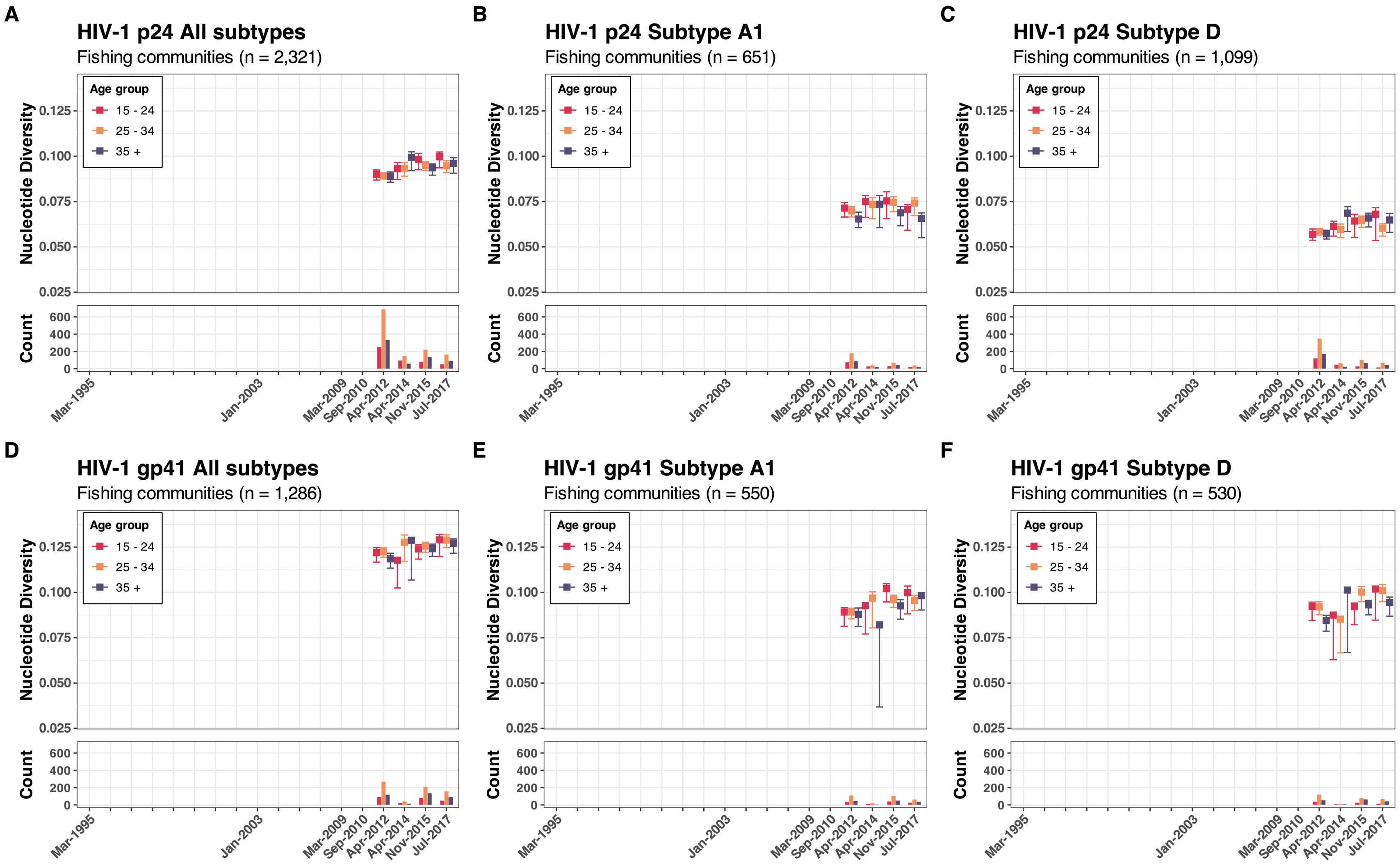


**Supplementary Figure 19.** The overall and subtype specific (A1 & D) nucleotide genetic diversity of p24 and gp41 by age group in four hyperendemic Lake Victoria fishing communities between 2012 and 2017

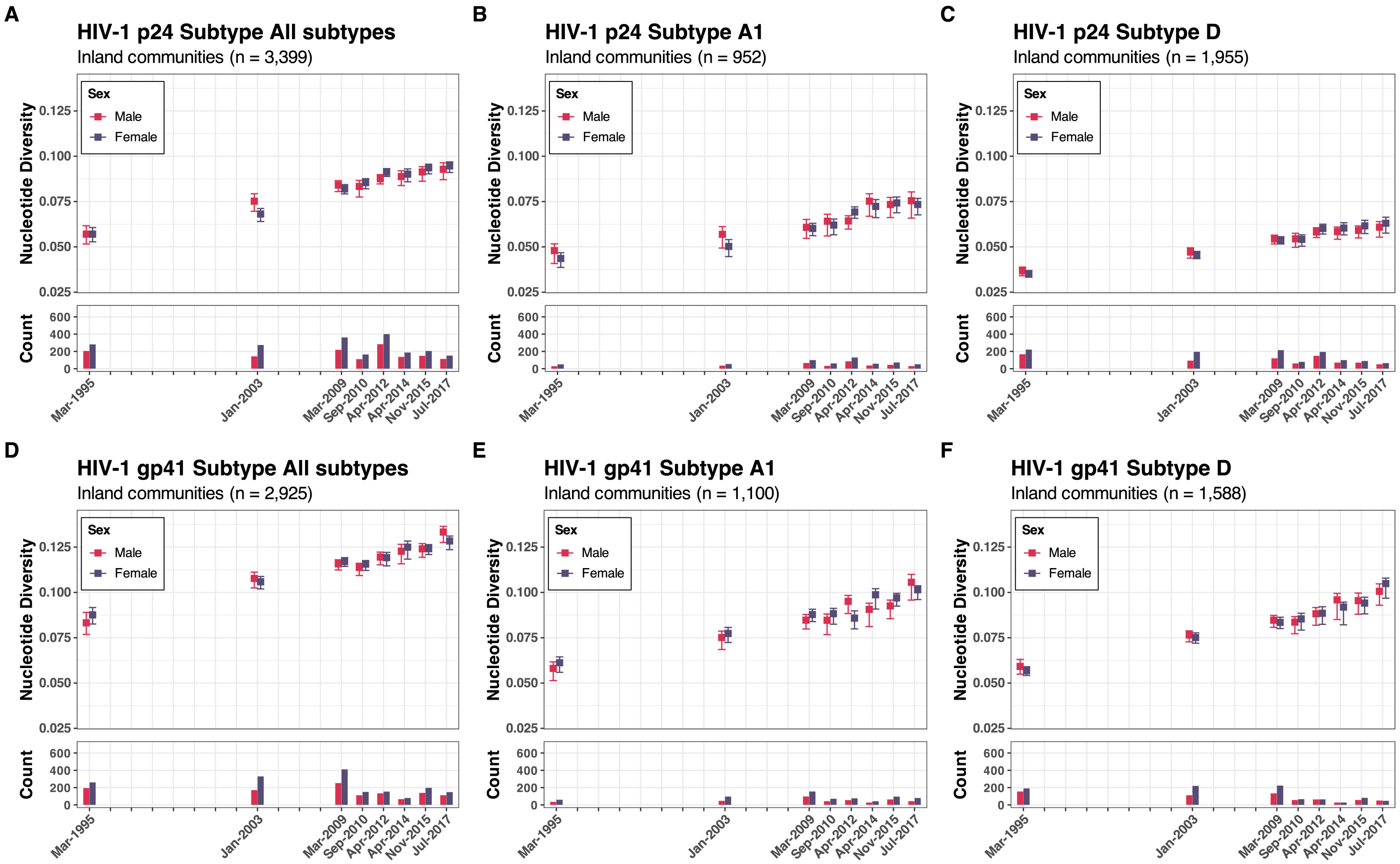


**Supplementary Figure 20.** The overall and subtype specific (A1 & D) nucleotide genetic diversity of p24 and gp41 by sex in 31 inland agrarian and semi-urban trading communities between 1995 and 2017

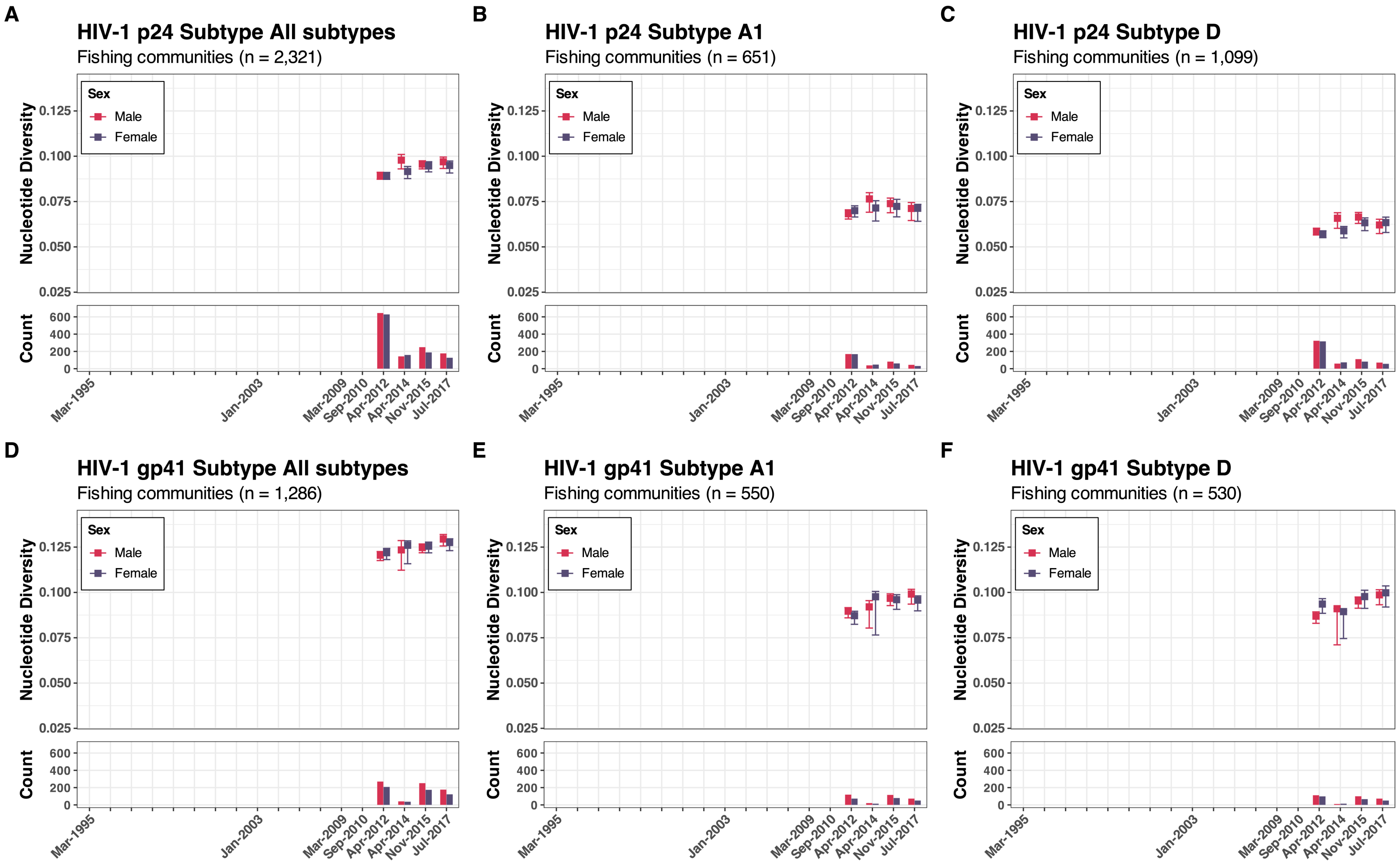


**Supplementary Figure 21.** The overall and subtype specific (A1 & D) nucleotide genetic diversity of p24 and gp41 by sex in four hyperendemic Lake Victoria fishing communities between 2012 and 2017

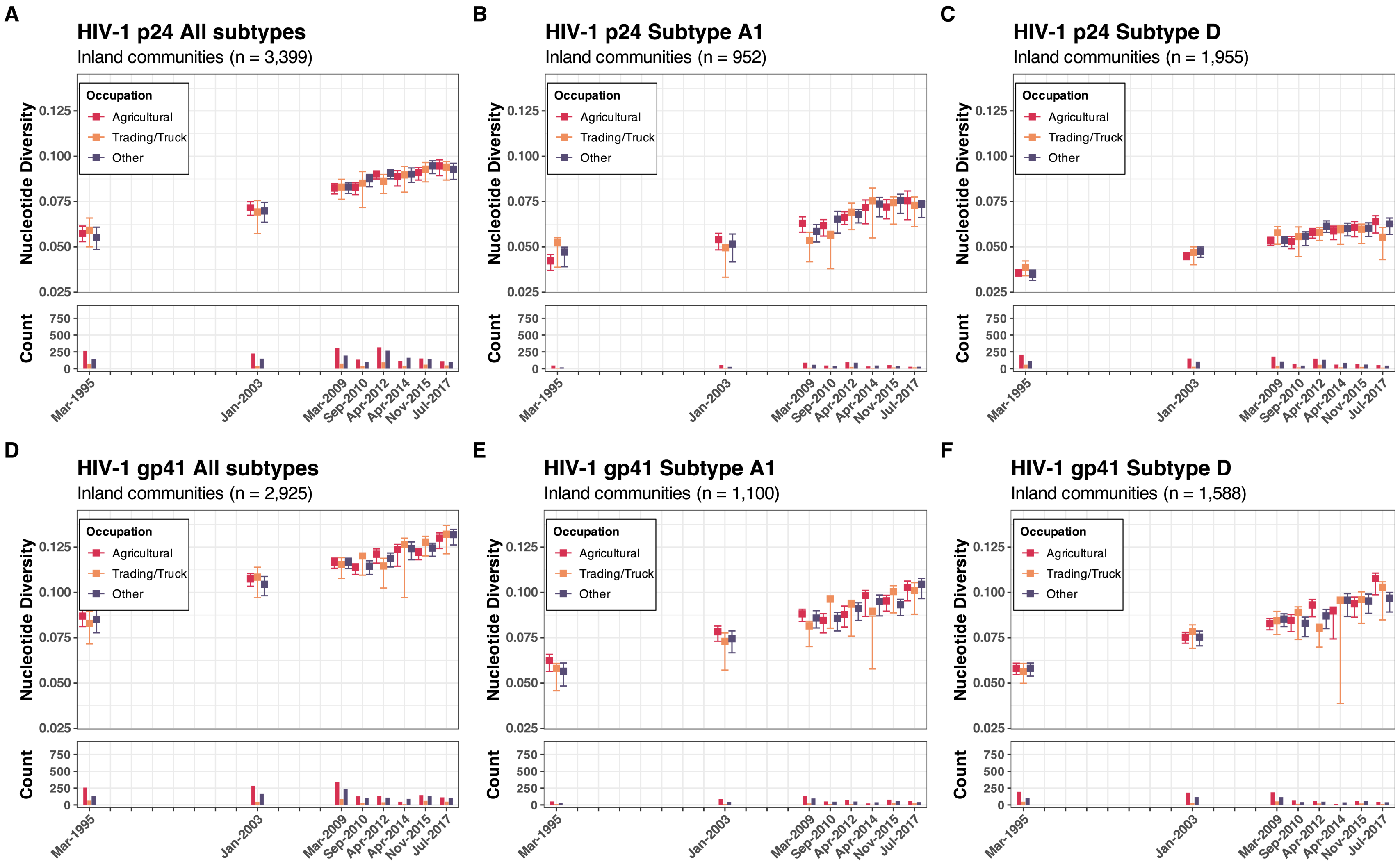


**Supplementary Figure 22.** The overall and subtype specific (A1 & D) nucleotide genetic diversity of p24 and gp41 by occupation in 31 inland agrarian and semi-urban trading communities between 1995 and 2017

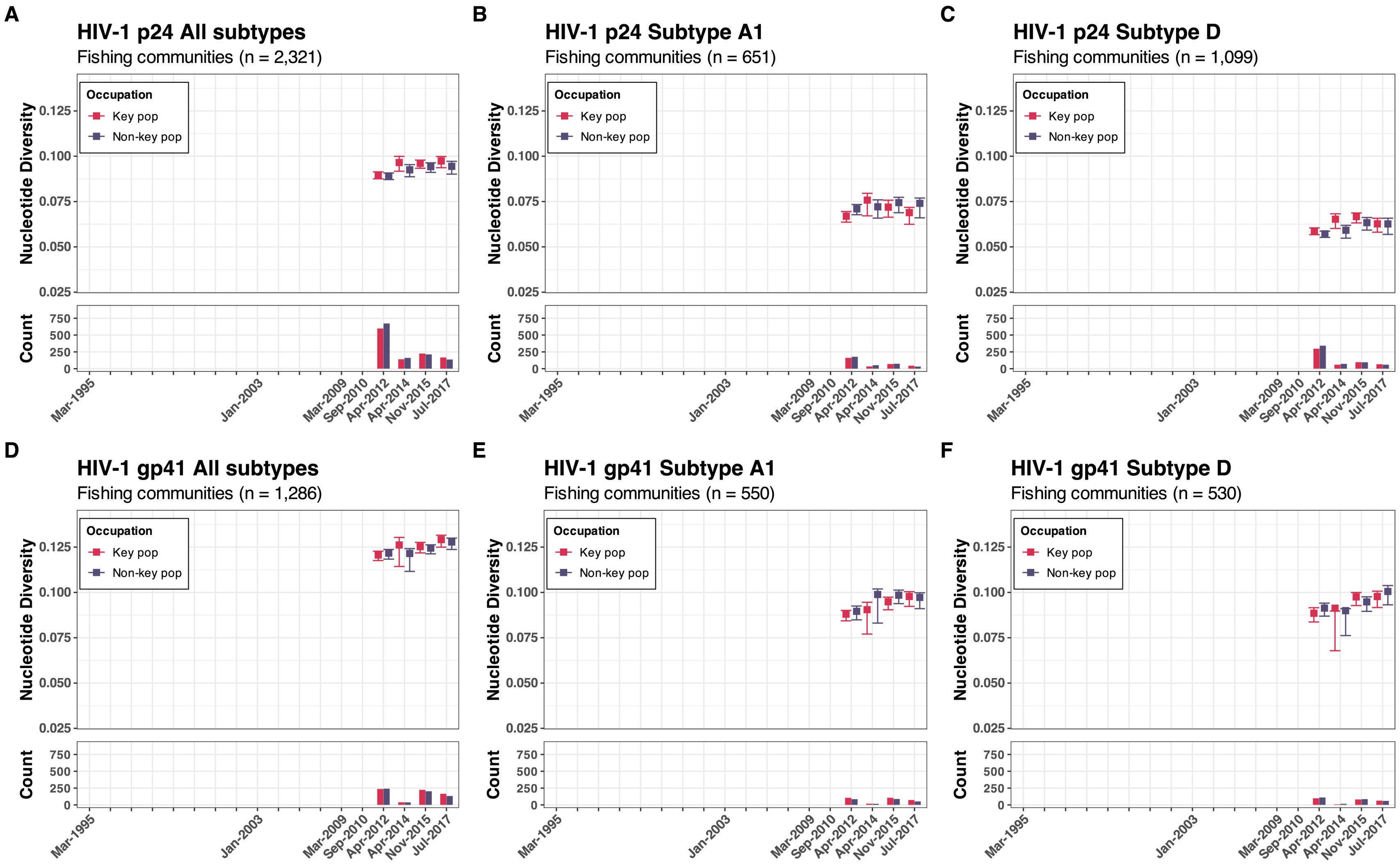


**Supplementary Figure 23.** The overall and subtype specific (A1 & D) nucleotide genetic diversity of p24 and gp41 by occupation in four hyperendemic Lake Victoria fishing communities between 2012 and 2017

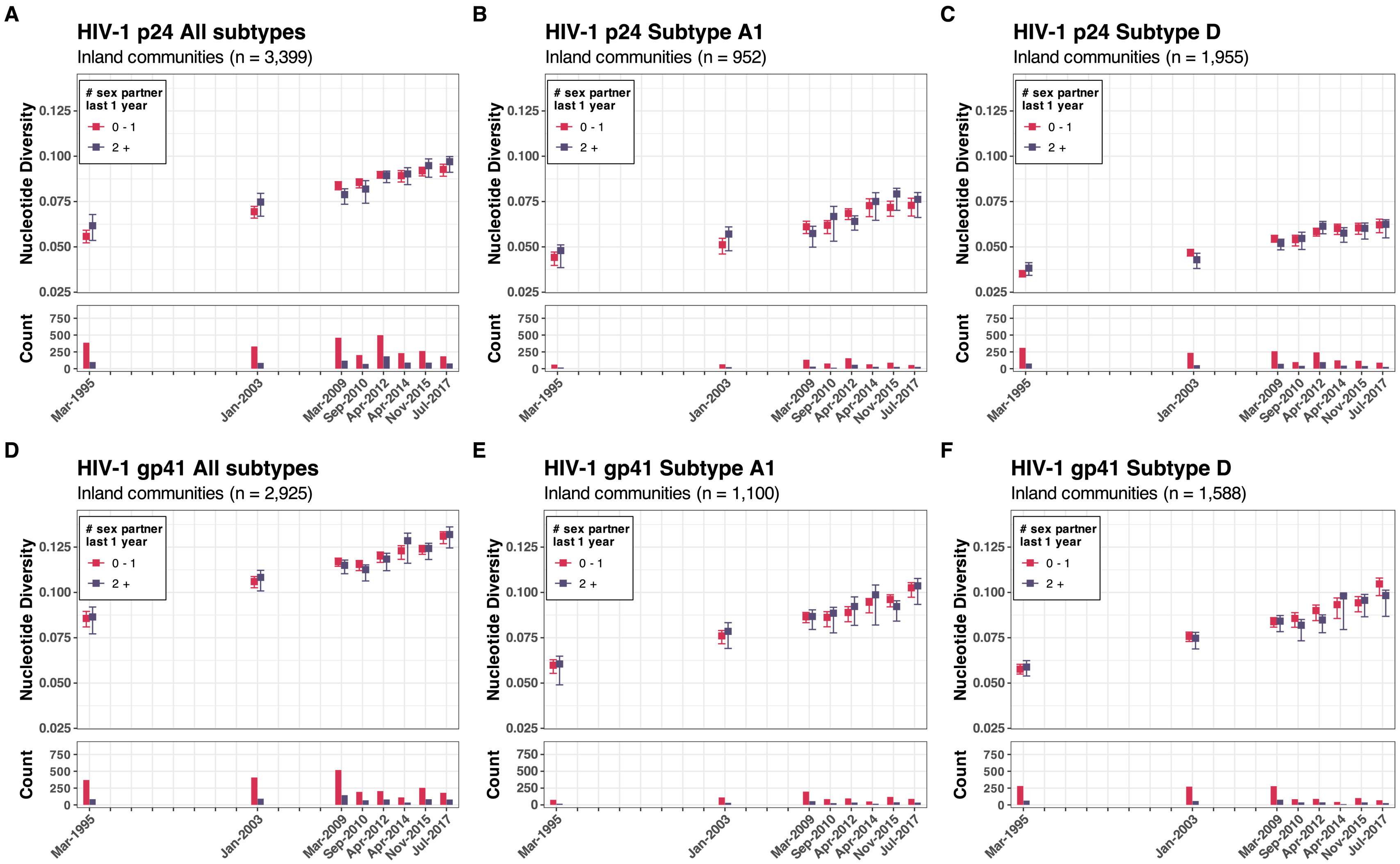


**Supplementary Figure 24.** The overall and subtype specific (A1 & D) nucleotide genetic diversity of p24 and gp41 by the number of sexual partners in the past year in 31 inland agrarian and semi-urban trading communities between 1995 and 2017

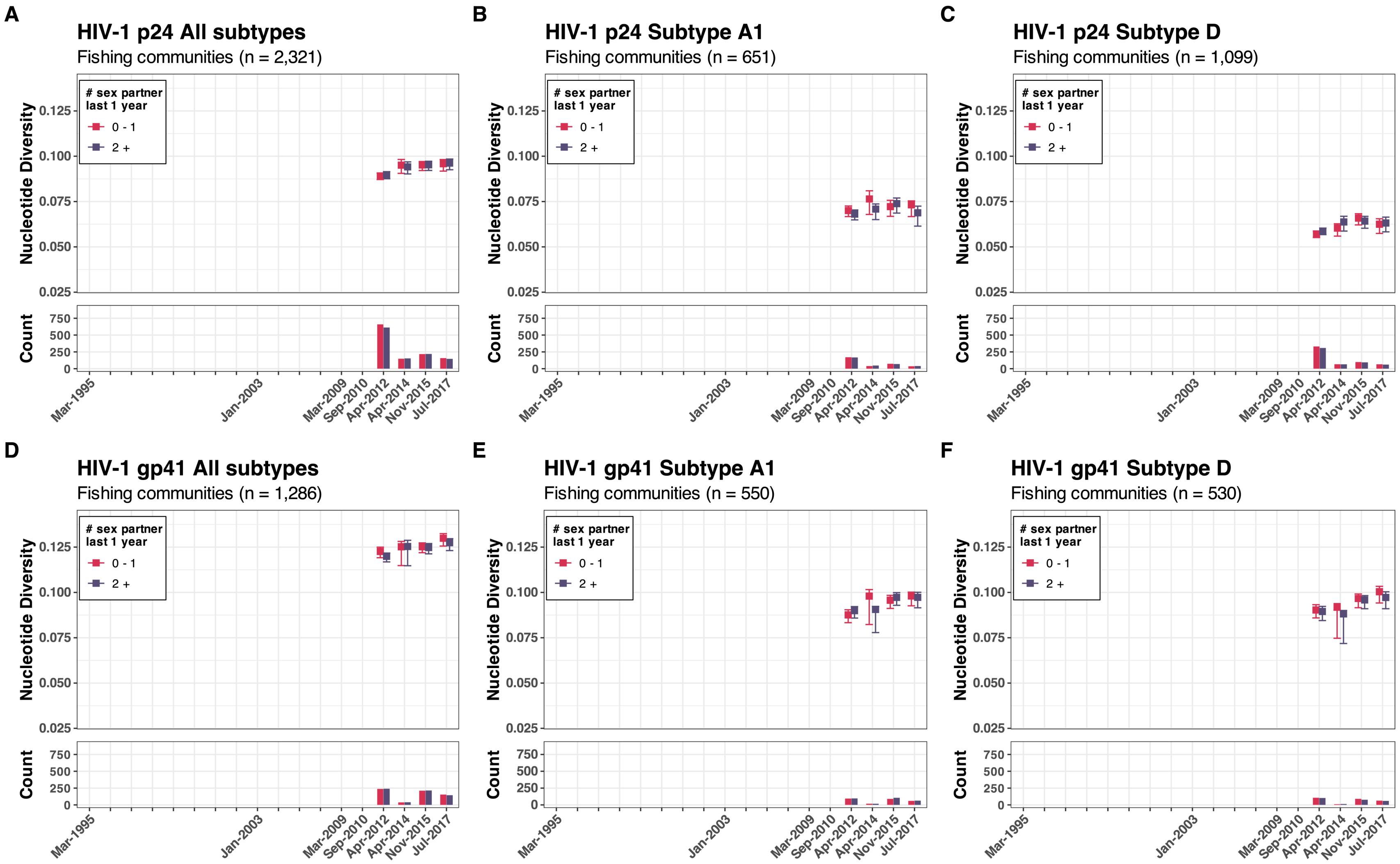


**Supplementary Figure 25.** The overall and subtype specific (A1 & D) nucleotide genetic diversity of p24 and gp41 by the number of sexual partners in the past year in four hyperendemic Lake Victoria fishing communities between 2012 and 2017

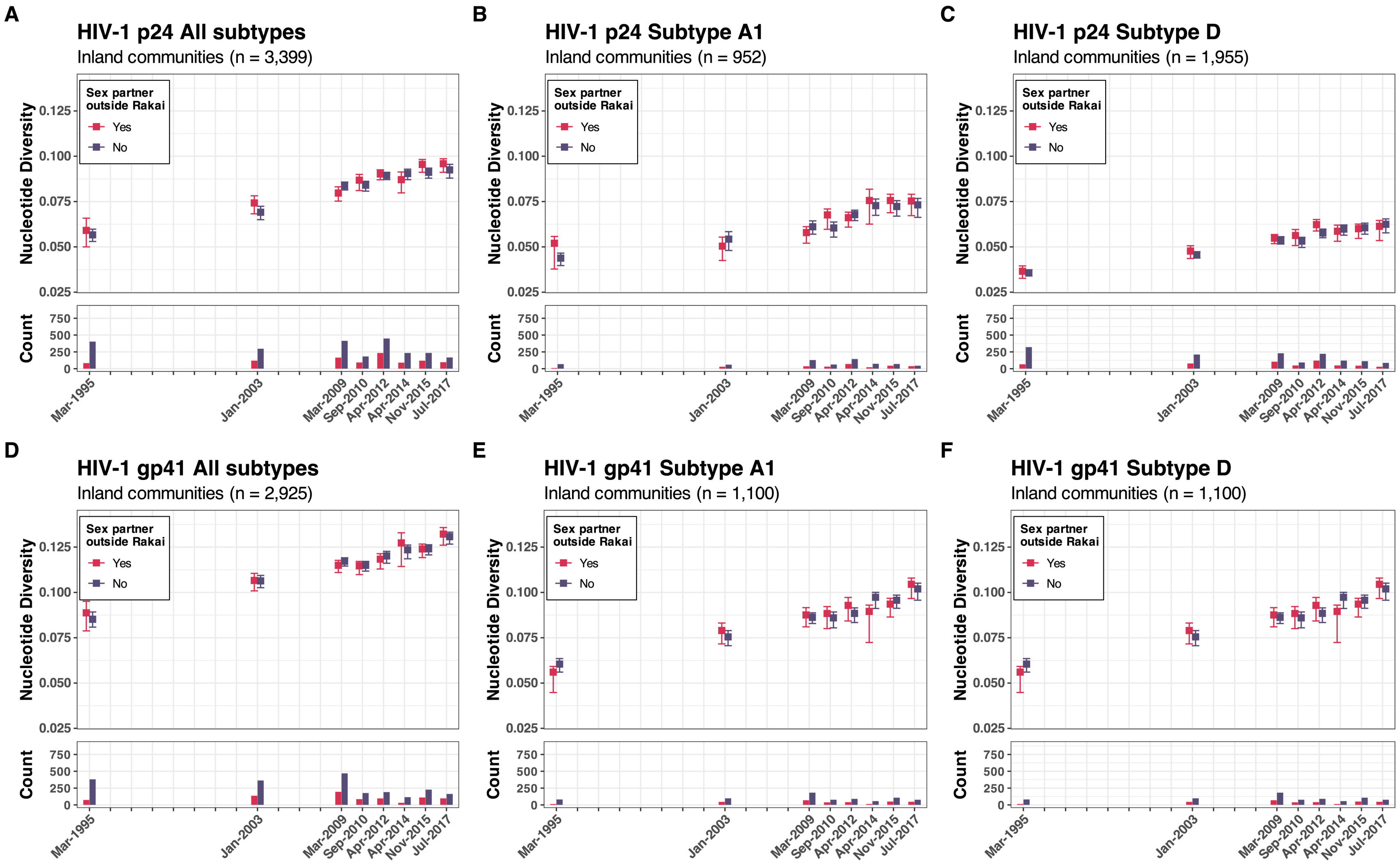


**Supplementary Figure 26.** The overall and subtype specific (A1 & D) nucleotide genetic diversity of p24 and gp41 by the presence of external sexual partners in the past year in 31 inland agrarian and semi-urban trading communities between 1995 and 2017

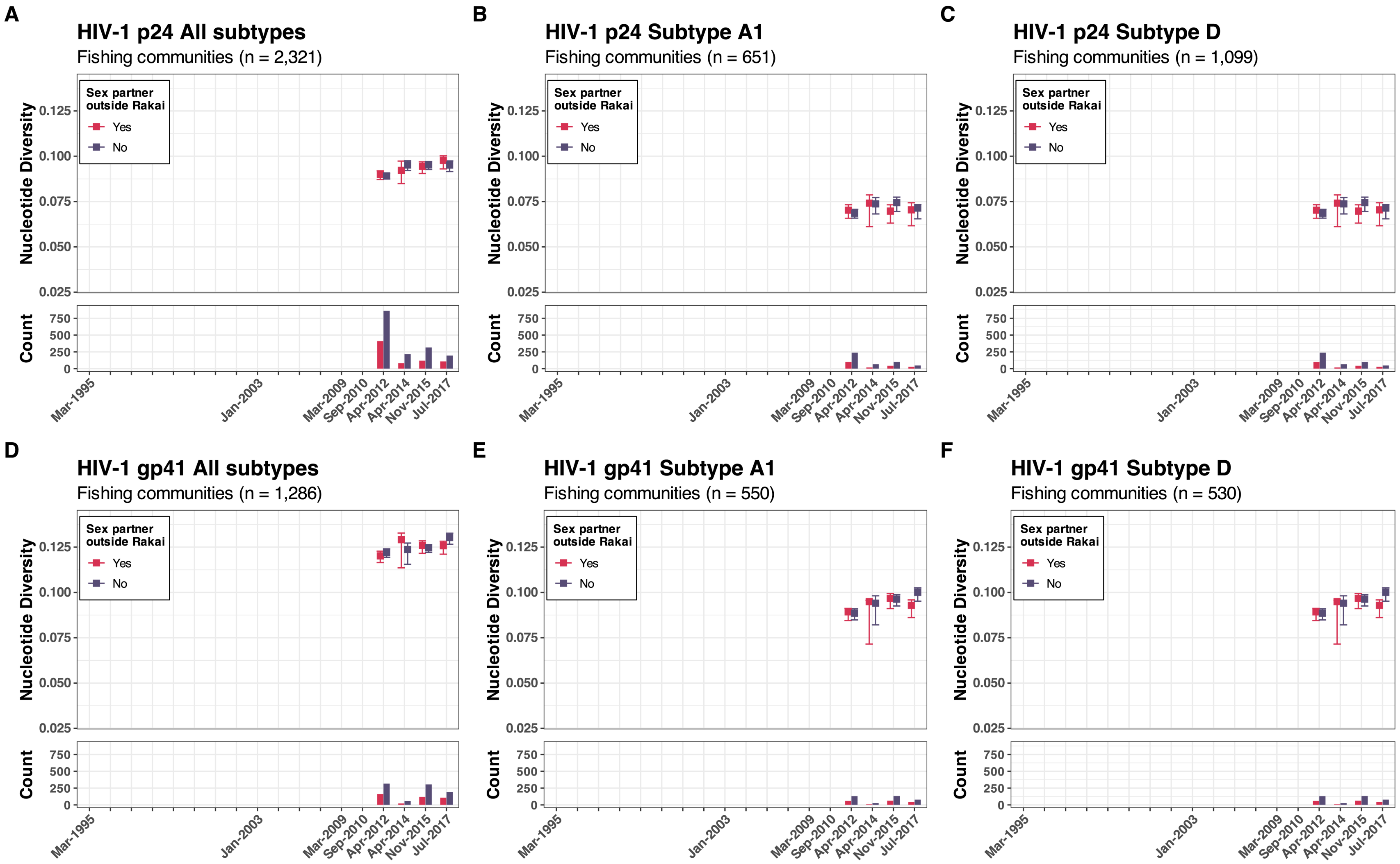


**Supplementary Figure 27.** The overall and subtype specific (A1 & D) nucleotide genetic diversity of p24 and gp41 by the presence of external sexual partners in the past year in four hyperendemic Lake Victoria fishing communities between 2012 and 2017

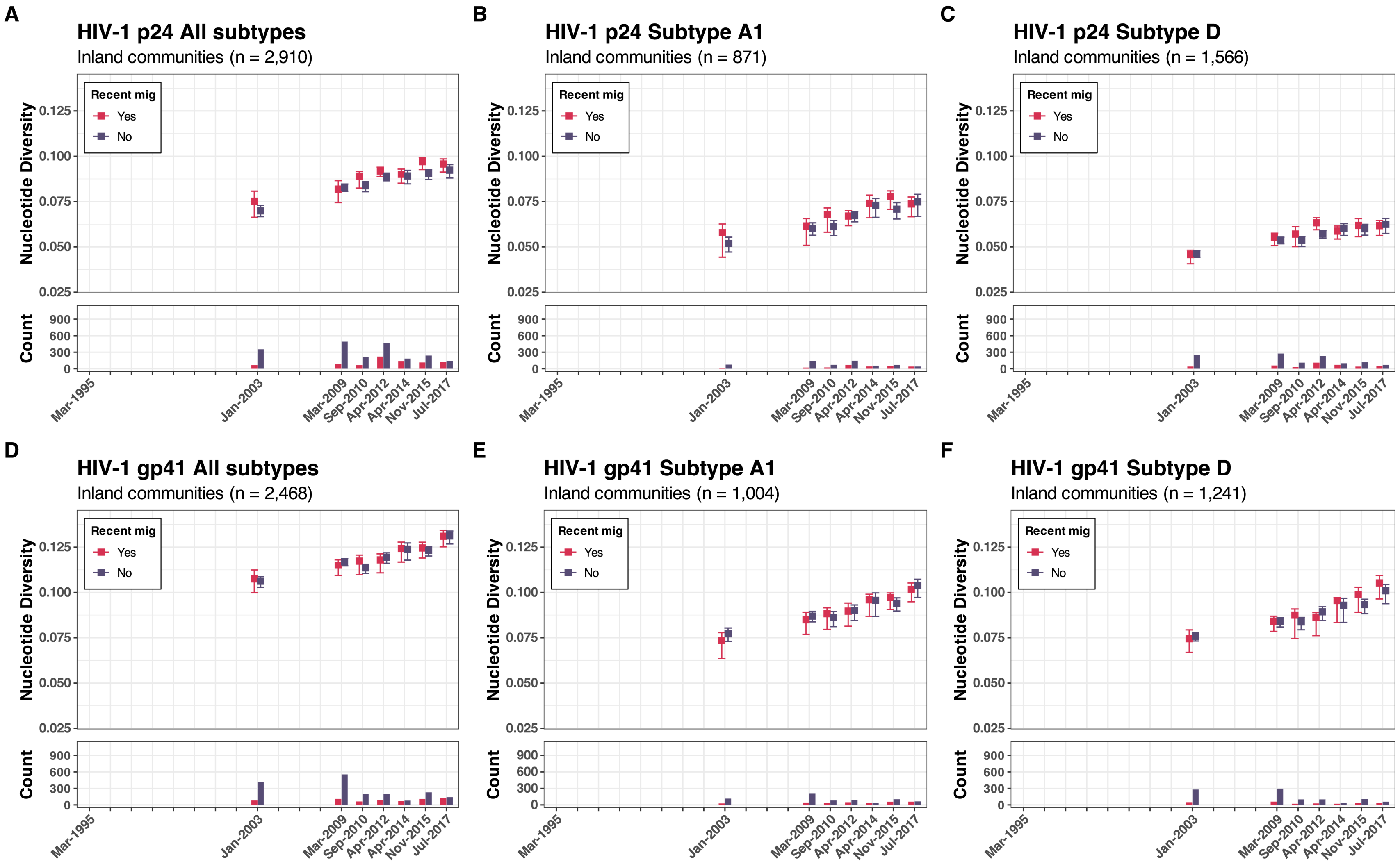


**Supplementary Figure 28.** The overall and subtype specific (A1 & D) nucleotide genetic diversity of p24 and gp41 by recent history of migration in the past year in 31 inland agrarian and semi-urban trading communities between 2003 and 2017

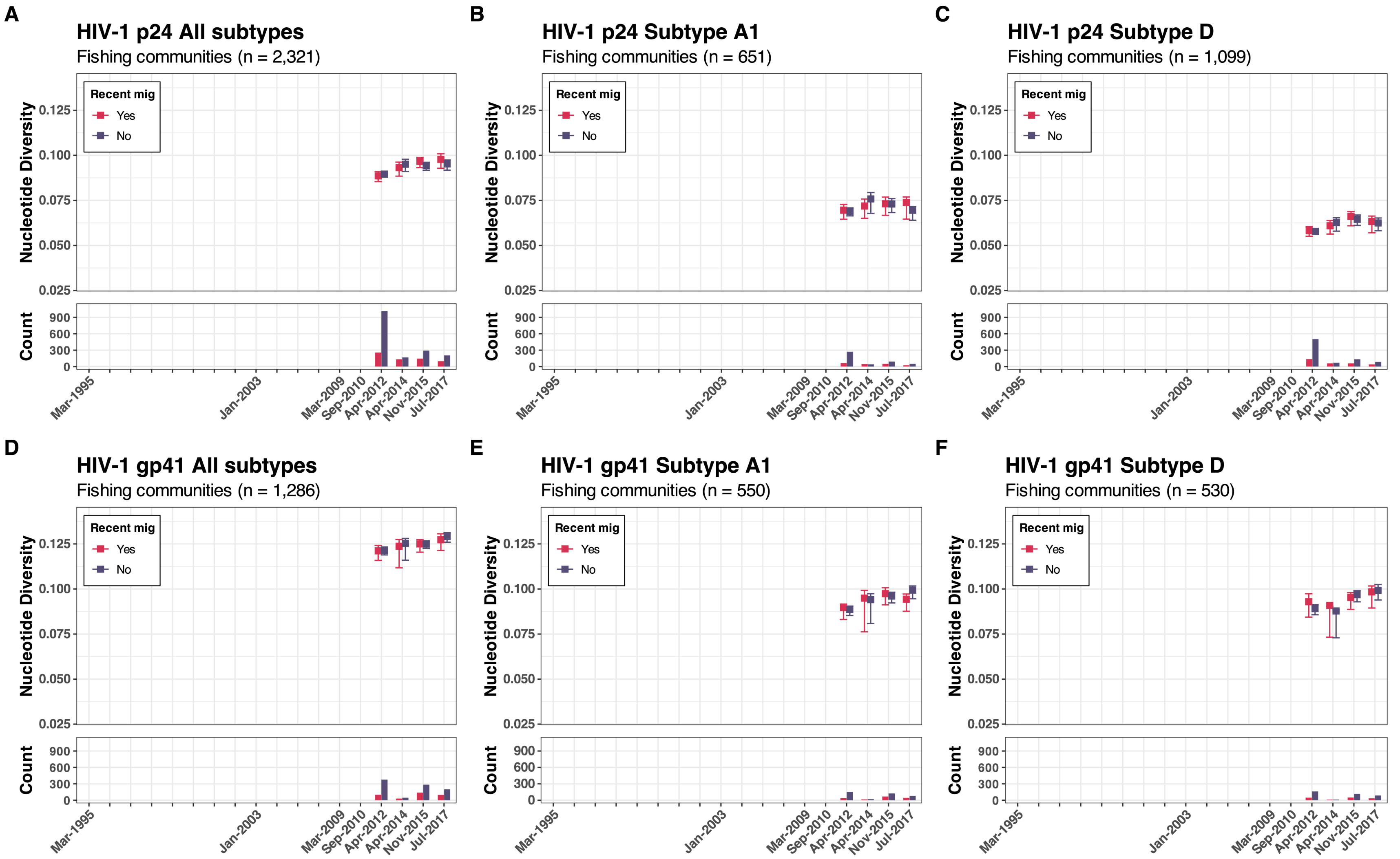


**Supplementary Figure 29.** The overall and subtype specific (A1 & D) nucleotide genetic diversity of p24 and gp41 by recent history of migration in the past year in four hyperendemic Lake Victoria fishing communities between 2012 and 2017

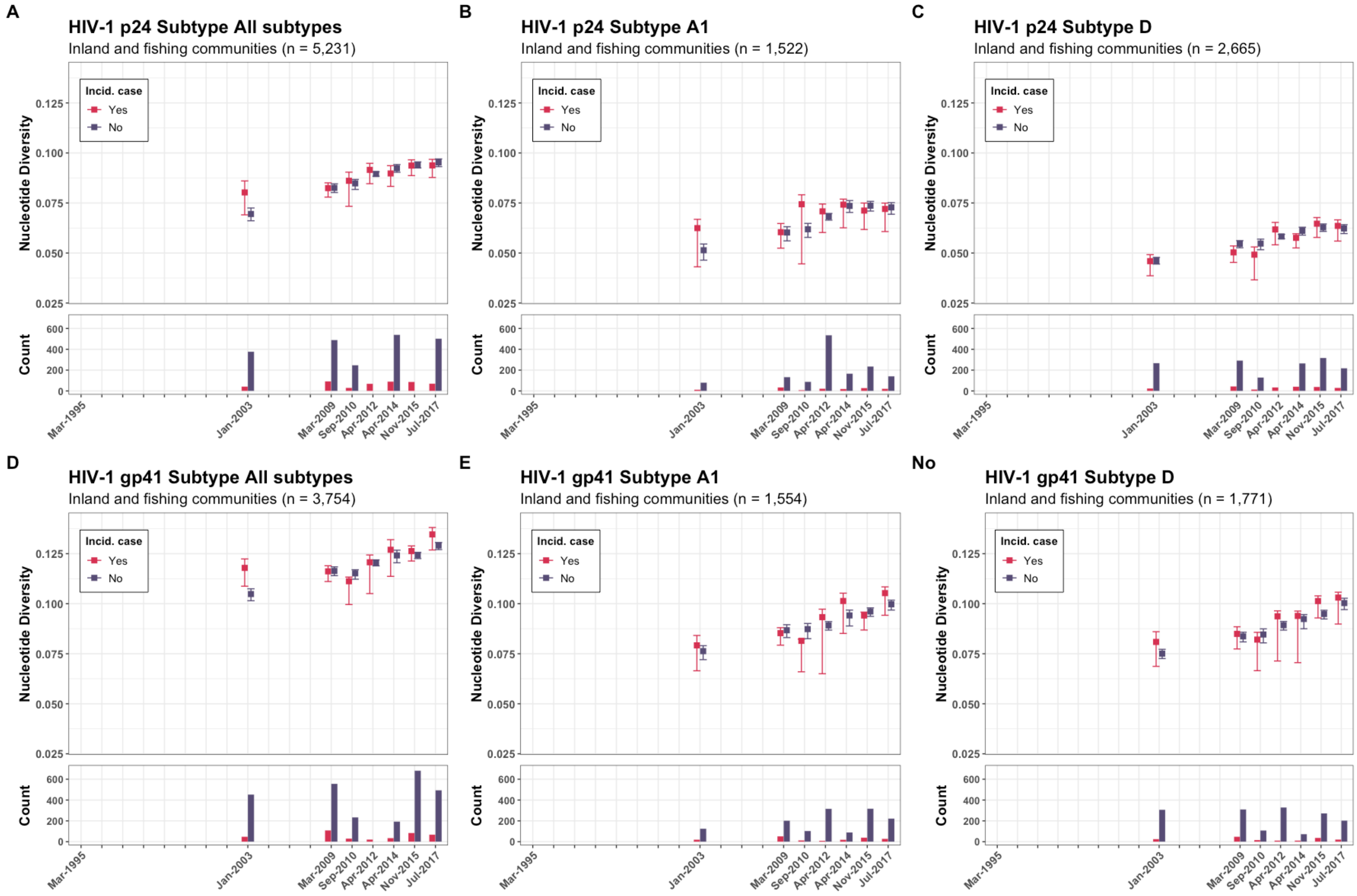


**Supplementary Figure 30.** The overall and subtype specific (A1 & D) nucleotide genetic diversity of p24 and gp41 by incident case in Rakai between 2003 and 2017

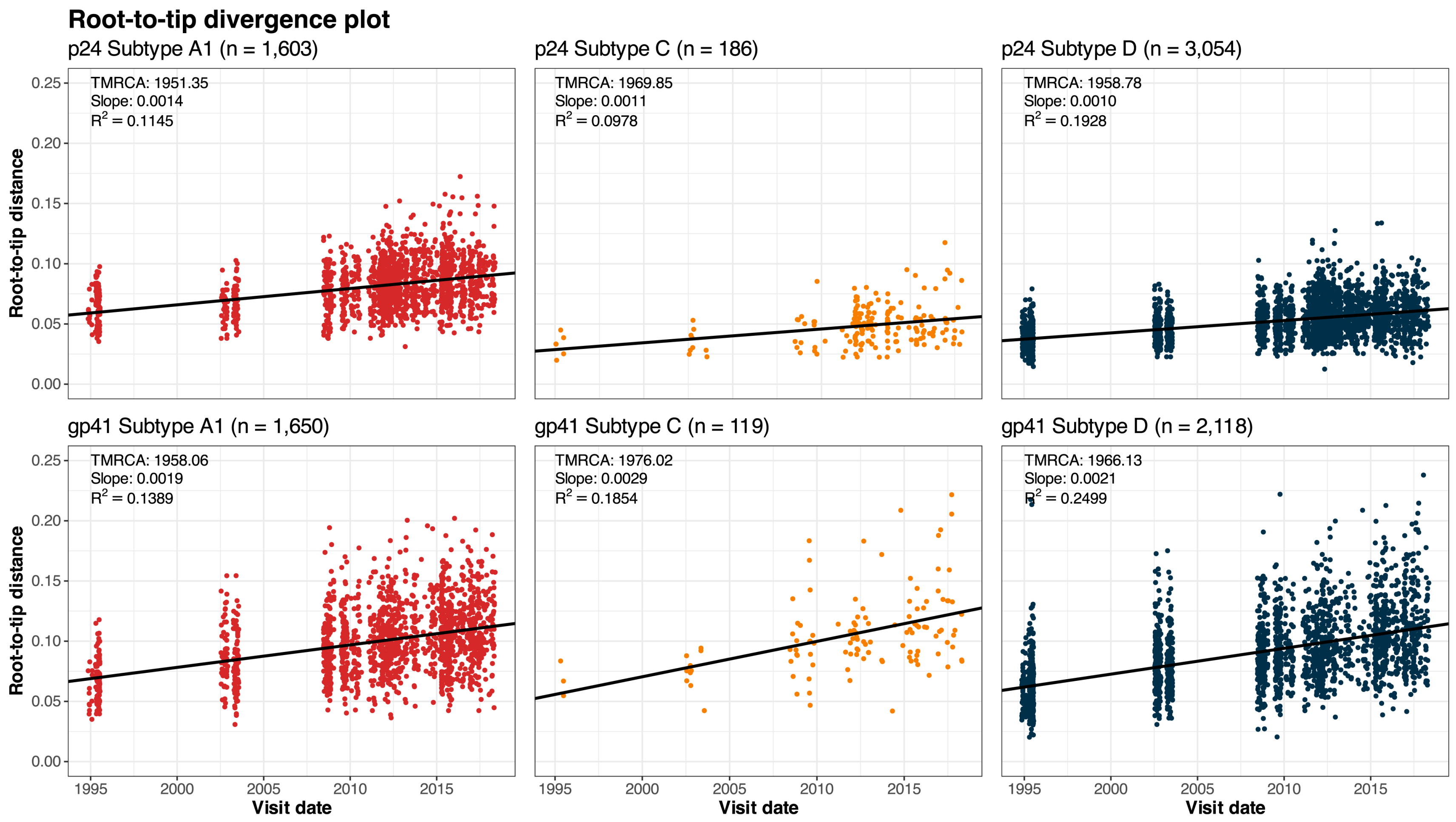


**Supplementary Figure 31.** Root-to-tip divergence analyses of HIV p24 and gp41 subtype A1, C, and D in Rakai Between 1995 and 2017

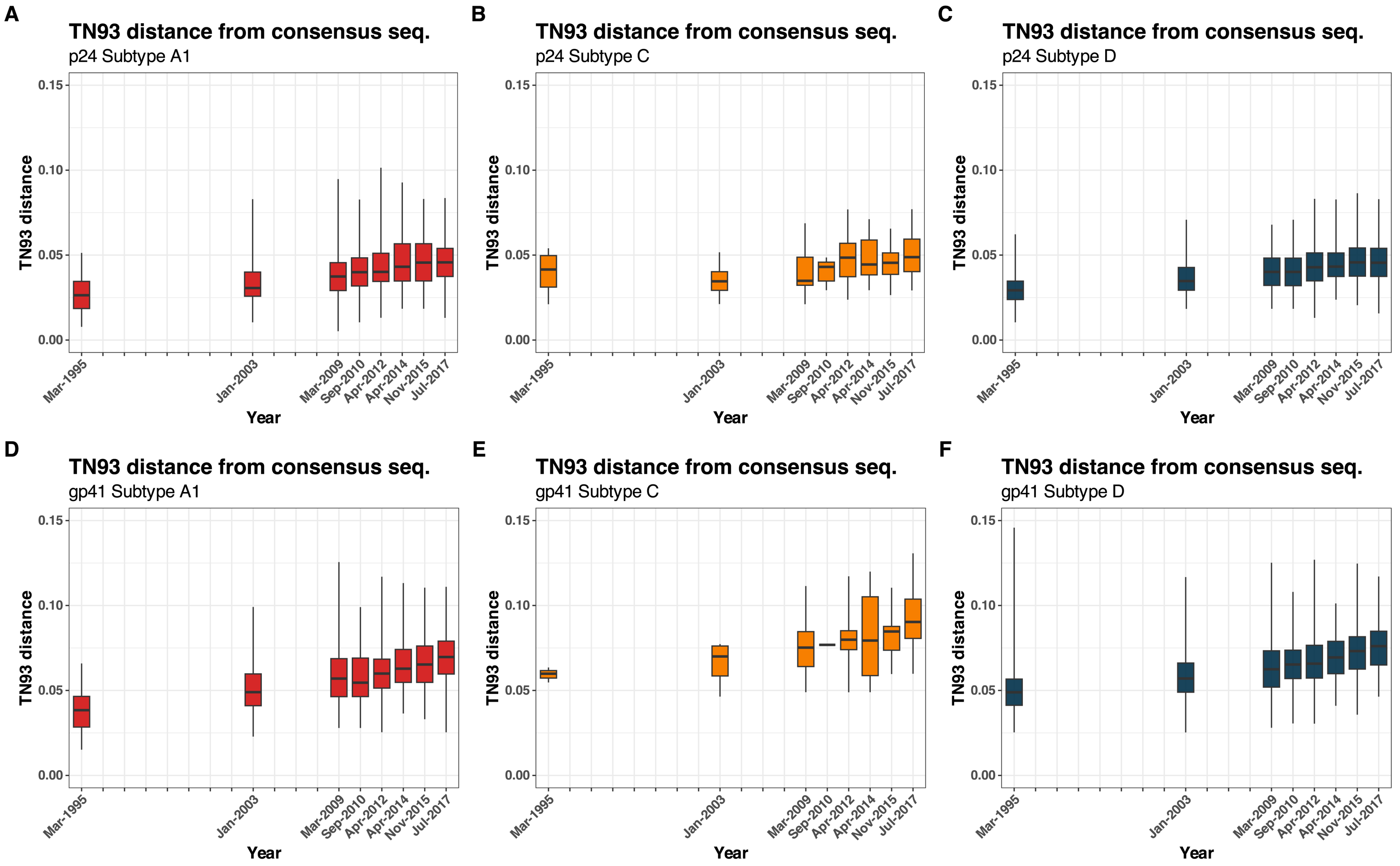


**Supplementary Figure 32.** Pairwise TN93 genetic distance between HIV subtype reference sequences (A1, C, and D) and HIV sequences in Rakai between 1995 to 2017

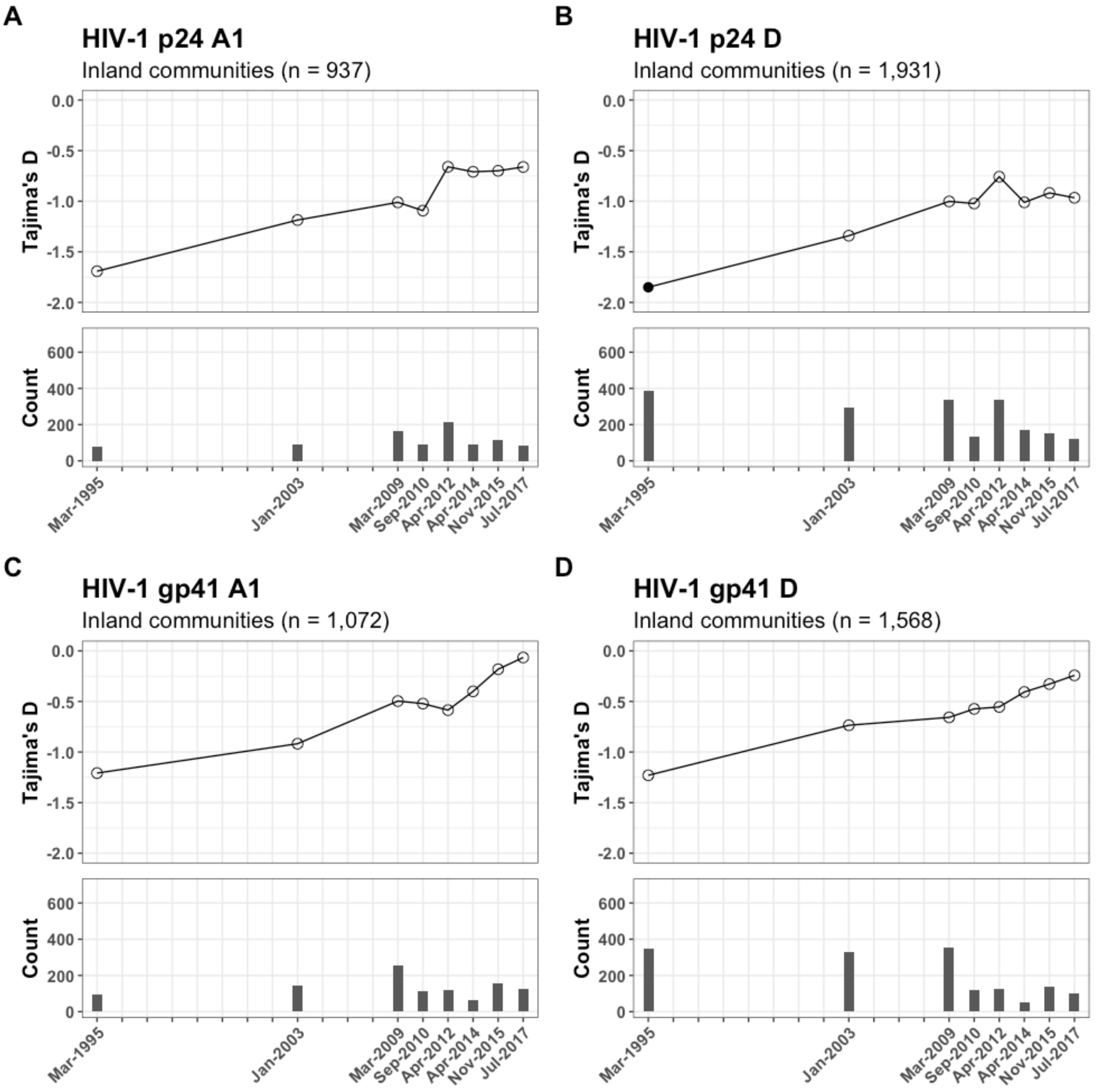


**Supplementary Figure 33.** Tajima's D in p24 and gp41 subtypes A1 and D in 31 inland agrarian and semi-urban trading communities between 1995 to 2017. Statistically significant D values with p-values of less than 0.05 under a beta distribution with a mean of zero and a variance of 1 are represented by filled circles or hollow circles otherwise.

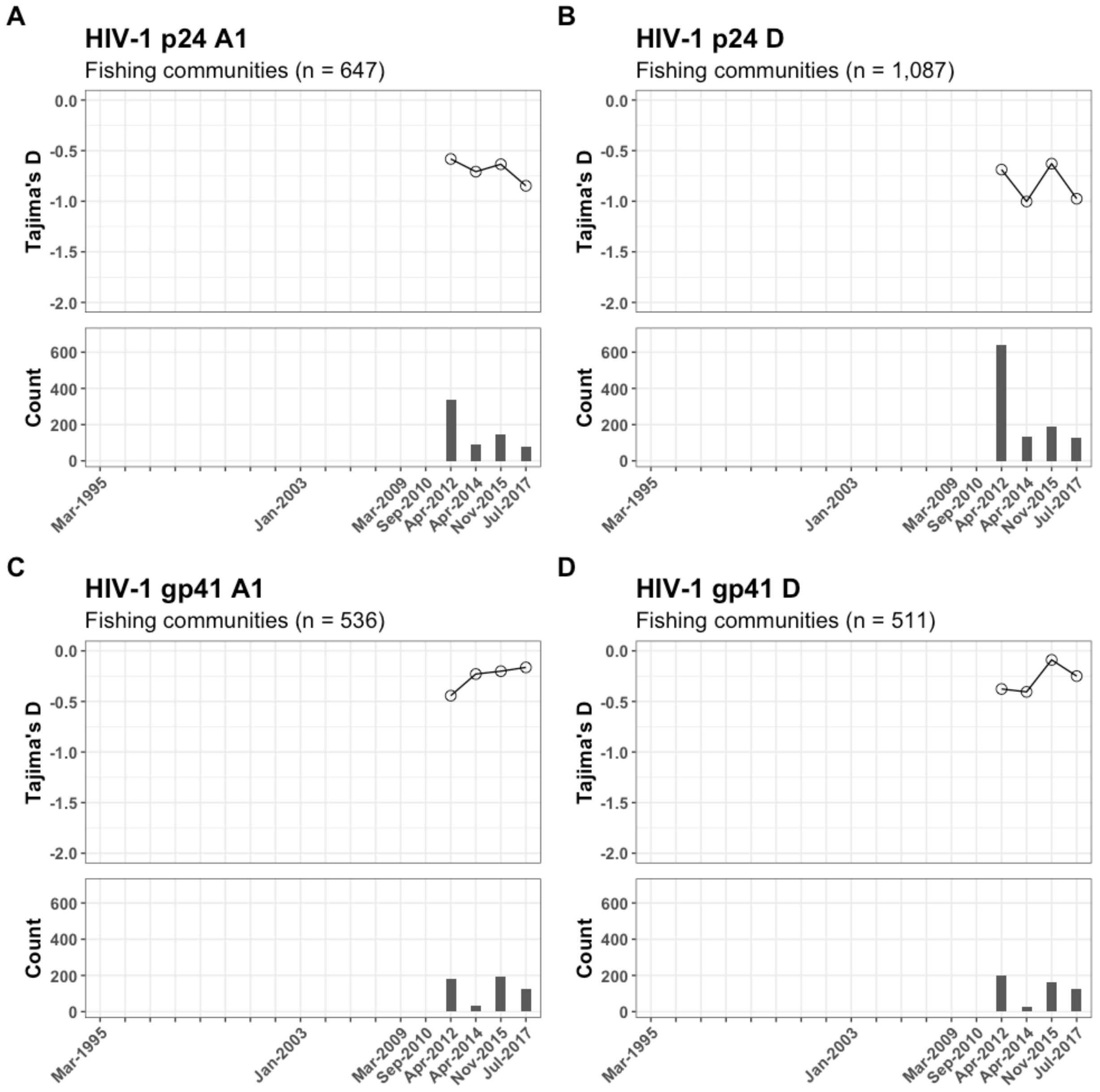


**Supplementary Figure 34.** Tajima's D in p24 and gp41 subtypes A1 and D in four hyperendemic Lake Victoria fishing communities between 2012 and 2017. Statistically significant D values with p-values of less than 0.05 under a beta distribution with a mean of zero and a variance of 1 are represented by filled circles or hollow circles otherwise.

**Supplementary Table 12.** dN/dS ratio in p24 and gp41 subtypes A1 and D from 31 inland agrarian and semi-urban trading communities and four hyperendemic Lake Victoria fishing communities

| **Metric** | **Inland communities** | | | | **Fishing communities** | | | |
|---|---|---|---|---|---|---|---|---|
| | **p24** | | **gp41** | | **p24** | | **gp41** | |
| | **A1** | **D** | **A1** | **D** | **A1** | **D** | **A1** | **D** |
| Number of sequences | 952 | 1,955 | 1,100 | 1,588 | 651 | 1,099 | 550 | 530 |
| **dN/dS ratio (ω)** | | | | | | | | |
| codeml | 0.15 | 0.12 | 0.47 | 0.52 | 0.15 | 0.12 | 0.45 | 0.48 |
| BUSTED | 0.18 | 0.14 | 0.49 | 0.60 | 0.18 | 0.14 | 0.47 | 0.55 |
